# Oropouche, Dengue, and Chikungunya differential diagnosis. Development and validation of predictive models with surveillance data from Espírito Santo/Brazil

**DOI:** 10.64898/2026.04.17.26350875

**Authors:** Elry Cristine Nickel Valerio, Gabriela Maria Coli Seidel, Rafael da Silva Nunes, Pedro Emmanuel Alvarenga Americano do Brasil

## Abstract

There is an ongoing Oropouche Fever (OF) outbreak in Brazil since 2024. There are dengue and chikungunya prediction models available, but none to help discriminate dengue, chikungunya, and OF.

**Objective:** This study aims to develop and validate clinical prediction models for dengue, chikungunya, OF.

**Methods:** This study uses surveillance data from Espírito Santo state / Brazil, from 2023-2025. Epidemiological investigations and biological samples were used to conclude cases as either (a) clinical-epidemiologically confirmed, (b) laboratory confirmed, or (c) “discarded”. The predictors were all data related to signs, symptoms, and comorbidities available in the notification forms. The analysis was performed using random forest regression models, one for each outcome, in development and validation datasets.

**Results:** A total of 465,280 observations were analyzed, 261,691 dengue cases (56.6%), 18,676 chikungunya cases (4.0%), 12,174 OF cases (2.6%), and 179,115 discarded cases (38.6%). All three models had good discrimination and moderate to good calibration after scaling prediction. The models retained from 26 to 16 predictors each. Leukopenia and vomiting were the most discriminatory predictors for dengue, arthritis, arthralgia, and rash were the most discriminatory for chikungunya, and epidemiological features were the most relevant for OF. The dengue, chikungunya, and OF models had ROC AUC of 0.726, 0.851, and 0.896 in the validation set, respectively.

**Conclusion:** This research identified predictors most discriminative between dengue, chikungunya, and OF. We developed and validated predictive models, one for each condition, with moderate to very good performance available at https://pedrobrasil.shinyapps.io/INDWELL/. One may use them in diagnostic work-up and arbovirus surveillance.

**LAY SUMMARY:**

- Dengue fever was by far the most common confirmed infection in the surveillance system (56.6%), followed by chikungunya cases (4.0%), oropouche cases (2.6%), zika cases (0.2%), with different seasonality.
- Although suspected dengue cases were more laboratory tested, laboratory confirmation was more frequent among oropouche fever cases. It was possible to develop and validate models for dengue, chikungunya, and oropouche diagnostic investigation, and their performance was from moderate to very good, with suggested decision limits to aid the users. All attempts to develop Zika models returned models with unacceptable performance.
- We developed an instrument to predict the three conditions with the same predictors and made it available for users at https://pedrobrasil.shinyapps.io/INDWELL/.

## INTRODUCTION

Oropouche fever (OF) has recently emerged as a significant arboviral disease in Brazil, with a marked geographic expansion beyond its historically endemic Amazon region. Since 2024, the country has experienced a substantial increase in reported cases, with transmission documented across multiple states, characterizing a large-scale epidemic scenario (1,2). According to the Pan American Health Organization, Brazil accounted for the majority of reported cases in the Americas, with over 13.000 confirmed infections in 2024 alone, underscoring the growing regional public health relevance of the disease (2). This expansion has been associated with environmental and socioecological drivers, including climate variability, deforestation, and urbanization, which may facilitate vector adaptation and viral spread into new territories (3,4).

Oropouche virus (OROV), of the Peribunyaviridae family, is the OF agent, and it is transmitted mainly by the mosquito *Culicoides paraensis*. Although prevalent in the Brazilian north region, the recent circulation of OROV in the state of Espírito Santo (southeast) constitutes a new epidemiological scenario. At the time, this novelty faced the absence of a specific clinical and surveillance protocols, and it was incorporated in the arboviruses surveillance effort due to the similarity of symptoms with other arboviral diseases (5–7).

In Brazil, the Espírito Santo state has played a central role in this new epidemiological scenario, accounting for approximately 45% of all reported cases in Brazil, with more than 6.000 confirmed infections. (1,8). This has changed the context of arboviruses diagnostic investigation and surveillance, as it is frequently hard to clinically distinguish dengue, chikungunya, OF, and OF (and other arboviruses) (8). Fever, headache, myalgia, and rash are common to all three conditions.

Dengue presents with a broad and heterogeneous symptom spectrum, usually with fever (>95% of the cases), headache and retro-orbital pain are more common among adults then children, myalgia (67.5%), arthralgia (typically less severe and less prolonged than in chikungunya), rash (20.5%) thrombocytopenia (61.4%), leucopenia (9–12). Chikungunya is primarily suspected with joint involvement (dengue affects vasculature, chikungunya targets joints) (13,14), which key features are fever (>95%) often is biphasic, severe polyarthralgia, morning stiffness (82.4%), joint swelling (35.3%), with elbow and knee being the most commonly affected joints, rash (maculopapular), myalgia (11,15,16). OF is mainly manifested as fever (97%) of cases, headache (86%), myalgia (69%), arthralgia (57%), dizziness/vertigo (frequently reported), odynophagia (sore throat) (28%), and abdominal pain (15%) (9,17,18). Across arboviruses, systematic reviews have consistently highlighted key challenges of diagnostic strategies, including co-infections (around 2.1% globally) (19), substantial heterogeneity between studies, variability in laboratory reference standards, differences in timing of sample collection, and limited representation of real-world primary care settings (2,12,19,20)

The discrimination with signs and symptoms between dengue and chikungunya has been the most extensively studied, with several examples (14,16,17,21–25). It seems that severe polyarthralgia with morning stiffness and joint swelling, thrombocytopenia (<100,000 cells/mL), hemorrhagic manifestations (petechiae, bleeding), and duration of joint symptoms are the most discriminative features (10,11,15,21). We could not find many attempts to discriminate OF from chikungunya, but here is some data regarding dengue and OF discrimination. Fever and headache do not discriminate between them, however, odynophagia and abdominal pain are more frequent in OF, while myalgia, arthralgia, and rash are less frequent in OF than in dengue (3,9).

There is considerable evidence regarding models predicting the probability of arboviral diseases through signs and symptoms, or different combinations of signs and symptoms, or to differentiate them with the aid of clinical prediction models before confirmatory tests are requested. But apparently, the exercise of differential diagnosis with prediction models is incipient (26). The co-circulation and features in common usually explain the models’ variable performance and turn the discussion into reinforcing laboratory work-up (2). Additionally, we were only able to find one study attempting to estimate the probability of OF through signs and symptoms developed in Peru without an evident applicability to the Brazilian context (27).

Therefore, it is opportune to investigate the differential diagnosis of OF from other arboviruses in our context, creating the possibility of assisting diagnostic investigation in suspected cases, supporting management protocols, surveillance actions, and control measures. This research aims to develop and validate prediction models containing simple signs and symptoms information from surveillance records from March 2023 to July 2025, in an attempt to help discriminate OF, dengue fever, and chikungunya in the diagnostic workup.

## METHOD

### Ethical aspects

The project was submitted to and approved by the Research Ethics Committee of INI-FIOCRUZ, CAAE: 92760425.0.0000.5262, (https://plataformabrasil.saude.gov.br/login.jsf) Number: 7.976.813 on 13/11/2025, in accordance with CNS Resolution No. 466/2012 and subsequent resolutions. The project had prior approval from the Special Center for Health Information Systems (Nesis) of the State Health Secretariat of Espírito Santo-ES, process n°. 2025-61G4S. The informed consent form (ICF) was waived, as it involves the use of a secondary data source, in the public domain, primarily used for the surveillance of notifiable diseases, and which was made available in an anonymized form by Nesis.

### Data source

The surveillance process of arboviruses in Espírito Santo follows the national guidelines and is adapted to state requirements. Arbovirus surveillance begins in local health units, where patients presenting with fever associated with at least two nonspecific symptoms, such as headache and myalgia, are considered suspected cases. At this stage, a standardized electronic notification form is completed, which includes clinical and sociodemographic data as well as the date of symptom onset. The case is then classified as “under investigation”, initially registered as Dengue due to its predominance among arboviruses. Simultaneously, an epidemiological investigation is initiated (8).

Notification must be accompanied by the collection of biological samples—blood serum, urine, or cerebrospinal fluid - which are sent by municipalities to LACEN-ES (Central Public Health Laboratory of Espírito Santo state). The laboratory performs analyses using RT-qPCR and serology, following a prioritization sequence flow diagram: first dengue, zika, and chikungunya; then OF and yellow fever; and finally West Nile fever and Mayaro fever. This hierarchy is essential, as Espírito Santo faces seasonal variations and overlapping seasonality (8).

After laboratory confirmation, the notification form is reassessed, and the case is reclassified as “concluded”, consolidating the data within the state surveillance system. When results confirm dengue or chikungunya, the case is closed directly in the initial form. In cases where the final diagnosis corresponds to zika, yellow fever, mayaro fever, west nile fever, or OF, the initial dengue notification is reclassified as discarded, and a new notification is opened, now classified as a “concluded notification” for the confirmed etiological agent (28,29);

### Participants

Patients presenting in public and private health units with fever associated with at least two nonspecific symptoms, such as headache and myalgia, are considered suspected cases. Suspected, reported, and investigated cases of arboviruses registered in the Espírito Santo state surveillance system, including unconfirmed cases, have their biological samples collected to perform laboratory confirmation (RT-PCR or serology) at LACEN-ES, including for other conditions (investigated simultaneously) such as dengue, zika, chikungunya, and west nile fever (30).

### Outcomes

The investigation can be closed in two ways: (a) clinical-epidemiological confirmed-when there is an epidemiological link with laboratory-confirmed cases in the patient’s territory, or when the patient has traveled to areas with circulation of symptomatic individuals, presenting a clinical picture compatible with arbovirus infection; (b) laboratory confirmed - when there is direct detection of the virus through molecular biology (RT-PCR) or the presence of specific IgM antibodies, associated with an epidemiological link. If the investigation does not come to a conclusion towards any of the initially suspected conditions, then the case is reclassified as “discarded” (28).

### Predictors

Data used as predictors were all data related to signs, symptoms, and co-morbidities available in the dengue notifications form, which was long optimized for Dengue and Chikungunya. After the OF outbreak, there was no further optimization of this data collection. The guidance on how to fill these forms may change over time, but there was no specific recommendation regarding signs and symptoms after OF outbreak. The predictors available and used were first symptoms week, age at notification, sex at birth, pregnancy, race or skin color, education, address, fever, headache, vomiting, back pain, arthritis, petechia, tourniquet test, myalgia, exanthema, nausea, conjunctivitis, arthralgia, leukopenia, retro-orbital pain, diabetes, hepatopathy, arterial hypertension, autoimmune diseases, hematological diseases, chronic renal diseases, acid peptic diseases (31–33).

### Data preparation

The data was downloaded as provided from the Espirito Santo Health Secretary in spreadsheets (CSV) files. Three main datasets were accessed: one for dengue, one for chikungunya, and one for zika and OF. All datasets were checked for valid identifications (CNS number or *cartão nacional do SUS*), valid period of interest, absence of outcome, and replicated entries. Registries were considered replicated if they were from the same CNS number and had initial symptoms within 14 days or less apart. The dengue dataset was considered the main dataset, given that Dengue was considered the main arbovirus at the time and had all the initial signs and symptoms data. The Zika/Oropouche dataset was matched to the Chikungunya dataset by the CNS number and initial symptoms date, and later this data was matched to the Dengue dataset with the same rationale (Figure 1)

**Figure 1:**
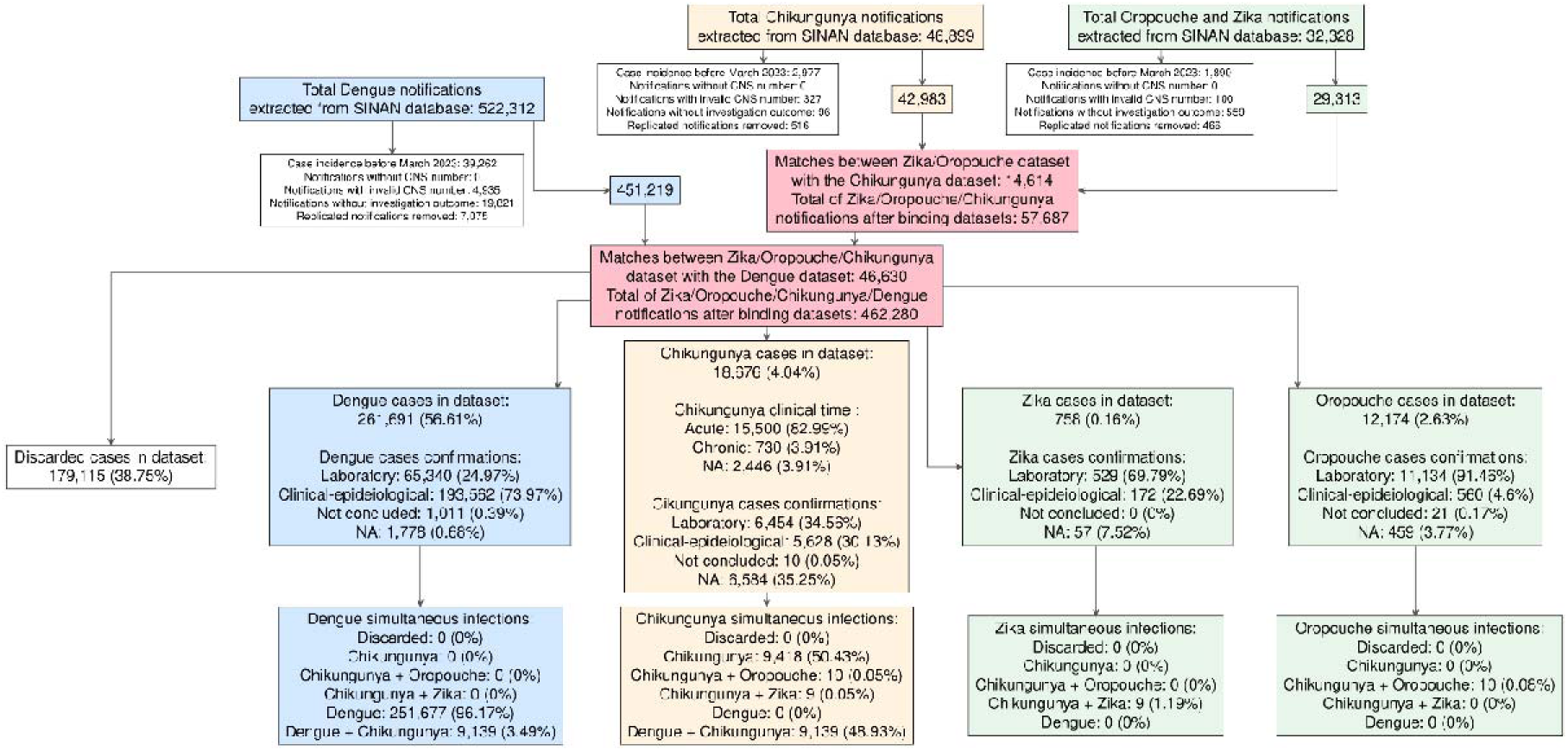
Inclusion, exclusion, and datasets merge flow diagram.

### Missing data and analysis plan

The analysis plan was performed using R-project software with packages tidyverse, tsibble, DiagrammeR, labelled, table1, flextable, missForestPredict, parallel, doParallel, Boruta, ranger, givityR, DiagnosisMed, UncertainInterval. First, the data was randomly marked into development/train data and validation/test data, so that the development had two-thirds, and the validation had one-third of the original observations. After selecting potential predictors, missing data patterns were explored (supplementary file) and imputed using random forest models fitted with random samples of 3.000 random observations from each outcome, and later predicted in the main data to fill the missing values.

The analysis was performed using random forest regression models, one for each outcome, therefore predicting probabilities of the outcome. As there was a considerable chance of co-infection (an observation having more than one outcome simultaneously), thus, a multiclass approach was considered less reasonable. The optimal value for the number of variables randomly selected in each tree division (*mtry* hyperparameter) was determined starting from a default value of 15, then a heuristic search by evaluating the Out-of-Bag (OOB) error for different predictor subset sizes. The process continued iteratively, adjusting the *mtry* value by a specified step factor until the improvement in the OOB error became negligible. The class imbalance was addressed by setting weights to observations with and without the outcome inversely proportional to the outcome density. Then, a sample size of 16.250 was set for each tree; therefore, a random sample of approximately 8.125 cases and non-cases should be selected with replacement in each of the 350 sampling generations.

Predictors for the final model were selected using iterative comparison of their relative importances with the importances of shadow attributes. Shadow attributes are created by shuffling original ones. Predictors that have significantly worse normalized permutation importance than shadow ones are consecutively dropped, and attributes that are significantly better than shadows are retained. The marginal effect of each selected predictor was estimated while maintaining fixed values of the remaining predictors of the model, and estimating the relative average effects of the predictions of the model. This approach allowed for the visualization of the functional relationship between the predictors and the predicted outcome.

After model fit, the predictions were scaled adjusted with the Platt method to avoid miscalibrated predictions. Performance was assessed in the train and test datasets using ROC AUC, R^2^, model intercept and slope, and prediction errors. Additionally, an uncertain range (using 0.55 error from mixed densities intersections) and an inconclusive range (using at least the 0.80 sensitivity and specificity threshold) were estimated using the scaled predictions. The predictions were dichotomized at the sensitivity equals the specificity threshold, and performance for a binary classification was estimated using this threshold. At last, we built a shiny app with these models for users to make predictions, available at https://pedrobrasil.shinyapps.io/INDWELL/.

## RESULTS

From the initial datasets summing more than 600,000 observations, 465,280 were analyzed after removing cases out of the period of interest, without CNS unique identifier, replicated registries, and binding the different datasets (Figure 1) In this merged final dataset, there were 261,691 dengue cases (56.6%), 18,676 chikungunya cases (4.0%), 758 zika cases (0.2%), 12,174 OF cases (2.6%), and 179,115 (38.6%) did not match any of these outcomes, therefore classified as “discarded”. Dengue fever was mostly diagnosed matching the clinical-epidemiological criteria; on the other hand, the remaining outcomes were mostly reached with laboratory confirmation, with an exceptionally high rate considering the OF cases. Chikungunya data were the most incomplete regarding how confirmation was reached and had the highest degree of co-infection with Dengue fever, reaching almost half of all Chikungunya cases (Figure 1).

Dengue and discarded cases had a lower median age, were more evenly distributed by sex, and were more frequent among black individuals. Urban living was also more frequent among the dengue and discarded cases. Most of the signs and symptoms were not evenly distributed among the different outcomes. Some comorbidities were evidently more frequent among chikungunya cases, for example, diabetes, arterial hypertension, and autoimmune diseases (Table 1).

**Table 1-.** Final dataset variables (including epidemiological, clinical at presentations) counts and frequencies by all arboviruses diagnostic investigation outcome.

|  | **Discarded**<br>**(N=179115)** | **Dengue**<br>**(N=261691)** | **Chikungunya**<br>**(N=18676)** | **Zika**<br>**(N=758)** | **Oropouche**<br>**(N=12174)** |
| --- | --- | --- | --- | --- | --- |
| **Age at notification** |  |  |  |  |  |
| Mean (SD) | 34.2 (19.6) | 35.4 (18.6) | 43.0 (19.0) | 40.1 (18.0) | 42.0 (18.2) |
| Median [Min, Max] | 32.0 [0, 106] | 33.0 [0, 104] | 43.0 [0, 98.0] | 40.0 [0, 90.0] | 41.0 [0, 99.0] |
| Missing | 3,991 (2.2%) | 528 (0.2%) | 6,569 (35.2%) | 57 (7.5%) | 412 (3.4%) |
| **Age at notification** |  |  |  |  |  |
| [0,1] | 3,402 (1.9%) | 1,900 (0.7%) | 20 (0.1%) | 3 (0.4%) | 10 (0.1%) |
| (1,12] | 20,867 (11.7%) | 23,448 (9.0%) | 663 (3.6%) | 46 (6.1%) | 448 (3.7%) |
| (12,18] | 16,688 (9.3%) | 25,041 (9.6%) | 706 (3.8%) | 50 (6.6%) | 770 (6.3%) |
| (18,25] | 24,893 (13.9%) | 41,307 (15.8%) | 1,105 (5.9%) | 66 (8.7%) | 1,270 (10.4%) |
| (25,35] | 31,731 (17.7%) | 49,094 (18.8%) | 1,829 (9.8%) | 115 (15.2%) | 2,038 (16.7%) |
| (35,45] | 28,890 (16.1%) | 45,025 (17.2%) | 2,242 (12.0%) | 149 (19.7%) | 2,313 (19.0%) |
| (45,55] | 21,066 (11.8%) | 32,995 (12.6%) | 2,116 (11.3%) | 113 (14.9%) | 1,960 (16.1%) |
| (55,65] | 14,642 (8.2%) | 23,666 (9.0%) | 1,880 (10.1%) | 106 (14.0%) | 1,619 (13.3%) |
| (65,75] | 8,526 (4.8%) | 13,092 (5.0%) | 1,075 (5.8%) | 40 (5.3%) | 930 (7.6%) |
| (75,85] | 3,447 (1.9%) | 4,528 (1.7%) | 409 (2.2%) | 12 (1.6%) | 331 (2.7%) |
| (85,95] | 922 (0.5%) | 1,015 (0.4%) | 61 (0.3%) | 1 (0.1%) | 70 (0.6%) |
| Missing | 4,041 (2.3%) | 580 (0.2%) | 6,570 (35.2%) | 57 (7.5%) | 415 (3.4%) |
| **Sex at notification** |  |  |  |  |  |
| Female | 94,159 (52.6%) | 142,642 (54.5%) | 7,535 (40.3%) | 399 (52.6%) | 5,448 (44.8%) |
| Male | 80,956 (45.2%) | 118,514 (45.3%) | 4,572 (24.5%) | 302 (39.8%) | 6,313 (51.9%) |
| Ignored | 9 (0.0%) | 7 (0.0%) | 0 (0%) | 0 (0%) | 1 (0.0%) |
| Missing | 3,991 (2.2%) | 528 (0.2%) | 6,569 (35.2%) | 57 (7.5%) | 412 (3.4%) |
| **Pregnancy** |  |  |  |  |  |
| No | 161,266 (90.0%) | 228,559 (87.3%) | 11,003 (58.9%) | 642 (84.7%) | 11,004 (90.4%) |
| Yes | 2,250 (1.3%) | 2,575 (1.0%) | 228 (1.2%) | 24 (3.2%) | 246 (2.0%) |
| Ignored/Unknown | 11,608 (6.5%) | 30,029 (11.5%) | 876 (4.7%) | 35 (4.6%) | 512 (4.2%) |
| Missing | 3,991 (2.2%) | 528 (0.2%) | 6,569 (35.2%) | 57 (7.5%) | 412 (3.4%) |
| **Race or skin color** |  |  |  |  |  |
| White | 62,975 (35.2%) | 68,668 (26.2%) | 3,104 (16.6%) | 290 (38.3%) | 6,595 (54.2%) |
| Black | 89,629 (50.0%) | 145,605 (55.6%) | 7,317 (39.2%) | 250 (33.0%) | 3,880 (31.9%) |
| Yellow | 16,202 (9.0%) | 23,399 (8.9%) | 1,081 (5.8%) | 76 (10.0%) | 1,279 (10.5%) |
| Indigenous | 207 (0.1%) | 210 (0.1%) | 10 (0.1%) | 0 (0%) | 8 (0.1%) |
| Ignored/Unknown | 6,111 (3.4%) | 23,281 (8.9%) | 595 (3.2%) | 85 (11.2%) | 0 (0%) |
| Missing | 3,991 (2.2%) | 528 (0.2%) | 6,569 (35.2%) | 57 (7.5%) | 412 (3.4%) |
| **Indigenous ethnicity** |  |  |  |  |  |
| Tupiniquim | 117 (0.1%) | 44 (0.0%) | 0 (0%) | 0 (0%) | 1 (0.0%) |
| Guarani | 20 (0.0%) | 23 (0.0%) | 0 (0%) | 0 (0%) | 0 (0%) |
| Ignored/Unknown | 70 (0.0%) | 143 (0.1%) | 10 (0.1%) | 0 (0%) | 7 (0.1%) |
| Not applicable | 174,917 (97.7%) | 260,953 (99.7%) | 12,097 (64.8%) | 701 (92.5%) | 11,754 (96.6%) |
| Missing | 3,991 (2.2%) | 528 (0.2%) | 6,569 (35.2%) | 57 (7.5%) | 412 (3.4%) |
| **Person with special needs** |  |  |  |  |  |
| Yes | 83 (0.0%) | 374 (0.1%) | 46 (0.2%) | 1 (0.1%) | 1 (0.0%) |
| No | 175,041 (97.7%) | 260,789 (99.7%) | 12,061 (64.6%) | 700 (92.3%) | 11,761 (96.6%) |
| Missing | 3,991 (2.2%) | 528 (0.2%) | 6,569 (35.2%) | 57 (7.5%) | 412 (3.4%) |
| **Homeless** |  |  |  |  |  |
| Yes | 21 (0.0%) | 124 (0.0%) | 8 (0.0%) | 0 (0%) | 0 (0%) |
| No | 175,103 (97.8%) | 261,039 (99.8%) | 12,099 (64.8%) | 701 (92.5%) | 11,762 (96.6%) |
| Missing | 3,991 (2.2%) | 528 (0.2%) | 6,569 (35.2%) | 57 (7.5%) | 412 (3.4%) |
| **Education** |  |  |  |  |  |
| Illiterate | 9,907 (5.5%) | 9,906 (3.8%) | 601 (3.2%) | 52 (6.9%) | 956 (7.9%) |
| Elementary School Complete | 21,703 (12.1%) | 22,523 (8.6%) | 1,388 (7.4%) | 109 (14.4%) | 2,077 (17.1%) |
| Middle School Complete | 19,245 (10.7%) | 21,532 (8.2%) | 1,036 (5.5%) | 72 (9.5%) | 1,510 (12.4%) |
| High School Complete | 32,298 (18.0%) | 43,553 (16.6%) | 2,545 (13.6%) | 167 (22.0%) | 2,684 (22.0%) |
| Higher Education Complete | 8,797 (4.9%) | 12,164 (4.6%) | 803 (4.3%) | 89 (11.7%) | 729 (6.0%) |
| Ignored/Unknown | 83,174 (46.4%) | 151,485 (57.9%) | 5,734 (30.7%) | 212 (28.0%) | 3,806 (31.3%) |
| Missing | 3,991 (2.2%) | 528 (0.2%) | 6,569 (35.2%) | 57 (7.5%) | 412 (3.4%) |
| **Home address** |  |  |  |  |  |
| Urban | 119,650 (66.8%) | 224,116 (85.6%) | 10,937 (58.6%) | 467 (61.6%) | 5,579 (45.8%) |
| Rural | 38,198 (21.3%) | 24,978 (9.5%) | 565 (3.0%) | 159 (21.0%) | 5,193 (42.7%) |
| Peri-urban | 398 (0.2%) | 962 (0.4%) | 13 (0.1%) | 6 (0.8%) | 68 (0.6%) |
| Unknown | 16,878 (9.4%) | 11,101 (4.2%) | 592 (3.2%) | 69 (9.1%) | 922 (7.6%) |
| Missing | 3,991 (2.2%) | 534 (0.2%) | 6,569 (35.2%) | 57 (7.5%) | 412 (3.4%) |
| **Fever** |  |  |  |  |  |
| No | 55,707 (31.1%) | 45,381 (17.3%) | 5,814 (31.1%) | 136 (17.9%) | 1,763 (14.5%) |
| Yes | 123,071 (68.7%) | 216,310 (82.7%) | 12,862 (68.9%) | 577 (76.1%) | 10,026 (82.4%) |
| Missing | 337 (0.2%) | 0 (0%) | 0 (0%) | 45 (5.9%) | 385 (3.2%) |
| **Headache** |  |  |  |  |  |
| No | 52,406 (29.3%) | 51,298 (19.6%) | 7,159 (38.3%) | 139 (18.3%) | 1,987 (16.3%) |
| Yes | 126,372 (70.6%) | 210,393 (80.4%) | 11,517 (61.7%) | 574 (75.7%) | 9,802 (80.5%) |
| Missing | 337 (0.2%) | 0 (0%) | 0 (0%) | 45 (5.9%) | 385 (3.2%) |
| **Vomiting** |  |  |  |  |  |
| No | 141,018 (78.7%) | 206,468 (78.9%) | 16,163 (86.5%) | 524 (69.1%) | 10,050 (82.6%) |
| Yes | 37,760 (21.1%) | 55,223 (21.1%) | 2,513 (13.5%) | 189 (24.9%) | 1,739 (14.3%) |
| Missing | 337 (0.2%) | 0 (0%) | 0 (0%) | 45 (5.9%) | 385 (3.2%) |
| **Back pain** |  |  |  |  |  |
| No | 144,044 (80.4%) | 204,508 (78.1%) | 13,229 (70.8%) | 482 (63.6%) | 8,192 (67.3%) |
| Yes | 34,734 (19.4%) | 57,183 (21.9%) | 5,447 (29.2%) | 231 (30.5%) | 3,597 (29.5%) |
| Missing | 337 (0.2%) | 0 (0%) | 0 (0%) | 45 (5.9%) | 385 (3.2%) |
| **Arthritis** |  |  |  |  |  |
| No | 167,983 (93.8%) | 244,069 (93.3%) | 14,939 (80.0%) | 621 (81.9%) | 10,777 (88.5%) |
| Yes | 10,795 (6.0%) | 17,622 (6.7%) | 3,737 (20.0%) | 92 (12.1%) | 1,012 (8.3%) |
| Missing | 337 (0.2%) | 0 (0%) | 0 (0%) | 45 (5.9%) | 385 (3.2%) |
| **Petechiae** |  |  |  |  |  |
| No | 172,432 (96.3%) | 247,914 (94.7%) | 17,069 (91.4%) | 604 (79.7%) | 11,529 (94.7%) |
| Yes | 6,346 (3.5%) | 13,777 (5.3%) | 1,607 (8.6%) | 109 (14.4%) | 260 (2.1%) |
| Missing | 337 (0.2%) | 0 (0%) | 0 (0%) | 45 (5.9%) | 385 (3.2%) |
| **Tourniquet test** |  |  |  |  |  |
| No | 177,548 (99.1%) | 257,288 (98.3%) | 18,376 (98.4%) | 696 (91.8%) | 11,734 (96.4%) |
| Yes | 1,230 (0.7%) | 4,403 (1.7%) | 300 (1.6%) | 17 (2.2%) | 55 (0.5%) |
| Missing | 337 (0.2%) | 0 (0%) | 0 (0%) | 45 (5.9%) | 385 (3.2%) |
| **Myalgia** |  |  |  |  |  |
| No | 61,947 (34.6%) | 58,114 (22.2%) | 6,011 (32.2%) | 137 (18.1%) | 2,887 (23.7%) |
| Yes | 116,831 (65.2%) | 203,577 (77.8%) | 12,665 (67.8%) | 576 (76.0%) | 8,902 (73.1%) |
| Missing | 337 (0.2%) | 0 (0%) | 0 (0%) | 45 (5.9%) | 385 (3.2%) |
| **Rash** |  |  |  |  |  |
| No | 173,255 (96.7%) | 248,239 (94.9%) | 16,728 (89.6%) | 625 (82.5%) | 11,533 (94.7%) |
| Yes | 5,523 (3.1%) | 13,452 (5.1%) | 1,948 (10.4%) | 88 (11.6%) | 256 (2.1%) |
| Missing | 337 (0.2%) | 0 (0%) | 0 (0%) | 45 (5.9%) | 385 (3.2%) |
| **Nausea** |  |  |  |  |  |
| No | 120,285 (67.2%) | 166,011 (63.4%) | 13,019 (69.7%) | 384 (50.7%) | 7,333 (60.2%) |
| Yes | 58,493 (32.7%) | 95,680 (36.6%) | 5,657 (30.3%) | 329 (43.4%) | 4,456 (36.6%) |
| Missing | 337 (0.2%) | 0 (0%) | 0 (0%) | 45 (5.9%) | 385 (3.2%) |
| **Conjunctivitis** |  |  |  |  |  |
| No | 177,526 (99.1%) | 258,559 (98.8%) | 18,260 (97.8%) | 701 (92.5%) | 11,709 (96.2%) |
| Yes | 1,252 (0.7%) | 3,132 (1.2%) | 416 (2.2%) | 12 (1.6%) | 80 (0.7%) |
| Missing | 337 (0.2%) | 0 (0%) | 0 (0%) | 45 (5.9%) | 385 (3.2%) |
| **Intense arthralgia** |  |  |  |  |  |
| No | 150,650 (84.1%) | 203,175 (77.6%) | 8,311 (44.5%) | 527 (69.5%) | 9,833 (80.8%) |
| Yes | 28,128 (15.7%) | 58,516 (22.4%) | 10,365 (55.5%) | 186 (24.5%) | 1,956 (16.1%) |
| Missing | 337 (0.2%) | 0 (0%) | 0 (0%) | 45 (5.9%) | 385 (3.2%) |
| **Leukopenia** |  |  |  |  |  |
| No | 176,809 (98.7%) | 255,599 (97.7%) | 18,348 (98.2%) | 686 (90.5%) | 11,603 (95.3%) |
| Yes | 1,969 (1.1%) | 6,092 (2.3%) | 328 (1.8%) | 27 (3.6%) | 186 (1.5%) |
| Missing | 337 (0.2%) | 0 (0%) | 0 (0%) | 45 (5.9%) | 385 (3.2%) |
| **Retro-orbital pain** |  |  |  |  |  |
| No | 134,804 (75.3%) | 171,429 (65.5%) | 13,328 (71.4%) | 416 (54.9%) | 7,747 (63.6%) |
| Yes | 43,974 (24.6%) | 90,262 (34.5%) | 5,348 (28.6%) | 297 (39.2%) | 4,042 (33.2%) |
| Missing | 337 (0.2%) | 0 (0%) | 0 (0%) | 45 (5.9%) | 385 (3.2%) |
| **Diabetes** |  |  |  |  |  |
| No | 174,457 (97.4%) | 253,417 (96.8%) | 17,308 (92.7%) | 688 (90.8%) | 11,337 (93.1%) |
| Yes | 4,049 (2.3%) | 8,043 (3.1%) | 1,283 (6.9%) | 25 (3.3%) | 450 (3.7%) |
| Ignored | 272 (0.2%) | 231 (0.1%) | 85 (0.5%) | 0 (0%) | 2 (0.0%) |
| Missing | 337 (0.2%) | 0 (0%) | 0 (0%) | 45 (5.9%) | 385 (3.2%) |
| **Hepatopathies** |  |  |  |  |  |
| No | 178,349 (99.6%) | 261,119 (99.8%) | 18,539 (99.3%) | 712 (93.9%) | 11,771 (96.7%) |
| Yes | 159 (0.1%) | 343 (0.1%) | 45 (0.2%) | 1 (0.1%) | 16 (0.1%) |
| Ignored | 270 (0.2%) | 229 (0.1%) | 92 (0.5%) | 0 (0%) | 2 (0.0%) |
| Missing | 337 (0.2%) | 0 (0%) | 0 (0%) | 45 (5.9%) | 385 (3.2%) |
| **Arterial hypertension** |  |  |  |  |  |
| No | 167,372 (93.4%) | 241,038 (92.1%) | 15,678 (83.9%) | 622 (82.1%) | 10,546 (86.6%) |
| Yes | 11,136 (6.2%) | 20,423 (7.8%) | 2,915 (15.6%) | 91 (12.0%) | 1,241 (10.2%) |
| Ignored | 270 (0.2%) | 230 (0.1%) | 83 (0.4%) | 0 (0%) | 2 (0.0%) |
| Missing | 337 (0.2%) | 0 (0%) | 0 (0%) | 45 (5.9%) | 385 (3.2%) |
| **Autoimmune diseases** |  |  |  |  |  |
| No | 178,046 (99.4%) | 260,568 (99.6%) | 18,429 (98.7%) | 710 (93.7%) | 11,760 (96.6%) |
| Yes | 464 (0.3%) | 884 (0.3%) | 150 (0.8%) | 3 (0.4%) | 26 (0.2%) |
| Ignored | 268 (0.1%) | 239 (0.1%) | 97 (0.5%) | 0 (0%) | 3 (0.0%) |
| Missing | 337 (0.2%) | 0 (0%) | 0 (0%) | 45 (5.9%) | 385 (3.2%) |
| **Hematological diseases** |  |  |  |  |  |
| No | 178,380 (99.6%) | 261,182 (99.8%) | 18,548 (99.3%) | 713 (94.1%) | 11,778 (96.7%) |
| Yes | 136 (0.1%) | 285 (0.1%) | 36 (0.2%) | 0 (0%) | 9 (0.1%) |
| Ignored | 262 (0.1%) | 224 (0.1%) | 92 (0.5%) | 0 (0%) | 2 (0.0%) |
| Missing | 337 (0.2%) | 0 (0%) | 0 (0%) | 45 (5.9%) | 385 (3.2%) |
| **Chronic kidney disease** |  |  |  |  |  |
| No | 178,258 (99.5%) | 261,133 (99.8%) | 18,538 (99.3%) | 712 (93.9%) | 11,778 (96.7%) |
| Yes | 260 (0.1%) | 333 (0.1%) | 46 (0.2%) | 1 (0.1%) | 9 (0.1%) |
| Ignored | 260 (0.1%) | 225 (0.1%) | 92 (0.5%) | 0 (0%) | 2 (0.0%) |
| Missing | 337 (0.2%) | 0 (0%) | 0 (0%) | 45 (5.9%) | 385 (3.2%) |
| **Peptic acid disease** |  |  |  |  |  |
| No | 178,466 (99.6%) | 261,272 (99.8%) | 18,554 (99.3%) | 713 (94.1%) | 11,784 (96.8%) |
| Yes | 48 (0.0%) | 200 (0.1%) | 30 (0.2%) | 0 (0%) | 3 (0.0%) |
| Ignored | 264 (0.1%) | 219 (0.1%) | 92 (0.5%) | 0 (0%) | 2 (0.0%) |
| Missing | 337 (0.2%) | 0 (0%) | 0 (0%) | 45 (5.9%) | 385 (3.2%) |
| **Hospitalization** |  |  |  |  |  |
| No | 175,739 (98.1%) | 255,689 (97.7%) | 18,208 (97.5%) | 685 (90.4%) | 11,717 (96.2%) |
| Yes | 2,995 (1.7%) | 5,945 (2.3%) | 460 (2.5%) | 27 (3.6%) | 72 (0.6%) |
| Ignored | 32 (0.0%) | 49 (0.0%) | 6 (0.0%) | 1 (0.1%) | 0 (0%) |
| Missing | 349 (0.2%) | 8 (0.0%) | 2 (0.0%) | 45 (5.9%) | 385 (3.2%) |

IQR = interquartile range; SD = Standard deviation.

All attempts to develop Zika models returned models with unacceptable performance. From the 28 initially considered as potential predictors for dengue fever, the following 24 were selected (in order of importance): fever, first symptom week, home address, myalgia, race, education, headache, retro-orbital pain, age at notification, petechiae, rash/exanthema, leukopenia, back pain, intense arthralgia, arthritis, sex at notification, tourniquet test, arterial hypertension, nausea, vomiting, diabetes, conjunctivitis, pregnancy, and chronic kidney disease (Figure 2).

**Figure 2:**
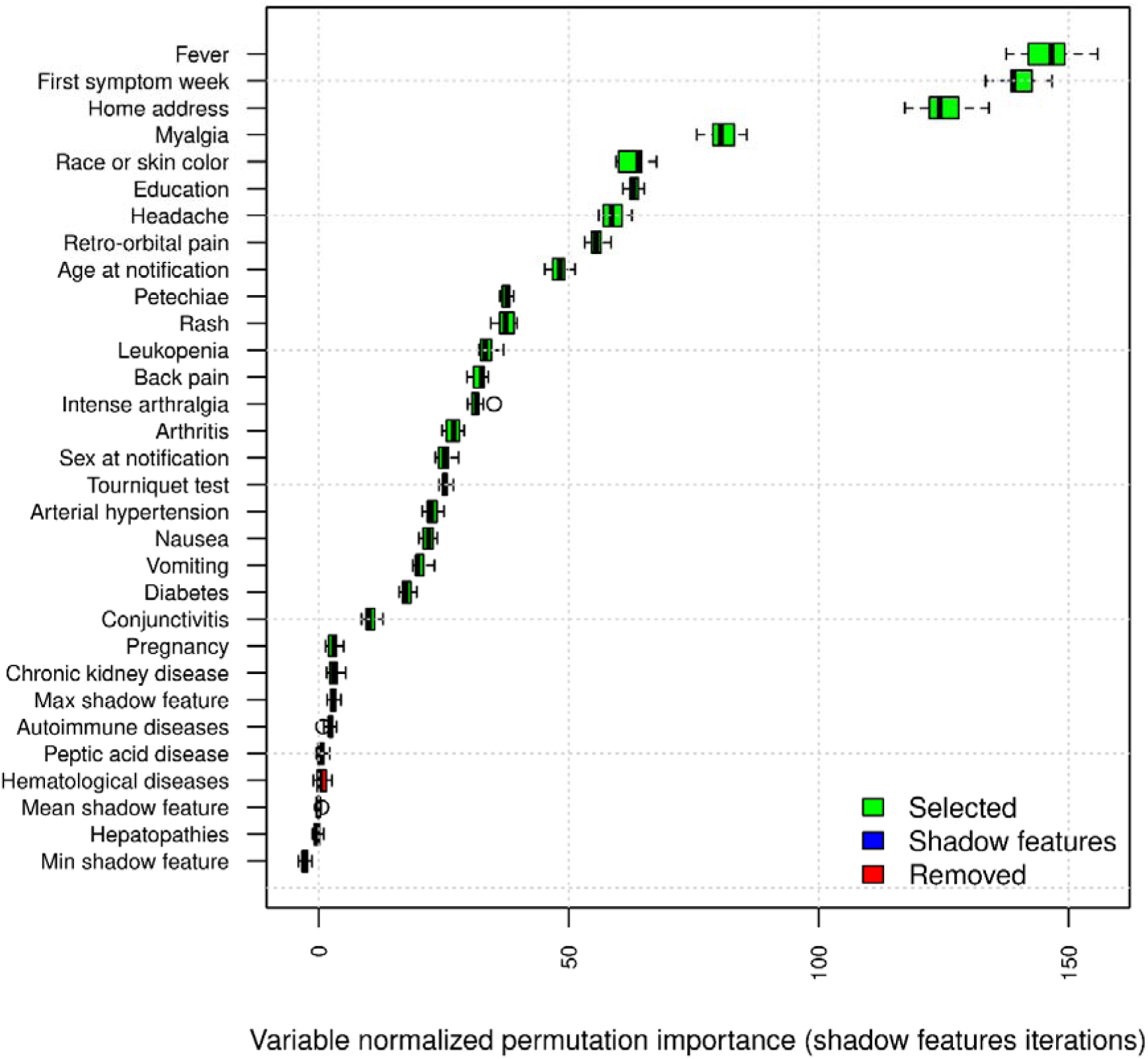
Predictors normalized permutations importance compared to shadow features for random forest regression predicting dengue.

One may see that the seasonality is not very well defined in the model, although cases in late summer are more likely (epidemiological weeks between 5 and 15), and age has two spikes, one in childhood and one in old age. Sex, pregnancy, vomiting, back pain, nausea, conjunctivitis, intense arthralgia, diabetes, arterial hypertension, and chronic kidney disease have very little mean effect. People with white skin are less likely to have dengue fever, as well as living in a rural area, having less schooling, having arthritis, and having chronic kidney disease. The remaining predictors, when present, increase the mean probability of having Dengue as expected (Figure 3). The most relevant features in discriminating from other arboviruses were first symptom week, age, race, headache, vomiting, petechiae, tourniquet test, myalgia, retro-orbital pain, and leukopenia (Table 2).

**Figure 3:**
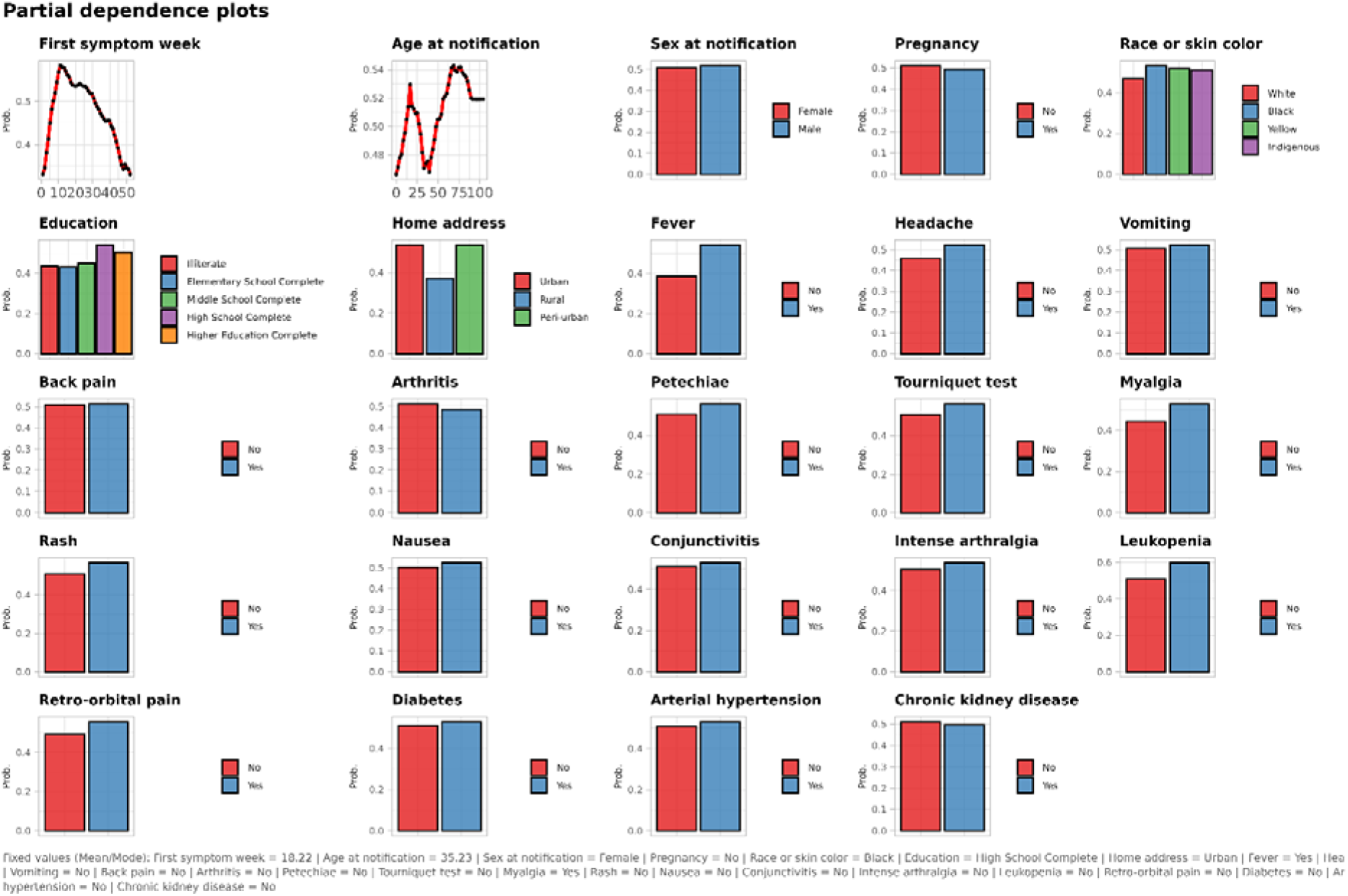
Partial dependency marginal effects for the retained predictors for random forest regression predicting dengue.

**Table 2:**
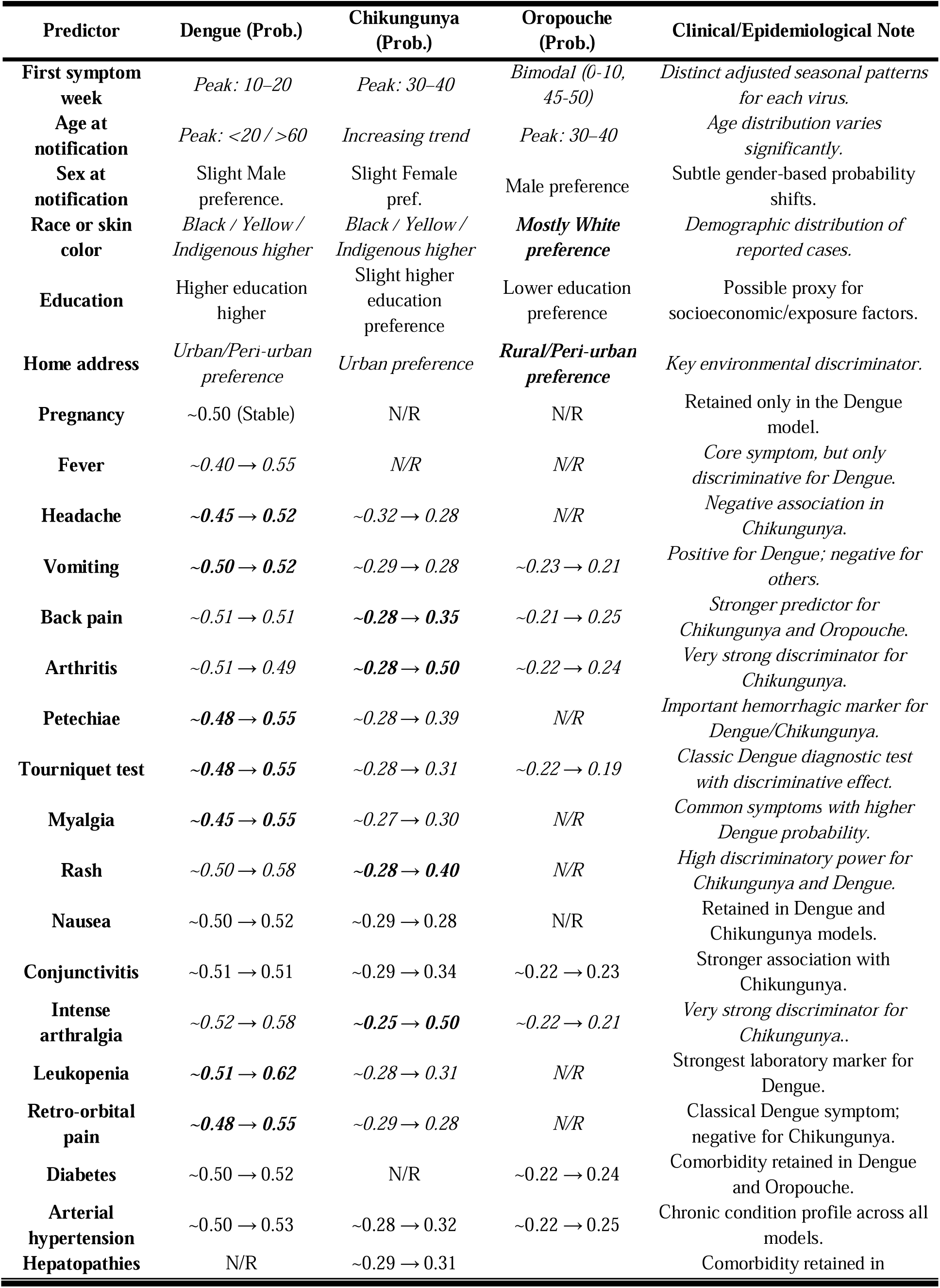
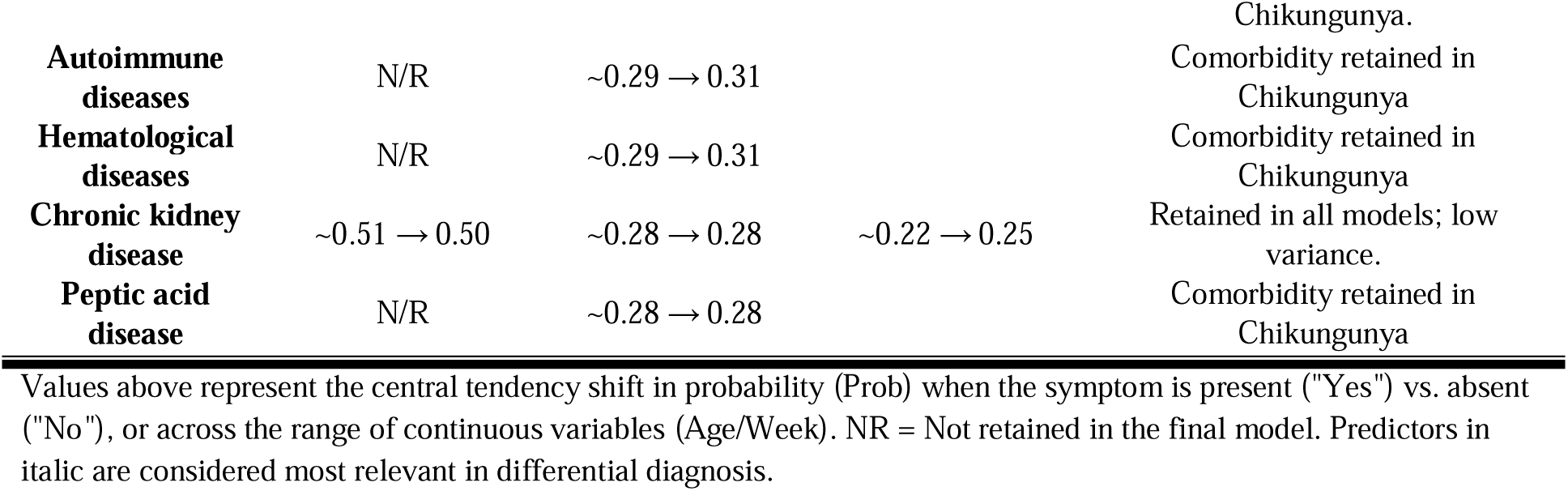
Summary comparative discriminative profile of the predictors from the random forest regression models based on the partial effect analysis.

From the 28 initially considered as potential predictors for chikungunya, the following 25 were selected (in order of importance): intense arthralgia, arthritis, petechiae, sex at notification, education, age at notification, retro-orbital pain, back pain, myalgia, headache, skin color, leukopenia, home address, tourniquet test, rash/exanthema, conjunctivitis, autoimmune diseases, nausea, first week symptoms, hepatopathies, arterial hypertension, vomiting, peptic acid disease, hematological disease, and chronic kidney disease (Figure 4).

**Figure 4.**
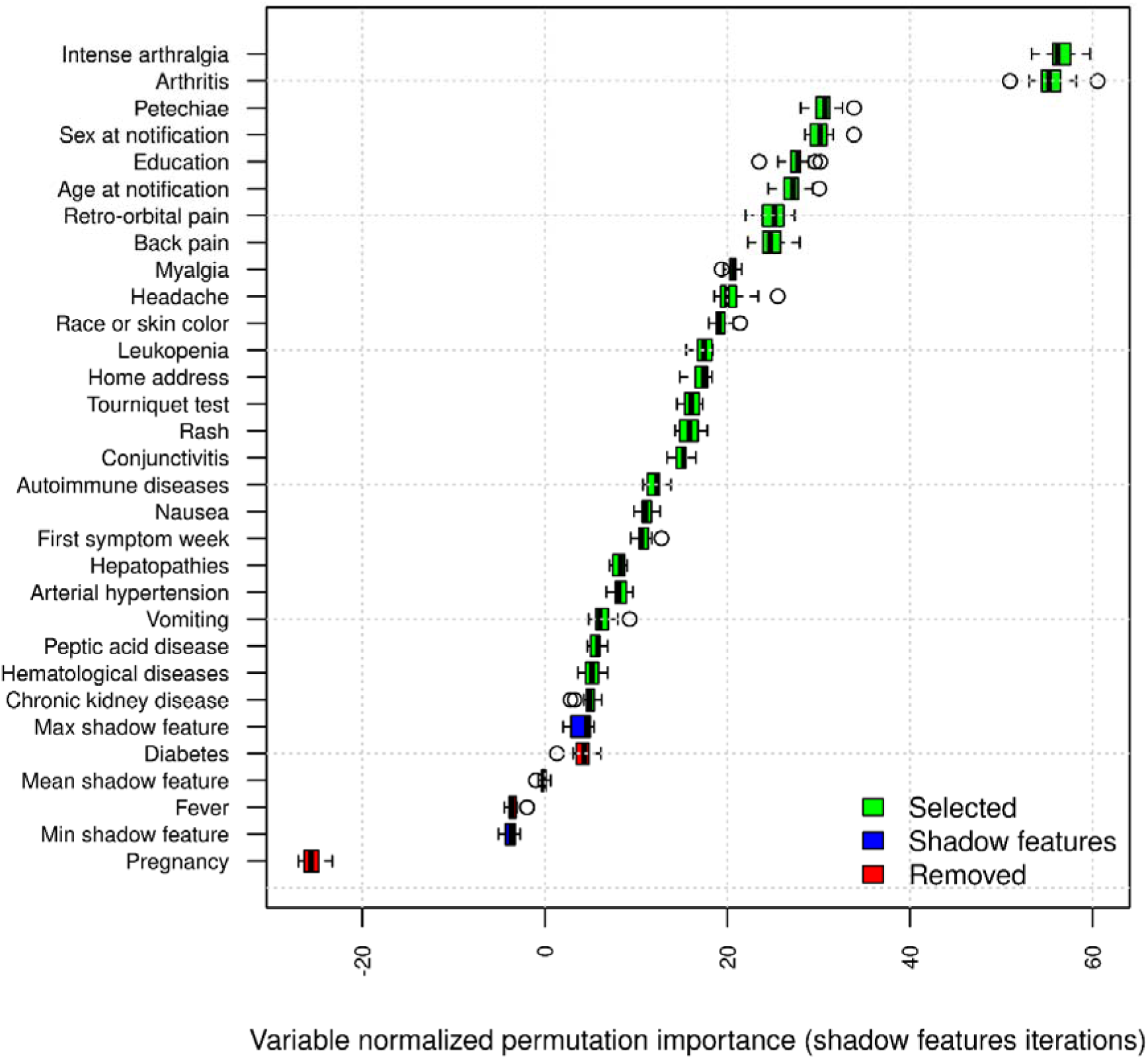
Predictors normalized permutations importance compared to shadow features for random forest regression predicting chikungunya.

One may see that Chikungunya is more likely in the winter period (between weeks 20 and 40). Also, the age probability is irregular, but it is more likely to have chikungunya at 35 or later. Chikungunya is less likely in males, subjects with white skin, and those living in rural areas.

Education, vomiting, nausea, leukopenia, retro-orbital pain, hepatopathies, hematological diseases, chronic kidney disease, and peptic acid disease have very little effect. The remaining predictors, when present, increase the probability of chikungunya diagnosis, except headache (Figure 5). The most relevant features in discriminating from other arboviruses were first symptom week, age, race, back pain, intense arthralgia, arthritis, and rash. One may notice that the chikungunya model also included a greater number of comorbidities (Table 2).

**Figure 5:**
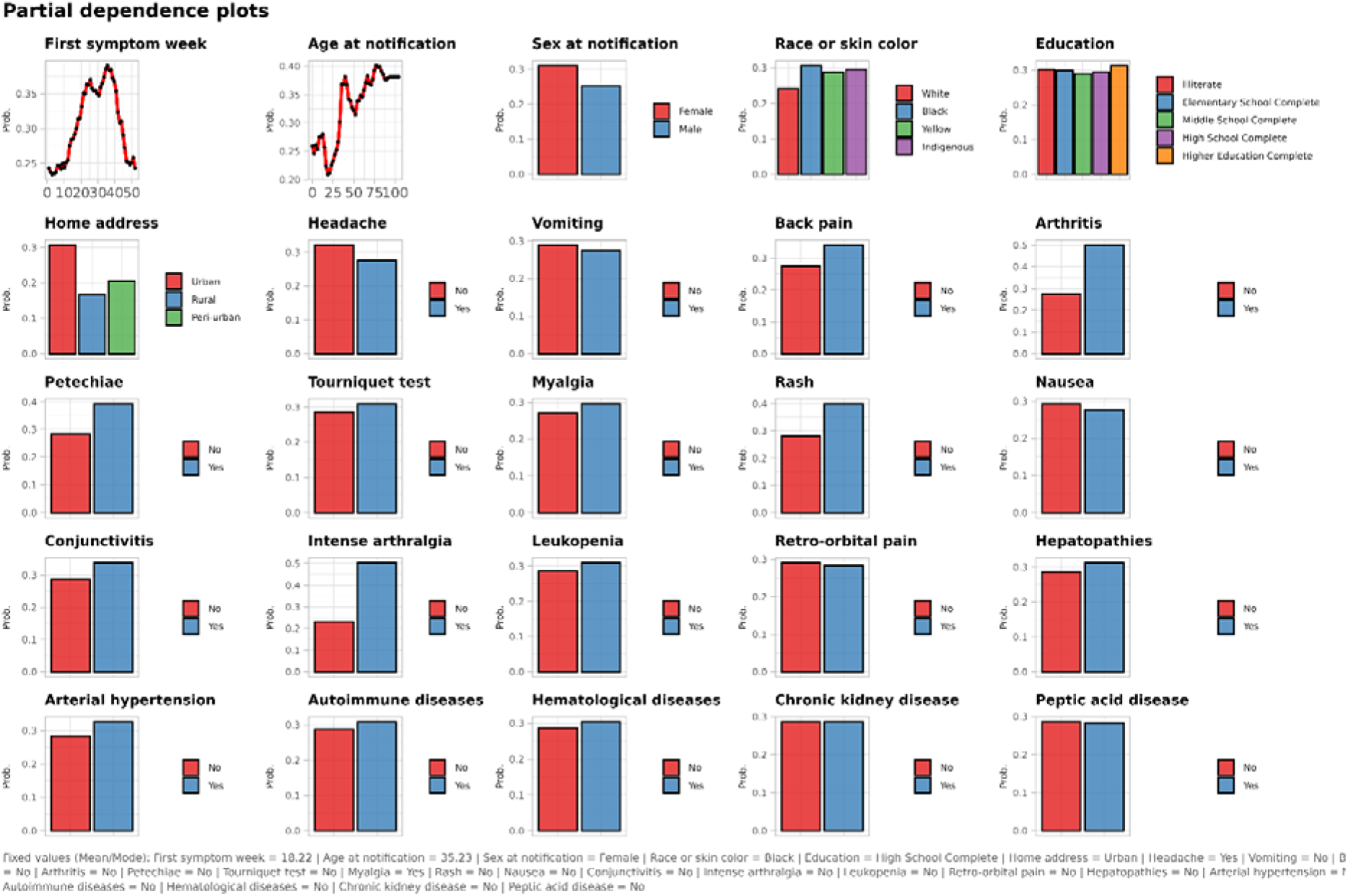
Partial dependency marginal effects for the retained predictors for random forest regression predicting chikungunya.

From the 28 initially considered as potential predictors for OF, the following 16 were selected (in order of importance): weeks of first symptoms, age, education, home address, skin color, arterial hypertension, intense arthralgia, back pain, diabetes, conjunctivitis, sex, arthritis, vomiting, chronic kidney diseases, tourniquet test, and autoimmune diseases (Figure 6) One may see that OF becomes more likely in the early summer (between weeks 45 and 5), and it is increasingly more likely as age increases, and its probability stabilizes after age 25. OF is more likely among subjects with white skin color, with lower education, and living in a rural area. Back pain and arthritis increase the OF probability, and the comorbidities such as diabetes, arterial hypertension, autoimmune diseases, and chronic kidney diseases also increase the likelihood. The remaining predictors either have very little effect or a negative effect, such as vomiting (Figure 7). For discriminatory rationale, only the epidemiological predictors were relevant for OF (Table 2)

**Figure 6.**
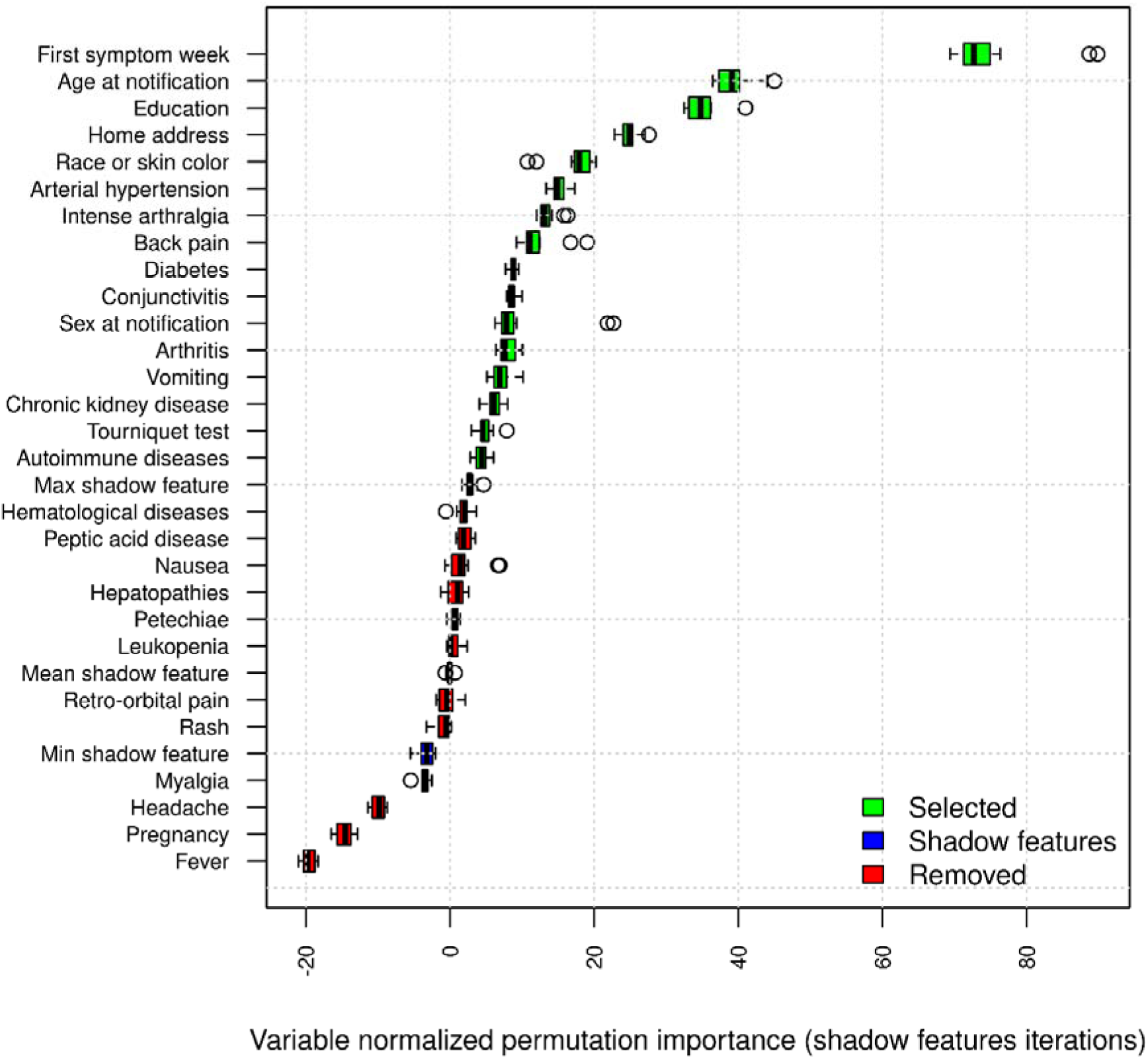
Predictors normalized permutations importance compared to shadow features for random forest regression predicting oropouche.

**Figure 7:**
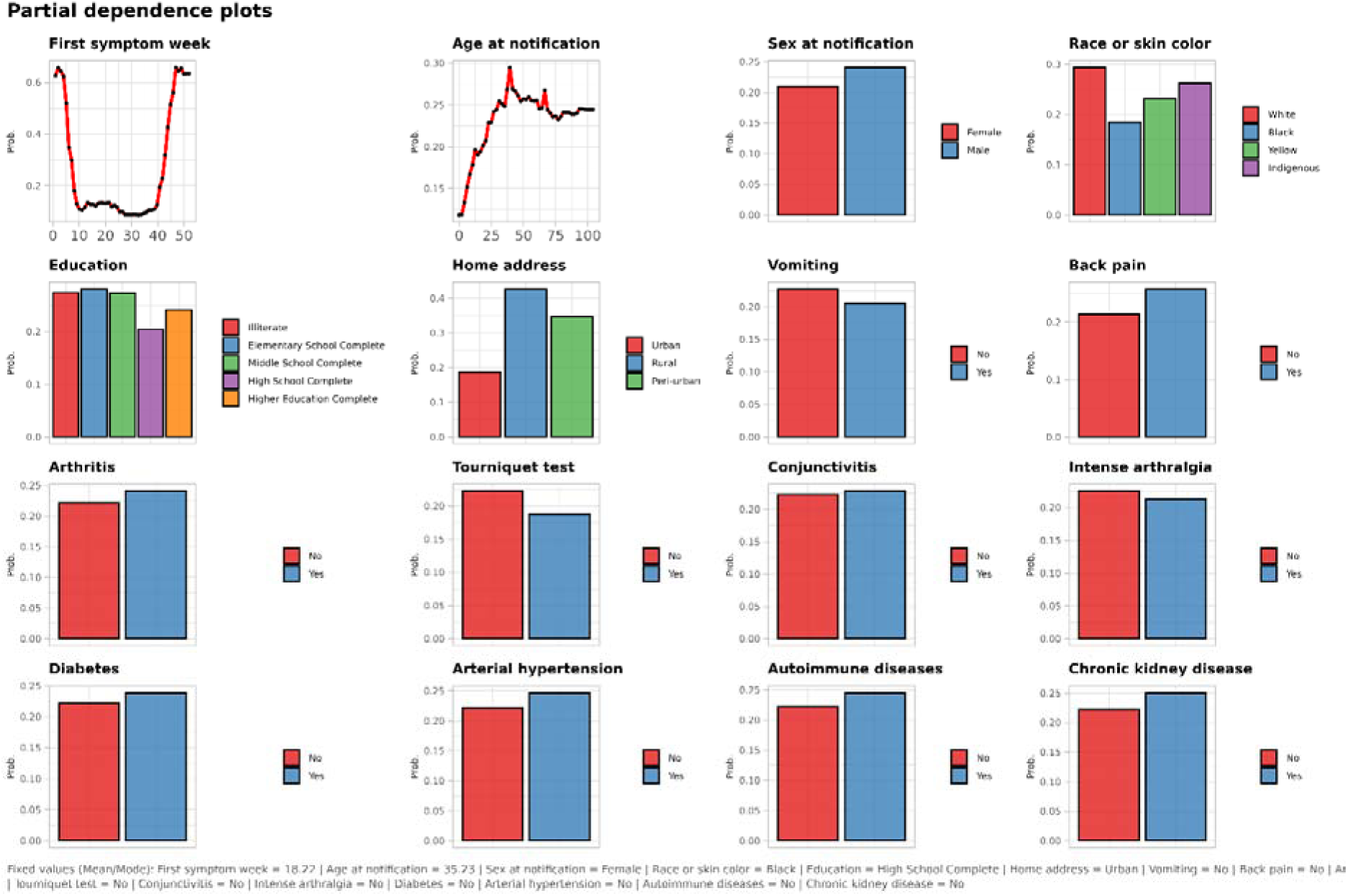
Partial dependency marginal effects for the retained predictors for random forest regression predicting oropouche.

The original models trained for chikungunya and OF had unusual performance metrics, most likely due to very skewed frequencies of the outcome. All three models had good discrimination and moderate to good calibration after scaling predictions. The dengue model was the least discriminative among all three, but with the most reasonable calibration metrics. On the other hand, the OF model was the most discriminative but with less optimized calibration metrics. (Table 3) The calibration belt, checking the correspondence between observed and predicted values, must be interpreted carefully, as the amount of data makes the belt narrow. Nevertheless, for the dengue model, it has very good correspondence for both training and testing data. On the other hand, for both OF and chikungunya models, the predictions up to 0.25 (where most predictions are) are reasonable, but after that, the models tend to underestimate the observed values (Figure 8).

**Figure 8.**
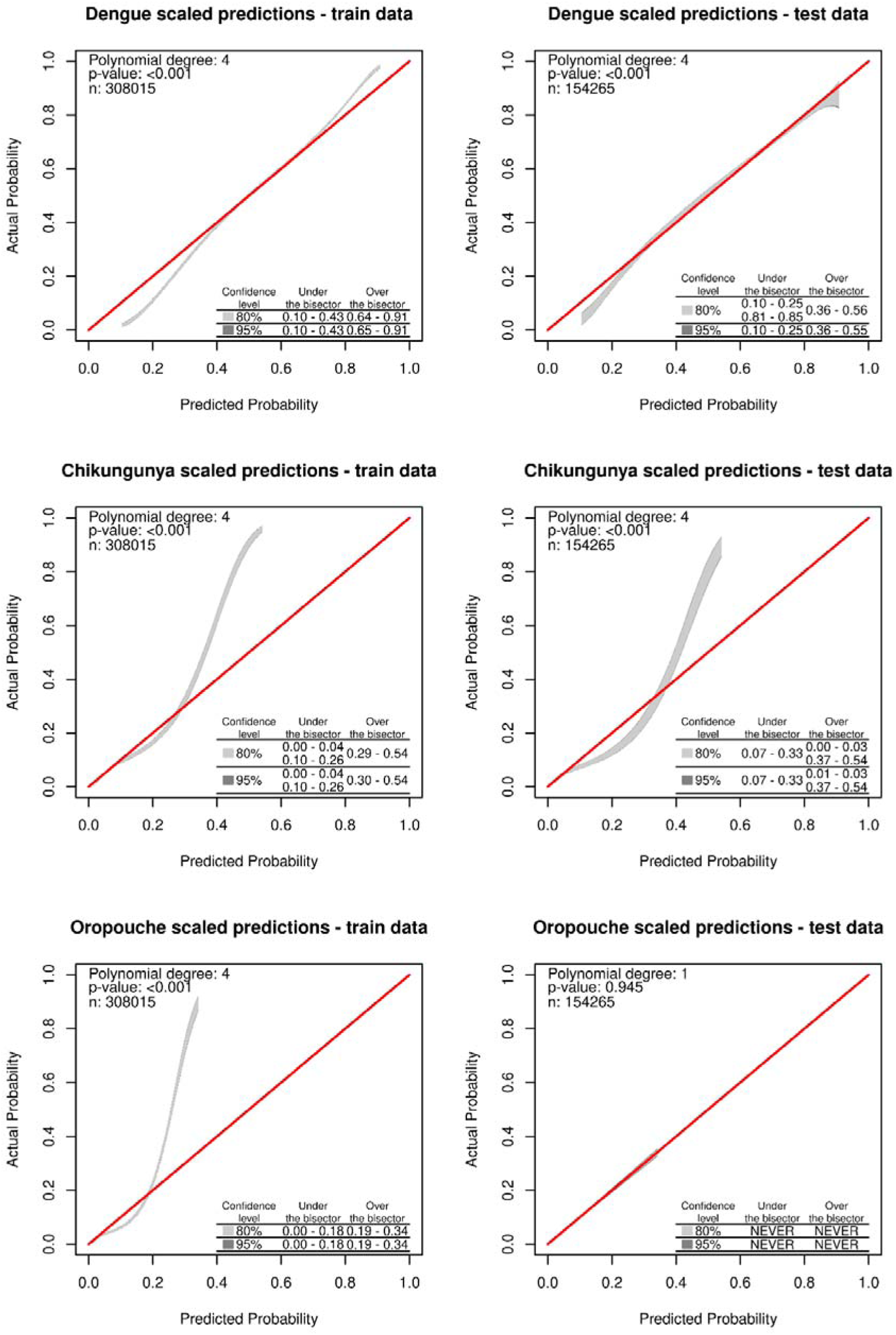
Calibration belt plots for regression random forest models predicting dengue, chikungunya, and oropouchee in the train and test datasets.

**Table 3-.**
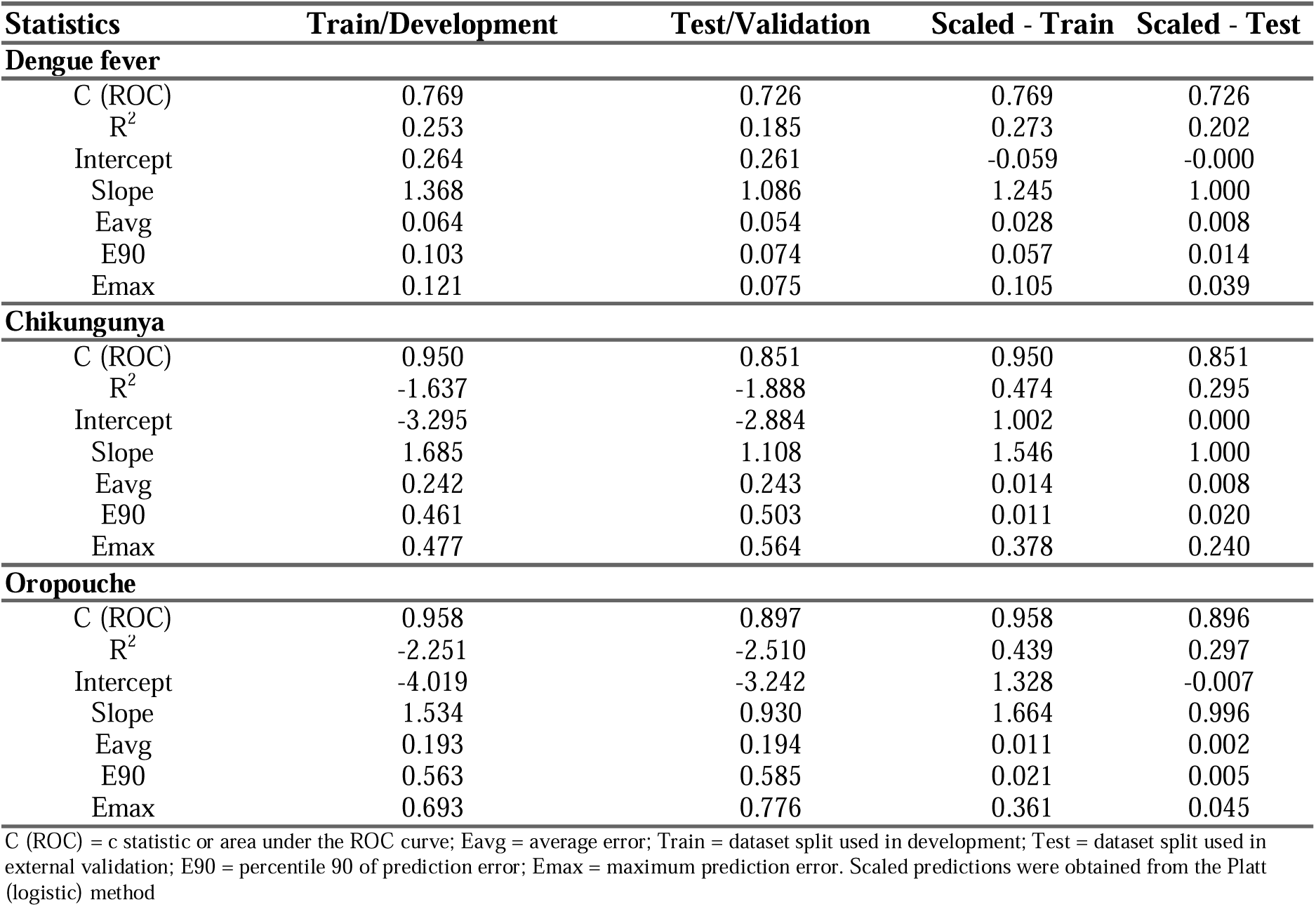
Random forest regression selected performance measures estimated from the predicted probabilities, scaled predicted probabilities of dengue fever, chikungunya, and oropouche on different datasets.

For the dengue model, the mixed density distributions in both datasets (training and testing) reveal a great overlap with an intersection close to the midpoint. In this situation, both the uncertain interval and the inconclusive interval, as well as the density intersection and the threshold where sensitivity equals specificity, are very similar. An inconclusive range between 0.47 and 0.67 estimated with the test data is recommended (Figure 9). For the chikungunya model, the predictions were skewed to the left, and the predictions did not reach the whole range from 0 to 1. Although the uncertain interval and the inconclusive range are narrower than in the dengue analysis, they seem to agree less with each other in the test and train data. The estimated and recommended inconclusive limits from the test set are 0.025 and 0.037, and the threshold at which sensitivity equals specificity is 0.031 (Figure 10).

**Figure 9:**
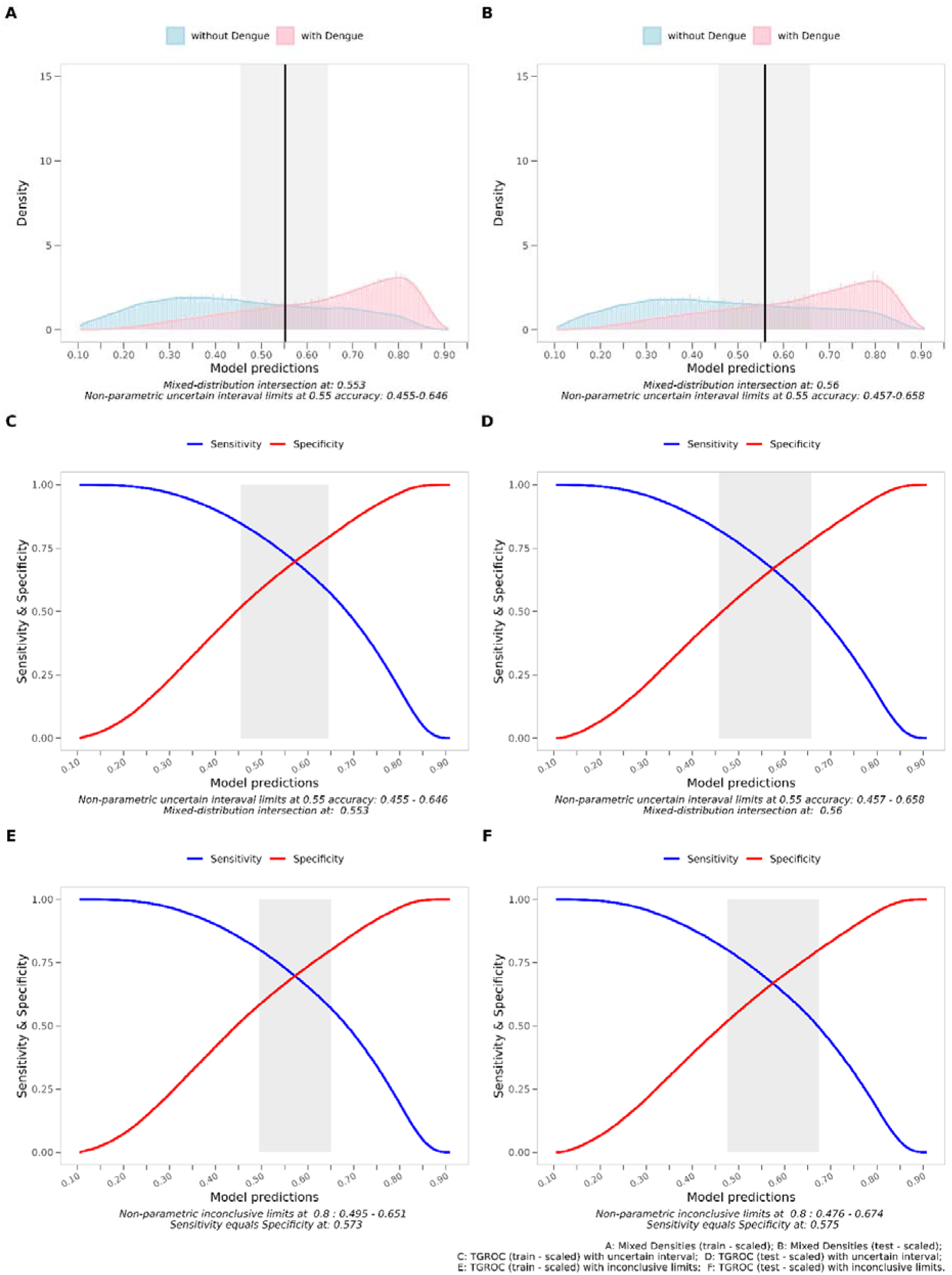
Random forest regression mixed densities, sensitivity, specificity trade-off, and gray intervals for dengue on different datasets.

**Figure 10:**
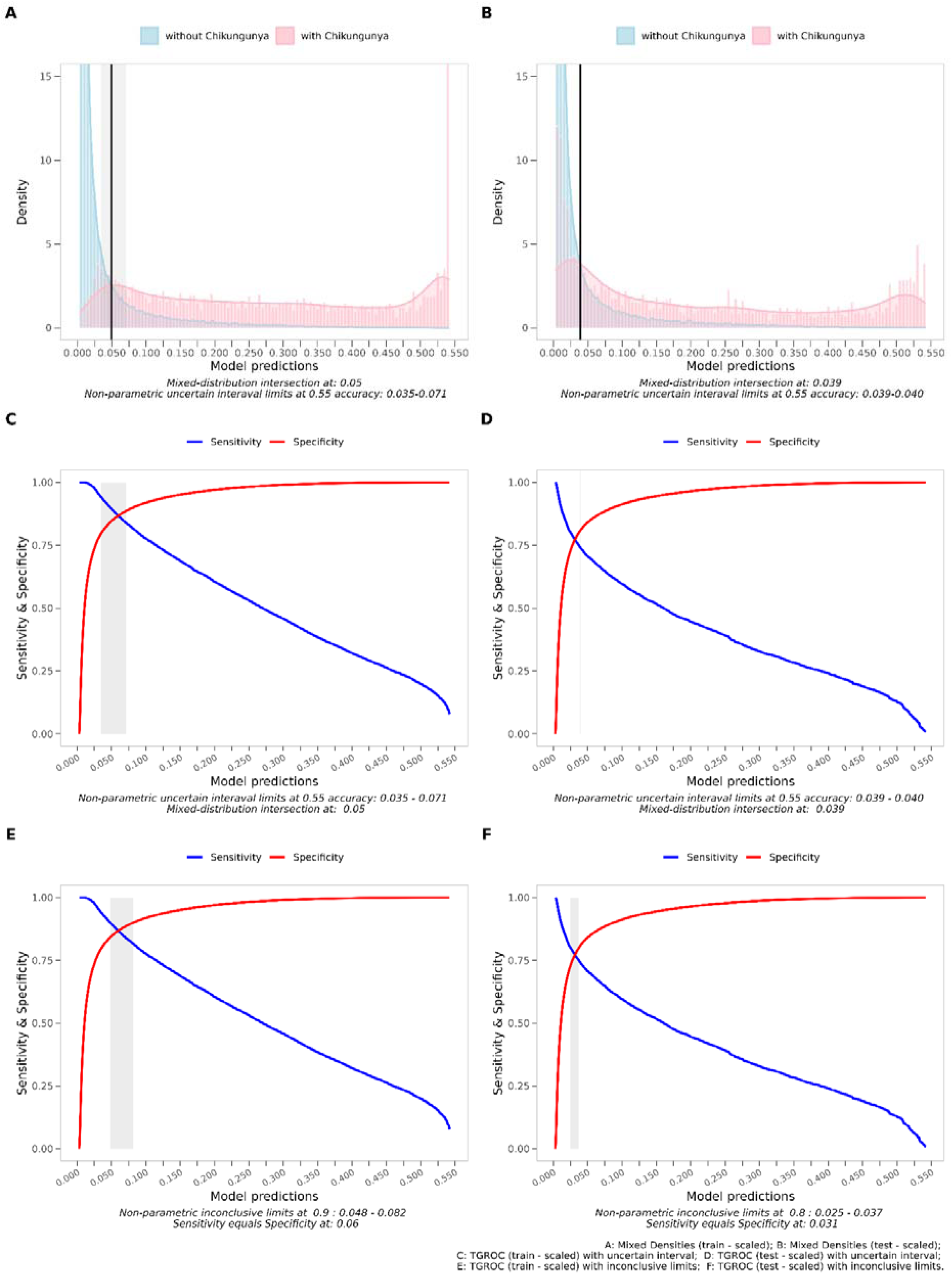
Random forest regression mixed densities, sensitivity, specificity trade-off, and gray intervals for chikungunya on different datasets.

Similarly, the OF model had skewed predictions to the left and did not reach the whole possible range with similar density overlaps, and narrower uncertain and inconclusive ranges. For the OF model, the inconclusive limit estimate with the test set was 0.025 and 0.040, with a threshold at which sensitivity equals specificity of 0.03 (Figure 11). Some of the analyses have inconclusive limits, with a threshold set higher than 0.8 for sensitivity and specificity. In these cases, setting with 0.8 would lead to the absence of inconclusive limits, or a higher limit lower than the lower limit, which makes no sense.

**Figure 11:**
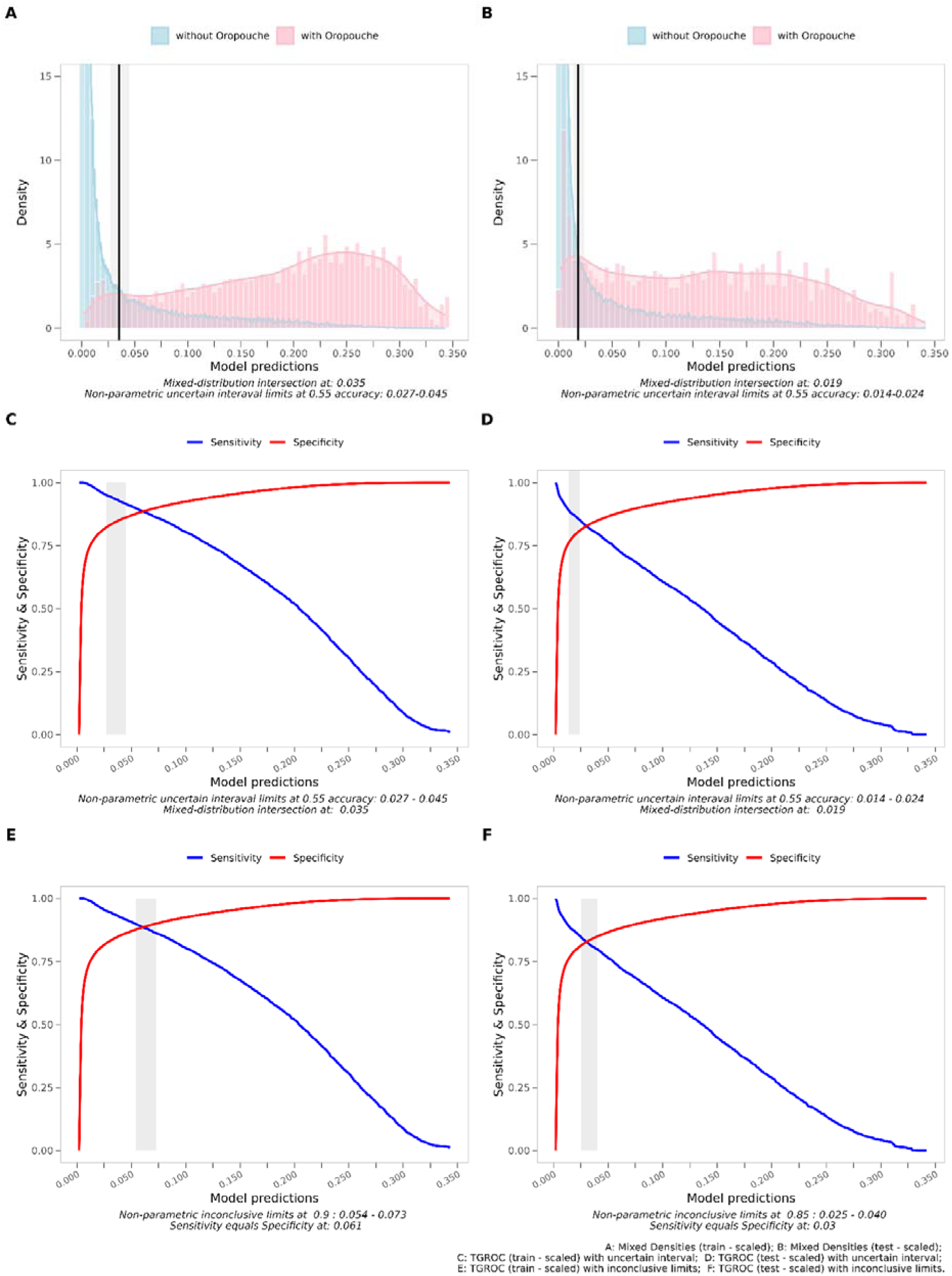
Random forest regression mixed densities, sensitivity, specificity trade-off, and gray intervals for oropouche on different datasets

One may understand that each prediction is associated with a pair of sensitivity and specificity, and by deciding on different thresholds, these metrics change accordingly. This may change the course of action, particularly for the Dengue model, which is less accurate with sparser, overlapping prediction densities. Nevertheless, we estimate the model performance as a classification (with and without dengue) by picking the threshold where sensitivity equals specificity in the test dataset. It returns a similar interpretation from the previous results, with an inherent loss of performance due to the dichotomization (Table 4). When examining the same analysis for the Chikungunya (Table 5) and OF models (Table 6), it is possible to have similar interpretations when compared to the previous performance analysis. However, if the classification is goal, here the sensitivity and specificity trade-off will be way less evident when picking different thresholds, as the inconclusive limits are narrower.

**Table 4:** Regression random forest performance measures for dengue fever with scaled predictions: Training vs. Validation.

| Statistics | Training dataset |  |  | Test dataset |  |  |
| --- | --- | --- | --- | --- | --- | --- |
|  | Estimate | 95% lower cl | 95% upper cl | Estimate | 95% lower cl | 95% upper cl |
| Sample size: | 308,015 | --- | --- | 154,265 | --- | --- |
| Prevalence: | 0.566 | 0.564 | 0.568 | 0.566 | 0.564 | 0.569 |
| Sensitivity: | 0.693 | 0.691 | 0.695 | 0.667 | 0.664 | 0.670 |
| Specificity: | 0.701 | 0.698 | 0.703 | 0.669 | 0.665 | 0.672 |
| Positive predictive value: | 0.751 | 0.749 | 0.753 | 0.724 | 0.721 | 0.727 |
| Negative predictive value: | 0.636 | 0.634 | 0.639 | 0.606 | 0.603 | 0.610 |
| Positive likelihood ratio: | 2.315 | 2.294 | 2.335 | 2.014 | 1.990 | 2.038 |
| Negative likelihood ratio: | 0.438 | 0.435 | 0.442 | 0.498 | 0.492 | 0.503 |
| Diagnostic Odds Ratio: | 5.279 | 5.197 | 5.362 | 4.047 | 3.961 | 4.134 |
| Error rate: | 0.304 | 0.302 | 0.305 | 0.332 | 0.330 | 0.335 |
| Accuracy: | 0.696 | 0.695 | 0.698 | 0.668 | 0.665 | 0.670 |
| Youden J index: | 0.393 | 0.390 | 0.397 | 0.336 | 0.331 | 0.341 |
| Area under ROC curve: | 0.697 | --- | --- | 0.668 | --- | --- |

lower cl = lower confidence limit; upper cl = upper confidence limit; Na = not assigned; Inf = Infinite. Decision limit set as 0.575 (from the sensitivity equals specificity from the test dataset)

**Table 5:** Regression random forest performance measures for Chikungunya fever with scaled predictions: Training vs. Validation.

| Statistics | Training dataset |  |  | Test dataset |  |  |
| --- | --- | --- | --- | --- | --- | --- |
|  | Estimate | 95% lower cl | 95% upper cl | Estimate | 95% lower cl | 95% upper cl |
| Sample size: | 308,015 | --- | --- | 154,265 | --- | --- |
| Prevalence: | 0.040 | 0.040 | 0.041 | 0.040 | 0.039 | 0.041 |
| Sensitivity: | 0.955 | 0.952 | 0.959 | 0.775 | 0.764 | 0.785 |
| Specificity: | 0.777 | 0.775 | 0.778 | 0.770 | 0.768 | 0.772 |
| Positive predictive value: | 0.153 | 0.150 | 0.155 | 0.124 | 0.121 | 0.127 |
| Negative predictive value: | 0.998 | 0.997 | 0.998 | 0.988 | 0.987 | 0.988 |
| Positive likelihood ratio: | 4.276 | 4.242 | 4.310 | 3.372 | 3.317 | 3.427 |
| Negative likelihood ratio: | 0.058 | 0.053 | 0.062 | 0.293 | 0.280 | 0.307 |
| Diagnostic Odds Ratio: | 74.328 | 68.253 | 80.958 | 11.516 | 10.835 | 12.247 |
| Error rate: | 0.216 | 0.215 | 0.218 | 0.230 | 0.227 | 0.232 |
| Accuracy: | 0.784 | 0.782 | 0.785 | 0.770 | 0.768 | 0.773 |
| Youden J index: | 0.732 | 0.728 | 0.736 | 0.545 | 0.534 | 0.555 |
| Area under ROC curve: | 0.866 | --- | --- | 0.772 | --- | --- |

lower cl = lower confidence limit; upper cl = upper confidence limit; Na = not assigned; Inf = Infinite. Decision limit set as 0.031 (from the sensitivity equals specificity from the test dataset)

**Table 6:** Regression random forest performance measures for Oropouche fever with scaled predictions: Training vs. Validation.

| Statistics | Training dataset |  |  | Test dataset |  |  |
| --- | --- | --- | --- | --- | --- | --- |
|  | Estimate | 95% lower cl | 95% upper cl | Estimate | 95% lower cl | 95% upper cl |
| Sample size: | 308,015 | --- | --- | 154,265 | --- | --- |
| Prevalence: | 0.026 | 0.026 | 0.027 | 0.026 | 0.025 | 0.027 |
| Sensitivity: | 0.946 | 0.941 | 0.951 | 0.830 | 0.818 | 0.841 |
| Specificity: | 0.831 | 0.830 | 0.832 | 0.827 | 0.826 | 0.829 |
| Positive predictive value: | 0.132 | 0.129 | 0.134 | 0.115 | 0.111 | 0.118 |
| Negative predictive value: | 0.998 | 0.998 | 0.998 | 0.994 | 0.994 | 0.995 |
| Positive likelihood ratio: | 5.594 | 5.540 | 5.649 | 4.808 | 4.723 | 4.894 |
| Negative likelihood ratio: | 0.065 | 0.059 | 0.071 | 0.206 | 0.192 | 0.220 |
| Diagnostic Odds Ratio: | 86.255 | 78.346 | 95.073 | 23.358 | 21.487 | 25.406 |
| Error rate: | 0.166 | 0.165 | 0.167 | 0.173 | 0.171 | 0.174 |
| Accuracy: | 0.834 | 0.833 | 0.835 | 0.827 | 0.826 | 0.829 |
| Youden J index: | 0.777 | 0.772 | 0.782 | 0.657 | 0.645 | 0.669 |
| Area under ROC curve: | 0.888 | --- | --- | 0.829 | --- | --- |

lower cl = lower confidence limit; upper cl = upper confidence limit; Na = not assigned; Inf = Infinite. Decision limit set as 0.030 (from the sensitivity equals specificity from the test dataset)

## DISCUSSION

The main results to be discussed are: (a) it was possible to develop and validate models for differential diagnosis of these arboviruses with good model performances; (b) although these conditions may have similar seasonality and symptoms in common, there is some preferential information that makes it possible to rank their likelihood; (c) applicability and integration in ordinary diagnostic investigation is discussed as the models are not intended to replace laboratory investigation.

From a clinical perspective, predictive models are best understood as decision-support tools rather than replacements for clinical judgment. Their applicability is particularly relevant in the context of arboviral diseases, where overlapping clinical manifestations—such as fever, myalgia, and rash—complicate differential diagnosis. By integrating clinical and laboratory variables, these models can improve probabilistic classification and reduce diagnostic uncertainty, especially in early disease stages (34,35).

Predictive models for arboviral diseases—particularly dengue, and chikungunya have shown increasing applicability in both epidemiological surveillance and clinical decision support. Evidence from systematic reviews highlights both their promising performance and their practical advantages in supporting diagnostic investigation, despite important methodological limitations (14,17,21,22,24,26,27,36–38).

Most predictive models for arboviruses rely on machine learning techniques, especially tree-based algorithms (e.g., Random Forest, Decision Trees) and Support Vector Machines. Several studies have demonstrated moderate-to-high discriminatory capacity in clinical datasets (AUC ≈ 0.80–0.90) (39,40). However, very high performance reported in some studies (e.g., accuracy >99%) is often associated with small datasets and potential overfitting, limiting external validity (34,41). For chikungunya, fewer predictive models are available, and most are developed within multi-class classification frameworks that include dengue and other arboviruses, reflecting the substantial clinical overlap between these infections (40). In contrast, predictive modeling for OF remains incipient, with most studies focusing on ecological niche modeling and outbreak prediction rather than clinical diagnostic support. This highlights a critical gap in the literature, particularly given the increasing recognition of Oropouche fever as an emerging arbovirus in Latin America (27,42,43).

The advantages of predictive models in diagnostic investigation include improved diagnostic accuracy, earlier case detection, and support for decision-making in resource-limited settings where access to confirmatory laboratory tests may be restricted. Additionally, they contribute to the standardization of clinical reasoning and optimization of healthcare resources during outbreaks, particularly in scenarios of co-circulation of multiple arboviruses (12,34,40).

Epidemiological context further influences both disease occurrence and model performance. Dengue is predominantly an urban disease, often affecting younger populations and disproportionately impacting black children in settings marked by social and environmental vulnerability, particularly towards the end of the summer season when vector density peaks. In contrast, OF fever has been more frequently associated with rural or peri-urban environments, affecting predominantly adult populations and occurring in the early summer, reflecting distinct ecological and transmission dynamics (2). Chikungunya, while also transmitted by urban-adapted vectors, tends to affect adults more severely and is characterized by debilitating joint symptoms, with transmission patterns that may overlap with dengue but often extend beyond peak dengue seasons depending on local vector competence and population immunity (44,45).

Taken together, these findings highlight that both clinical overlap and data quality constraints within surveillance systems can substantially influence the accuracy of predictive models. Integrating epidemiological variables—such as age, race, seasonality and place of residence—alongside improved completeness of clinical data may enhance model performance and support more accurate syndromic classification of arboviral diseases in endemic settings (46,47).

Clinical prediction models for seasonal infectious diseases usually employ one of four primary approaches to incorporate temporal patterns: rolling windows with dynamic thresholds, sliding window and ensemble methods, direct climate data integration with temporal lags, and temporal feature engineering. Studies of dengue used climate variables (temperature, rainfall, humidity, visibility) lagged by 3 months (48), categorical seasonal predictors (monsoon, winter) (36), or 30-day rolling windows (38), while models for hospital-acquired bacterial infections used sliding windows ranging from 3 to 9 months around the prediction time (49).

Direct climate integration with 3-month lags improves dengue prediction accuracy (48), while dynamic thresholds that continuously learned from recent cases maintained consistent negative predictive values above 90% throughout the year despite seasonal prevalence fluctuations (38). The effectiveness of different approaches depended critically on context: local, high-resolution meteorological data with appropriate lag times proved predictive (48), while aggregated regional climate data did not improve performance (38).

For vector-borne diseases like dengue, both climate-based and temporal window approaches showed benefit (38), whereas hospital-acquired infections responded well to sliding windows and ensemble methods without requiring external climate data (49). The evidence indicates that seasonal incorporation improves predictive performance across multiple infectious diseases (49), but optimal implementation requires matching the approach to disease transmission mechanisms, available data granularity, and geographical context (49). In our context, considering the characteristics of the surveillance data, we used the epidemiological week as a proxy for a temporal feature indicating seasonality. This significantly increased all models’ performance, but it assumes that the seasonality is stable over time. This may be true for dengue, but we do not know that yet regarding OF. If different seasonality becomes evident in the future, the models will require an update, or the user may use a different week of symptoms to best represent seasonality changes.

In the broader context of arboviral diseases, including chikungunya, systematic reviews demonstrate that predictive modelling remains predominantly focused on binary classification tasks, particularly distinguishing dengue cases from non-dengue febrile illnesses. Only a limited number of studies have addressed multiclass classification involving dengue and chikungunya simultaneously, and virtually none have incorporated emerging arboviruses such as OF into diagnostic models. This gap is particularly relevant in endemic settings where co-circulation is common and clinical overlap is substantial (22).

Despite these advances, there are several methodological and operational challenges. These include heterogeneity in study design, lack of standardized datasets, limited external validation, and insufficient integration of clinical and laboratory variables (22,23,26,38). Additionally, many models are developed using retrospective datasets from secondary or tertiary care settings, limiting their applicability in primary health care contexts where diagnostic uncertainty is highest. Another key limitation is the predominance of single-disease models, which fail to reflect real-world scenarios of arboviral co-infection and syndromic overlap (26).

Nevertheless, predictive models offer important advantages for clinical and epidemiological practice. When integrated into decision-support systems, these tools can enhance diagnostic accuracy, support early case identification, and optimize resource allocation, particularly in resource-limited settings (40,26). Furthermore, models capable of incorporating heterogeneous data sources—such as clinical features, laboratory findings, and environmental variables, may provide a more comprehensive approach to arboviral surveillance and diagnosis (23). The development of multi-disease, externally validated, and clinically integrated models represents a critical priority for improving the management of emerging arboviruses such as OF in endemic regions.

Within the clinical diagnostic pathway, predictive models can be integrated as decision-support tools to refine the rational use of complementary tests. For instance, patients classified as high probability for Dengue based on clinical–laboratory models may be prioritized for confirmatory testing (e.g. NS1 antigen detection or RT–PCR), while those with lower predicted probability may be managed with clinical observation or alternative diagnostic hypotheses (24,26). This stratified approach may optimize the use of limited laboratory resources, reduce unnecessary testing and improve turnaround times, particularly in resource-constrained settings.

From a surveillance perspective, integrating predictive models into routine health information systems can enhance case detection and reporting accuracy. Model outputs can be used to flag probable arboviral cases in real time, support syndromic surveillance and guide field investigations, especially during periods of co-epidemics (40,23).

## CONCLUSIONS

In conclusion, this research was able to identify predictors that most discriminate dengue, chikungunya, and OF, to develop and validate predictive models, one for each condition, with moderate to very good performance. The applicability is discussed, and it is reasonable to use these models in diagnostic investigation and surveillance within the Brazilian context. A web application with these models is available at https://pedrobrasil.shinyapps.io/INDWELL/. Integrating the models in different systems is possible; updating the models may be required, as the OF seasonality changes from what was observed in previous periods.

## Supporting information

Supplementary results

## AKNOWLEDGEMENTS

Nothing to declare

## LIST OF ABREVIATIONS

ES: Espírito Santo
FIOCRUZ: Fundação Oswaldo Cruz
INI: Instituto Nacional de Infectologia Evandro Chagas
WHO: World Health Organization
OF: Oropouche Fever
OROV: Oropouche virus
OOB: Out-of-Bag
PAHO: Pan American Health Organization

## DATA AVAILABILITY STATEMENT

The authors provided data within the submission and can provide additional information upon reasonable request.

## AUTHORS CONTRIBUTIONS

Elry Cristine Nickel Valerio - conceptualization, investigation, writing - original draft

Gabriela Maria Coli Seidel - data curation, writing - review

Rafael da Silva Nunes - data curation, writing - review

Pedro Emmanuel Alvarenga Americano do Brasil - conceptualization, methodology, visualization, roles/writing - original draft, writing - review & editing

## DECLARATION OF GENERATIVE AI AND AI-ASSISTED TECHNOLOGIES IN THE WRITING PROCESS

During the preparation of this work, the author(s) used Grammarly and Microsoft Co-Pilot to check grammar, spelling, reduce text word count, and increase reading fluidity. After using these tools/services, the author(s) reviewed and edited the content as needed and take full responsibility for the content of the publication.

