## Supplementary results for "Oropouche, Dengue, and Chikungunya differential diagnosis. Development and validation of predictive models with surveillance data from Espírito Santo/Brazil"

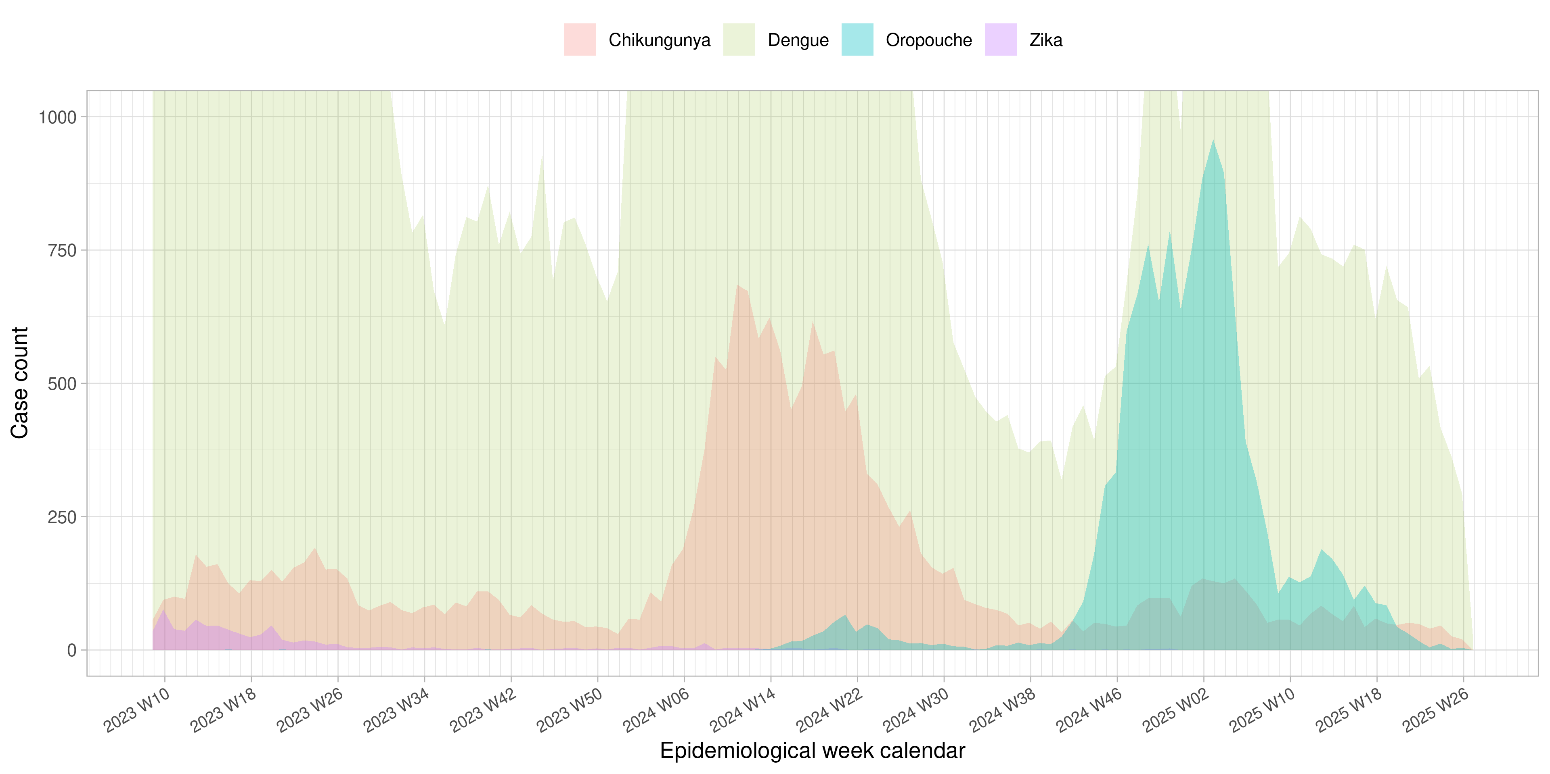

**Figure S 1:** Time series (reduced count scale) of oropouche, dengue, zika and chikungunya case counts from March 2023 to July 2025

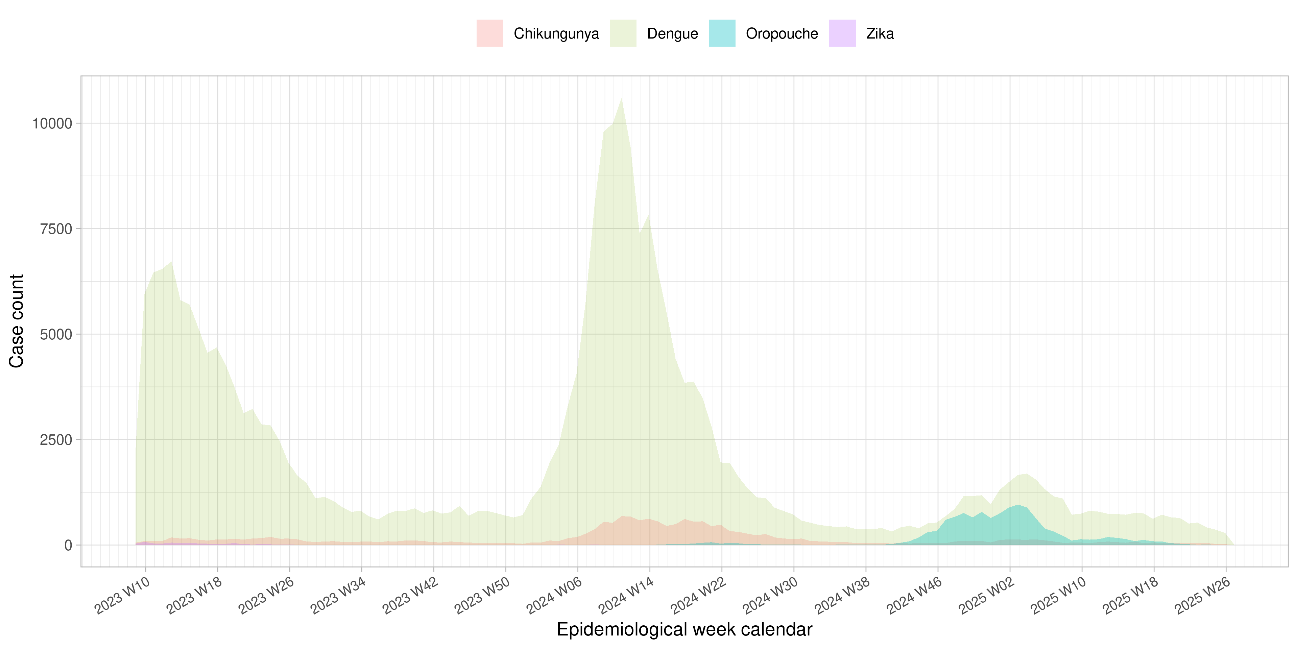

**Figure S 2**: Time series (original count scale) of oropouche, dengue, zika and chikungunya case counts from March 2023 to July 2025

Table S 1Dengue dataset variables from the original notification form (including epidemiological, clinical at presentations and disease progressions) counts and frequencies by case diagnostic investigation outcome.

|  | **Not Dengue (N=209893)** | **Dengue (N=284133)** | **Dengue with alarm (N=6564)** | **Severe Dengue (N=287)** | **Overall (N=500877)** |
| --- | --- | --- | --- | --- | --- |
| **Valid CNS number** |  |  |  |  |  |
| Yes | 208,513 (99.3%) | 280,507 (98.7%) | 6,401 (97.5%) | 284 (99.0%) | 495,705 (99.0%) |
| No | 1,380 (0.7%) | 3,626 (1.3%) | 163 (2.5%) | 3 (1.0%) | 5,172 (1.0%) |
| **Age at notification** |  |  |  |  |  |
| Mean (SD) | 35.0 (19.7) | 35.2 (18.5) | 37.2 (22.2) | 51.8 (25.4) | 35.2 (19.1) |
| Median [Min, Max] | 33.0 [0, 108] | 33.0 [0, 110] | 34.0 [0, 98.0] | 55.0 [0, 97.0] | 33.0 [0, 110] |
| **Age at notification** |  |  |  |  |  |
| [0,1] | 3,781 (1.8%) | 2,155 (0.8%) | 94 (1.4%) | 4 (1.4%) | 6,034 (1.2%) |
| (1,12] | 23,515 (11.2%) | 26,009 (9.2%) | 815 (12.4%) | 21 (7.3%) | 50,360 (10.1%) |
| (12,18] | 19,207 (9.2%) | 27,449 (9.7%) | 705 (10.7%) | 12 (4.2%) | 47,373 (9.5%) |
| (18,25] | 29,078 (13.9%) | 45,155 (15.9%) | 822 (12.5%) | 13 (4.5%) | 75,068 (15.0%) |
| (25,35] | 37,743 (18.0%) | 53,526 (18.8%) | 955 (14.5%) | 39 (13.6%) | 92,263 (18.4%) |
| (35,45] | 35,160 (16.8%) | 48,903 (17.2%) | 891 (13.6%) | 30 (10.5%) | 84,984 (17.0%) |
| (45,55] | 26,137 (12.5%) | 35,700 (12.6%) | 731 (11.1%) | 31 (10.8%) | 62,599 (12.5%) |
| (55,65] | 18,763 (8.9%) | 25,573 (9.0%) | 672 (10.2%) | 31 (10.8%) | 45,039 (9.0%) |
| (65,75] | 10,970 (5.2%) | 13,910 (4.9%) | 504 (7.7%) | 48 (16.7%) | 25,432 (5.1%) |
| (75,85] | 4,347 (2.1%) | 4,658 (1.6%) | 298 (4.5%) | 33 (11.5%) | 9,336 (1.9%) |
| (85,95] | 1,125 (0.5%) | 1,037 (0.4%) | 75 (1.1%) | 22 (7.7%) | 2,259 (0.5%) |
| Missing | 67 (0.0%) | 58 (0.0%) | 2 (0.0%) | 3 (1.0%) | 130 (0.0%) |
| **Sex at notification** |  |  |  |  |  |
| Female | 112,677 (53.7%) | 155,088 (54.6%) | 3,594 (54.8%) | 155 (54.0%) | 271,514 (54.2%) |
| Male | 97,203 (46.3%) | 129,032 (45.4%) | 2,970 (45.2%) | 132 (46.0%) | 229,337 (45.8%) |
| Ignored | 13 (0.0%) | 13 (0.0%) | 0 (0%) | 0 (0%) | 26 (0.0%) |
| **Pregnancy** |  |  |  |  |  |
| 1st trimester | 663 (0.3%) | 687 (0.2%) | 17 (0.3%) | 0 (0%) | 1,367 (0.3%) |
| 2nd trimester | 877 (0.4%) | 836 (0.3%) | 31 (0.5%) | 0 (0%) | 1,744 (0.3%) |
| 3rd trimester | 678 (0.3%) | 637 (0.2%) | 20 (0.3%) | 0 (0%) | 1,335 (0.3%) |
| Gestational age unknown | 614 (0.3%) | 617 (0.2%) | 10 (0.2%) | 1 (0.3%) | 1,242 (0.2%) |
| No | 89,049 (42.4%) | 112,332 (39.5%) | 2,701 (41.1%) | 124 (43.2%) | 204,206 (40.8%) |
| Not applicable | 99,997 (47.6%) | 120,697 (42.5%) | 2,206 (33.6%) | 134 (46.7%) | 223,034 (44.5%) |
| Ignored/Unknown | 18,015 (8.6%) | 48,327 (17.0%) | 1,579 (24.1%) | 28 (9.8%) | 67,949 (13.6%) |
| **Race or skin color** |  |  |  |  |  |
| White | 76,487 (36.4%) | 74,085 (26.1%) | 1,882 (28.7%) | 103 (35.9%) | 152,557 (30.5%) |
| Black | 11,486 (5.5%) | 17,408 (6.1%) | 365 (5.6%) | 13 (4.5%) | 29,272 (5.8%) |
| Yellow | 19,143 (9.1%) | 24,693 (8.7%) | 329 (5.0%) | 15 (5.2%) | 44,180 (8.8%) |
| Brown | 92,815 (44.2%) | 136,630 (48.1%) | 3,003 (45.7%) | 132 (46.0%) | 232,580 (46.4%) |
| Indigenous | 228 (0.1%) | 219 (0.1%) | 4 (0.1%) | 0 (0%) | 451 (0.1%) |
| Ignored/Unknown | 9,734 (4.6%) | 31,098 (10.9%) | 981 (14.9%) | 24 (8.4%) | 41,837 (8.4%) |
| **Indigenous etnicity** |  |  |  |  |  |
| Tupiniquim | 120 (0.1%) | 45 (0.0%) | 0 (0%) | 0 (0%) | 165 (0.0%) |
| Guarani | 20 (0.0%) | 23 (0.0%) | 0 (0%) | 0 (0%) | 43 (0.0%) |
| Ignored/Unknown | 88 (0.0%) | 151 (0.1%) | 4 (0.1%) | 0 (0%) | 243 (0.0%) |
| Missing | 209,665 (99.9%) | 283,914 (99.9%) | 6,560 (99.9%) | 287 (100%) | 500,426 (99.9%) |
| **Person with special needs** |  |  |  |  |  |
| Yes | 123 (0.1%) | 361 (0.1%) | 45 (0.7%) | 3 (1.0%) | 532 (0.1%) |
| No | 209,770 (99.9%) | 283,772 (99.9%) | 6,519 (99.3%) | 284 (99.0%) | 500,345 (99.9%) |
| **Homeless** |  |  |  |  |  |
| Yes | 37 (0.0%) | 132 (0.0%) | 14 (0.2%) | 0 (0%) | 183 (0.0%) |
| No | 209,856 (100.0%) | 284,001 (100.0%) | 6,550 (99.8%) | 287 (100%) | 500,694 (100.0%) |
| **Education** |  |  |  |  |  |
| Illiterate | 1,549 (0.7%) | 1,222 (0.4%) | 18 (0.3%) | 9 (3.1%) | 2,798 (0.6%) |
| Elementary School Incomplete | 10,319 (4.9%) | 9,445 (3.3%) | 238 (3.6%) | 14 (4.9%) | 20,016 (4.0%) |
| Elementary School Complete | 6,418 (3.1%) | 5,700 (2.0%) | 93 (1.4%) | 11 (3.8%) | 12,222 (2.4%) |
| Middle School Incomplete | 19,631 (9.4%) | 18,871 (6.6%) | 332 (5.1%) | 15 (5.2%) | 38,849 (7.8%) |
| Middle School Complete | 10,478 (5.0%) | 9,999 (3.5%) | 188 (2.9%) | 14 (4.9%) | 20,679 (4.1%) |
| High School Incomplete | 12,304 (5.9%) | 13,466 (4.7%) | 317 (4.8%) | 8 (2.8%) | 26,095 (5.2%) |
| High School Complete | 35,471 (16.9%) | 42,609 (15.0%) | 620 (9.4%) | 25 (8.7%) | 78,725 (15.7%) |
| Higher Education Incomplete | 3,199 (1.5%) | 4,581 (1.6%) | 134 (2.0%) | 4 (1.4%) | 7,918 (1.6%) |
| Higher Education Complete | 10,669 (5.1%) | 13,140 (4.6%) | 315 (4.8%) | 10 (3.5%) | 24,134 (4.8%) |
| Ignored/Unknown | 85,102 (40.5%) | 148,698 (52.3%) | 3,726 (56.8%) | 161 (56.1%) | 237,687 (47.5%) |
| Not applicable | 14,753 (7.0%) | 16,402 (5.8%) | 583 (8.9%) | 16 (5.6%) | 31,754 (6.3%) |
| **Home address** |  |  |  |  |  |
| Urban | 143,405 (68.3%) | 243,831 (85.8%) | 6,070 (92.5%) | 244 (85.0%) | 393,550 (78.6%) |
| Rural | 46,294 (22.1%) | 27,204 (9.6%) | 304 (4.6%) | 29 (10.1%) | 73,831 (14.7%) |
| Peri-urban | 513 (0.2%) | 1,014 (0.4%) | 29 (0.4%) | 1 (0.3%) | 1,557 (0.3%) |
| Unknown | 19,681 (9.4%) | 12,075 (4.2%) | 161 (2.5%) | 13 (4.5%) | 31,930 (6.4%) |
| Missing | 0 (0%) | 9 (0.0%) | 0 (0%) | 0 (0%) | 9 (0.0%) |
| **Autochthonous case** |  |  |  |  |  |
| No | 454 (0.2%) | 445 (0.2%) | 34 (0.5%) | 4 (1.4%) | 937 (0.2%) |
| Yes | 208,869 (99.5%) | 283,298 (99.7%) | 6,499 (99.0%) | 277 (96.5%) | 498,943 (99.6%) |
| Unknown | 239 (0.1%) | 348 (0.1%) | 27 (0.4%) | 5 (1.7%) | 619 (0.1%) |
| Missing | 331 (0.2%) | 42 (0.0%) | 4 (0.1%) | 1 (0.3%) | 378 (0.1%) |
| **Fever** |  |  |  |  |  |
| No | 63,661 (30.3%) | 50,790 (17.9%) | 1,065 (16.2%) | 59 (20.6%) | 115,575 (23.1%) |
| Yes | 146,232 (69.7%) | 233,343 (82.1%) | 5,499 (83.8%) | 228 (79.4%) | 385,302 (76.9%) |
| **Headache** |  |  |  |  |  |
| No | 60,827 (29.0%) | 57,616 (20.3%) | 1,771 (27.0%) | 131 (45.6%) | 120,345 (24.0%) |
| Yes | 149,066 (71.0%) | 226,517 (79.7%) | 4,793 (73.0%) | 156 (54.4%) | 380,532 (76.0%) |
| **Vomiting** |  |  |  |  |  |
| No | 166,557 (79.4%) | 224,562 (79.0%) | 4,251 (64.8%) | 176 (61.3%) | 395,546 (79.0%) |
| Yes | 43,336 (20.6%) | 59,571 (21.0%) | 2,313 (35.2%) | 111 (38.7%) | 105,331 (21.0%) |
| **Back pain** |  |  |  |  |  |
| No | 167,277 (79.7%) | 222,947 (78.5%) | 4,859 (74.0%) | 224 (78.0%) | 395,307 (78.9%) |
| Yes | 42,616 (20.3%) | 61,186 (21.5%) | 1,705 (26.0%) | 63 (22.0%) | 105,570 (21.1%) |
| **Arthritis** |  |  |  |  |  |
| No | 196,568 (93.7%) | 265,752 (93.5%) | 5,985 (91.2%) | 265 (92.3%) | 468,570 (93.6%) |
| Yes | 13,325 (6.3%) | 18,381 (6.5%) | 579 (8.8%) | 22 (7.7%) | 32,307 (6.5%) |
| **Petechiae** |  |  |  |  |  |
| No | 201,974 (96.2%) | 268,843 (94.6%) | 5,910 (90.0%) | 251 (87.5%) | 476,978 (95.2%) |
| Yes | 7,919 (3.8%) | 15,290 (5.4%) | 654 (10.0%) | 36 (12.5%) | 23,899 (4.8%) |
| **Tourniquet test** |  |  |  |  |  |
| No | 208,366 (99.3%) | 279,440 (98.3%) | 6,262 (95.4%) | 283 (98.6%) | 494,351 (98.7%) |
| Yes | 1,527 (0.7%) | 4,693 (1.7%) | 302 (4.6%) | 4 (1.4%) | 6,526 (1.3%) |
| **Myalgia** |  |  |  |  |  |
| No | 71,624 (34.1%) | 65,720 (23.1%) | 1,562 (23.8%) | 89 (31.0%) | 138,995 (27.8%) |
| Yes | 138,269 (65.9%) | 218,413 (76.9%) | 5,002 (76.2%) | 198 (69.0%) | 361,882 (72.2%) |
| **Rash** |  |  |  |  |  |
| No | 202,779 (96.6%) | 269,490 (94.8%) | 5,854 (89.2%) | 269 (93.7%) | 478,392 (95.5%) |
| Yes | 7,114 (3.4%) | 14,643 (5.2%) | 710 (10.8%) | 18 (6.3%) | 22,485 (4.5%) |
| **Nausea** |  |  |  |  |  |
| No | 140,947 (67.2%) | 181,204 (63.8%) | 3,369 (51.3%) | 176 (61.3%) | 325,696 (65.0%) |
| Yes | 68,946 (32.8%) | 102,929 (36.2%) | 3,195 (48.7%) | 111 (38.7%) | 175,181 (35.0%) |
| **Conjunctivitis** |  |  |  |  |  |
| No | 208,267 (99.2%) | 280,723 (98.8%) | 6,420 (97.8%) | 283 (98.6%) | 495,693 (99.0%) |
| Yes | 1,626 (0.8%) | 3,410 (1.2%) | 144 (2.2%) | 4 (1.4%) | 5,184 (1.0%) |
| **Intense arthralgia** |  |  |  |  |  |
| No | 175,340 (83.5%) | 223,124 (78.5%) | 4,970 (75.7%) | 236 (82.2%) | 403,670 (80.6%) |
| Yes | 34,553 (16.5%) | 61,009 (21.5%) | 1,594 (24.3%) | 51 (17.8%) | 97,207 (19.4%) |
| **Leukopenia** |  |  |  |  |  |
| No | 207,455 (98.8%) | 278,140 (97.9%) | 5,195 (79.1%) | 232 (80.8%) | 491,022 (98.0%) |
| Yes | 2,438 (1.2%) | 5,993 (2.1%) | 1,369 (20.9%) | 55 (19.2%) | 9,855 (2.0%) |
| **Retro-orbital pain** |  |  |  |  |  |
| No | 157,022 (74.8%) | 187,493 (66.0%) | 4,358 (66.4%) | 227 (79.1%) | 349,100 (69.7%) |
| Yes | 52,871 (25.2%) | 96,640 (34.0%) | 2,206 (33.6%) | 60 (20.9%) | 151,777 (30.3%) |
| **Diabetes** |  |  |  |  |  |
| No | 204,549 (97.5%) | 275,760 (97.1%) | 6,074 (92.5%) | 232 (80.8%) | 486,615 (97.2%) |
| Yes | 5,112 (2.4%) | 8,140 (2.9%) | 486 (7.4%) | 55 (19.2%) | 13,793 (2.8%) |
| Ignored | 232 (0.1%) | 233 (0.1%) | 4 (0.1%) | 0 (0%) | 469 (0.1%) |
| **Hepatopathies** |  |  |  |  |  |
| No | 209,465 (99.8%) | 283,591 (99.8%) | 6,504 (99.1%) | 281 (97.9%) | 499,841 (99.8%) |
| Yes | 198 (0.1%) | 313 (0.1%) | 55 (0.8%) | 5 (1.7%) | 571 (0.1%) |
| Ignored | 230 (0.1%) | 229 (0.1%) | 5 (0.1%) | 1 (0.3%) | 465 (0.1%) |
| **Arterial hypertension** |  |  |  |  |  |
| No | 195,694 (93.2%) | 263,008 (92.6%) | 5,554 (84.6%) | 183 (63.8%) | 464,439 (92.7%) |
| Yes | 13,970 (6.7%) | 20,894 (7.4%) | 1,007 (15.3%) | 104 (36.2%) | 35,975 (7.2%) |
| Ignored | 229 (0.1%) | 231 (0.1%) | 3 (0.0%) | 0 (0%) | 463 (0.1%) |
| **Autoimmune diseases** |  |  |  |  |  |
| No | 209,127 (99.6%) | 283,011 (99.6%) | 6,483 (98.8%) | 280 (97.6%) | 498,901 (99.6%) |
| Yes | 537 (0.3%) | 885 (0.3%) | 76 (1.2%) | 7 (2.4%) | 1,505 (0.3%) |
| Ignored | 229 (0.1%) | 237 (0.1%) | 5 (0.1%) | 0 (0%) | 471 (0.1%) |
| **Hematological diseases** |  |  |  |  |  |
| No | 209,504 (99.8%) | 283,630 (99.8%) | 6,516 (99.3%) | 281 (97.9%) | 499,931 (99.8%) |
| Yes | 168 (0.1%) | 279 (0.1%) | 42 (0.6%) | 6 (2.1%) | 495 (0.1%) |
| Ignored | 221 (0.1%) | 224 (0.1%) | 6 (0.1%) | 0 (0%) | 451 (0.1%) |
| **Chronic kidney disease** |  |  |  |  |  |
| No | 209,383 (99.8%) | 283,603 (99.8%) | 6,504 (99.1%) | 281 (97.9%) | 499,771 (99.8%) |
| Yes | 289 (0.1%) | 303 (0.1%) | 55 (0.8%) | 6 (2.1%) | 653 (0.1%) |
| Ignored | 221 (0.1%) | 227 (0.1%) | 5 (0.1%) | 0 (0%) | 453 (0.1%) |
| **Peptic acid disease** |  |  |  |  |  |
| No | 209,613 (99.9%) | 283,718 (99.9%) | 6,531 (99.5%) | 285 (99.3%) | 500,147 (99.9%) |
| Yes | 57 (0.0%) | 195 (0.1%) | 28 (0.4%) | 2 (0.7%) | 282 (0.1%) |
| Ignored | 223 (0.1%) | 220 (0.1%) | 5 (0.1%) | 0 (0%) | 448 (0.1%) |
| **Dengue IgM result** |  |  |  |  |  |
| Negative | 34,679 (16.5%) | 10,936 (3.8%) | 504 (7.7%) | 51 (17.8%) | 46,170 (9.2%) |
| Positive | 458 (0.2%) | 26,717 (9.4%) | 989 (15.1%) | 88 (30.7%) | 28,252 (5.6%) |
| Inconclusive | 954 (0.5%) | 1,129 (0.4%) | 28 (0.4%) | 5 (1.7%) | 2,116 (0.4%) |
| Not performed | 172,445 (82.2%) | 243,640 (85.7%) | 4,976 (75.8%) | 133 (46.3%) | 421,194 (84.1%) |
| Missing | 1,357 (0.6%) | 1,711 (0.6%) | 67 (1.0%) | 10 (3.5%) | 3,145 (0.6%) |
| **Dengue NS1 result** |  |  |  |  |  |
| Negative | 39,247 (18.7%) | 14,519 (5.1%) | 350 (5.3%) | 25 (8.7%) | 54,141 (10.8%) |
| Positive | 655 (0.3%) | 36,490 (12.8%) | 1,360 (20.7%) | 74 (25.8%) | 38,579 (7.7%) |
| Inconclusive | 565 (0.3%) | 667 (0.2%) | 8 (0.1%) | 0 (0%) | 1,240 (0.2%) |
| Not performed | 167,615 (79.9%) | 231,341 (81.4%) | 4,808 (73.2%) | 185 (64.5%) | 403,949 (80.6%) |
| Missing | 1,811 (0.9%) | 1,116 (0.4%) | 38 (0.6%) | 3 (1.0%) | 2,968 (0.6%) |
| **Dengue viral isolation result** |  |  |  |  |  |
| Negative | 625 (0.3%) | 216 (0.1%) | 15 (0.2%) | 2 (0.7%) | 858 (0.2%) |
| Positive | 4 (0.0%) | 624 (0.2%) | 17 (0.3%) | 5 (1.7%) | 650 (0.1%) |
| Inconclusive | 71 (0.0%) | 71 (0.0%) | 0 (0%) | 0 (0%) | 142 (0.0%) |
| Not performed | 208,970 (99.6%) | 282,984 (99.6%) | 6,520 (99.3%) | 278 (96.9%) | 498,752 (99.6%) |
| Missing | 223 (0.1%) | 238 (0.1%) | 12 (0.2%) | 2 (0.7%) | 475 (0.1%) |
| **Dengue RT-PCR result** |  |  |  |  |  |
| Negative | 46,853 (22.3%) | 8,034 (2.8%) | 288 (4.4%) | 22 (7.7%) | 55,197 (11.0%) |
| Positive | 130 (0.1%) | 11,755 (4.1%) | 388 (5.9%) | 54 (18.8%) | 12,327 (2.5%) |
| Inconclusive | 670 (0.3%) | 638 (0.2%) | 6 (0.1%) | 1 (0.3%) | 1,315 (0.3%) |
| Not performed | 161,628 (77.0%) | 263,268 (92.7%) | 5,859 (89.3%) | 205 (71.4%) | 430,960 (86.0%) |
| Missing | 612 (0.3%) | 438 (0.2%) | 23 (0.4%) | 5 (1.7%) | 1,078 (0.2%) |
| **Dengue RT-PCR serotype** |  |  |  |  |  |
| DENV-1 | 65 (0.0%) | 6,710 (2.4%) | 278 (4.2%) | 24 (8.4%) | 7,077 (1.4%) |
| DENV-2 | 26 (0.0%) | 2,407 (0.8%) | 82 (1.2%) | 11 (3.8%) | 2,526 (0.5%) |
| DENV-3 | 0 (0%) | 5 (0.0%) | 0 (0%) | 0 (0%) | 5 (0.0%) |
| DENV-4 | 0 (0%) | 0 (0%) | 0 (0%) | 0 (0%) | 0 (0%) |
| Missing | 209,802 (100.0%) | 275,011 (96.8%) | 6,204 (94.5%) | 252 (87.8%) | 491,269 (98.1%) |
| **Dengue histopathology result** |  |  |  |  |  |
| Dengue compatible | 7 (0.0%) | 484 (0.2%) | 2 (0.0%) | 0 (0%) | 493 (0.1%) |
| Dengue not compatible | 12 (0.0%) | 0 (0%) | 0 (0%) | 1 (0.3%) | 13 (0.0%) |
| Inconclusive | 4 (0.0%) | 17 (0.0%) | 1 (0.0%) | 0 (0%) | 22 (0.0%) |
| Not performed | 5,560 (2.6%) | 4,885 (1.7%) | 170 (2.6%) | 14 (4.9%) | 10,629 (2.1%) |
| Missing | 204,310 (97.3%) | 278,747 (98.1%) | 6,391 (97.4%) | 272 (94.8%) | 489,720 (97.8%) |
| **Dengue immunohistochemistry result** |  |  |  |  |  |
| Negative | 31 (0.0%) | 3 (0.0%) | 0 (0%) | 0 (0%) | 34 (0.0%) |
| Positive | 7 (0.0%) | 417 (0.1%) | 2 (0.0%) | 0 (0%) | 426 (0.1%) |
| Inconclusive | 14 (0.0%) | 15 (0.0%) | 0 (0%) | 0 (0%) | 29 (0.0%) |
| Not performed | 5,549 (2.6%) | 4,868 (1.7%) | 169 (2.6%) | 14 (4.9%) | 10,600 (2.1%) |
| Missing | 204,292 (97.3%) | 278,830 (98.1%) | 6,393 (97.4%) | 273 (95.1%) | 489,788 (97.8%) |
| **Final Dengue classification criteria** |  |  |  |  |  |
| Laboratory | 109,831 (52.3%) | 71,817 (25.3%) | 2,618 (39.9%) | 195 (67.9%) | 184,461 (36.8%) |
| Clinical-epideiological | 98,762 (47.1%) | 209,900 (73.9%) | 3,880 (59.1%) | 86 (30.0%) | 312,628 (62.4%) |
| Not concluded | 271 (0.1%) | 1,085 (0.4%) | 41 (0.6%) | 6 (2.1%) | 1,403 (0.3%) |
| Missing | 1,029 (0.5%) | 1,331 (0.5%) | 25 (0.4%) | 0 (0%) | 2,385 (0.5%) |
| **Case outcome** |  |  |  |  |  |
| Recovered | 54,444 (25.9%) | 241,887 (85.1%) | 4,926 (75.0%) | 144 (50.2%) | 301,401 (60.2%) |
| Death (arbovirus) | 0 (0%) | 0 (0%) | 28 (0.4%) | 117 (40.8%) | 145 (0.0%) |
| Death (other causes) | 333 (0.2%) | 16 (0.0%) | 7 (0.1%) | 10 (3.5%) | 366 (0.1%) |
| Death (investigating) | 1 (0.0%) | 2 (0.0%) | 0 (0%) | 1 (0.3%) | 4 (0.0%) |
| Ignored | 125,467 (59.8%) | 36,537 (12.9%) | 1,519 (23.1%) | 12 (4.2%) | 163,535 (32.6%) |
| Missing | 29,648 (14.1%) | 5,691 (2.0%) | 84 (1.3%) | 3 (1.0%) | 35,426 (7.1%) |
| **Hospitalization** |  |  |  |  |  |
| No | 206,293 (98.3%) | 280,192 (98.6%) | 3,628 (55.3%) | 54 (18.8%) | 490,167 (97.9%) |
| Yes | 3,545 (1.7%) | 3,870 (1.4%) | 2,932 (44.7%) | 233 (81.2%) | 10,580 (2.1%) |
| Ignored | 38 (0.0%) | 63 (0.0%) | 3 (0.0%) | 0 (0%) | 104 (0.0%) |
| Missing | 17 (0.0%) | 8 (0.0%) | 1 (0.0%) | 0 (0%) | 26 (0.0%) |
| **Postural hypotension and/or syncope** |  |  |  |  |  |
| No | 444 (0.2%) | 217 (0.1%) | 5,086 (77.5%) | 128 (44.6%) | 5,875 (1.2%) |
| Yes | 153 (0.1%) | 37 (0.0%) | 1,104 (16.8%) | 111 (38.7%) | 1,405 (0.3%) |
| Ignored | 46 (0.0%) | 9 (0.0%) | 374 (5.7%) | 29 (10.1%) | 458 (0.1%) |
| Missing | 209,250 (99.7%) | 283,870 (99.9%) | 0 (0%) | 19 (6.6%) | 493,139 (98.5%) |
| **Abrupt platelet count drop** |  |  |  |  |  |
| No | 320 (0.2%) | 132 (0.0%) | 2,468 (37.6%) | 75 (26.1%) | 2,995 (0.6%) |
| Yes | 260 (0.1%) | 102 (0.0%) | 3,669 (55.9%) | 167 (58.2%) | 4,198 (0.8%) |
| Ignored | 62 (0.0%) | 18 (0.0%) | 427 (6.5%) | 26 (9.1%) | 533 (0.1%) |
| Missing | 209,251 (99.7%) | 283,881 (99.9%) | 0 (0%) | 19 (6.6%) | 493,151 (98.5%) |
| **Persistent vomiting** |  |  |  |  |  |
| No | 483 (0.2%) | 203 (0.1%) | 5,087 (77.5%) | 170 (59.2%) | 5,943 (1.2%) |
| Yes | 115 (0.1%) | 33 (0.0%) | 1,115 (17.0%) | 68 (23.7%) | 1,331 (0.3%) |
| Ignored | 42 (0.0%) | 14 (0.0%) | 362 (5.5%) | 30 (10.5%) | 448 (0.1%) |
| Missing | 209,253 (99.7%) | 283,883 (99.9%) | 0 (0%) | 19 (6.6%) | 493,155 (98.5%) |
| **Intense and continuous abdominal pain** |  |  |  |  |  |
| No | 381 (0.2%) | 170 (0.1%) | 4,072 (62.0%) | 153 (53.3%) | 4,776 (1.0%) |
| Yes | 217 (0.1%) | 75 (0.0%) | 2,171 (33.1%) | 96 (33.4%) | 2,559 (0.5%) |
| Ignored | 44 (0.0%) | 12 (0.0%) | 321 (4.9%) | 19 (6.6%) | 396 (0.1%) |
| Missing | 209,251 (99.7%) | 283,876 (99.9%) | 0 (0%) | 19 (6.6%) | 493,146 (98.5%) |
| **Lethargy or irritability** |  |  |  |  |  |
| No | 475 (0.2%) | 203 (0.1%) | 4,932 (75.1%) | 143 (49.8%) | 5,753 (1.1%) |
| Yes | 112 (0.1%) | 26 (0.0%) | 1,219 (18.6%) | 89 (31.0%) | 1,446 (0.3%) |
| Ignored | 54 (0.0%) | 15 (0.0%) | 413 (6.3%) | 36 (12.5%) | 518 (0.1%) |
| Missing | 209,252 (99.7%) | 283,889 (99.9%) | 0 (0%) | 19 (6.6%) | 493,160 (98.5%) |
| **Mucosal bleeding/other hemorrhages** |  |  |  |  |  |
| No | 448 (0.2%) | 183 (0.1%) | 5,004 (76.2%) | 160 (55.7%) | 5,795 (1.2%) |
| Yes | 148 (0.1%) | 46 (0.0%) | 1,187 (18.1%) | 79 (27.5%) | 1,460 (0.3%) |
| Ignored | 44 (0.0%) | 19 (0.0%) | 373 (5.7%) | 29 (10.1%) | 465 (0.1%) |
| Missing | 209,253 (99.7%) | 283,885 (99.9%) | 0 (0%) | 19 (6.6%) | 493,157 (98.5%) |
| **Progressive hematocrit increase** |  |  |  |  |  |
| No | 529 (0.3%) | 212 (0.1%) | 5,343 (81.4%) | 170 (59.2%) | 6,254 (1.2%) |
| Yes | 21 (0.0%) | 18 (0.0%) | 393 (6.0%) | 49 (17.1%) | 481 (0.1%) |
| Ignored | 91 (0.0%) | 20 (0.0%) | 828 (12.6%) | 49 (17.1%) | 988 (0.2%) |
| Missing | 209,252 (99.7%) | 283,883 (99.9%) | 0 (0%) | 19 (6.6%) | 493,154 (98.5%) |
| **Hepatomegaly >= 2cm** |  |  |  |  |  |
| No | 534 (0.3%) | 216 (0.1%) | 5,515 (84.0%) | 207 (72.1%) | 6,472 (1.3%) |
| Yes | 20 (0.0%) | 4 (0.0%) | 124 (1.9%) | 10 (3.5%) | 158 (0.0%) |
| Ignored | 86 (0.0%) | 24 (0.0%) | 925 (14.1%) | 51 (17.8%) | 1,086 (0.2%) |
| Missing | 209,253 (99.7%) | 283,889 (99.9%) | 0 (0%) | 19 (6.6%) | 493,161 (98.5%) |
| **Fluid accumulation** |  |  |  |  |  |
| No | 523 (0.2%) | 210 (0.1%) | 5,669 (86.4%) | 171 (59.6%) | 6,573 (1.3%) |
| Yes | 52 (0.0%) | 11 (0.0%) | 176 (2.7%) | 53 (18.5%) | 292 (0.1%) |
| Ignored | 64 (0.0%) | 24 (0.0%) | 719 (11.0%) | 44 (15.3%) | 851 (0.2%) |
| Missing | 209,254 (99.7%) | 283,888 (99.9%) | 0 (0%) | 19 (6.6%) | 493,161 (98.5%) |
| **Weak or undetectable pulse** |  |  |  |  |  |
| No | 54 (0.0%) | 44 (0.0%) | 77 (1.2%) | 174 (60.6%) | 349 (0.1%) |
| Yes | 24 (0.0%) | 2 (0.0%) | 2 (0.0%) | 73 (25.4%) | 101 (0.0%) |
| Ignored | 23 (0.0%) | 1 (0.0%) | 7 (0.1%) | 40 (13.9%) | 71 (0.0%) |
| Missing | 209,792 (100.0%) | 284,086 (100.0%) | 6,478 (98.7%) | 0 (0%) | 500,356 (99.9%) |
| **Convergent pulse pressure <= 20 mmHg** |  |  |  |  |  |
| No | 62 (0.0%) | 43 (0.0%) | 72 (1.1%) | 217 (75.6%) | 394 (0.1%) |
| Yes | 8 (0.0%) | 0 (0%) | 4 (0.1%) | 24 (8.4%) | 36 (0.0%) |
| Ignored | 31 (0.0%) | 4 (0.0%) | 7 (0.1%) | 46 (16.0%) | 88 (0.0%) |
| Missing | 209,792 (100.0%) | 284,086 (100.0%) | 6,481 (98.7%) | 0 (0%) | 500,359 (99.9%) |
| **Capillary refill time** |  |  |  |  |  |
| No | 48 (0.0%) | 13 (0.0%) | 61 (0.9%) | 177 (61.7%) | 299 (0.1%) |
| Yes | 18 (0.0%) | 30 (0.0%) | 12 (0.2%) | 53 (18.5%) | 113 (0.0%) |
| Ignored | 35 (0.0%) | 4 (0.0%) | 7 (0.1%) | 57 (19.9%) | 103 (0.0%) |
| Missing | 209,792 (100.0%) | 284,086 (100.0%) | 6,484 (98.8%) | 0 (0%) | 500,362 (99.9%) |
| **Fluid accumulation with respiratory insufficiency** |  |  |  |  |  |
| No | 51 (0.0%) | 43 (0.0%) | 75 (1.1%) | 157 (54.7%) | 326 (0.1%) |
| Yes | 24 (0.0%) | 3 (0.0%) | 2 (0.0%) | 86 (30.0%) | 115 (0.0%) |
| Ignored | 26 (0.0%) | 1 (0.0%) | 5 (0.1%) | 44 (15.3%) | 76 (0.0%) |
| Missing | 209,792 (100.0%) | 284,086 (100.0%) | 6,482 (98.8%) | 0 (0%) | 500,360 (99.9%) |
| **Tachycardia** |  |  |  |  |  |
| No | 34 (0.0%) | 36 (0.0%) | 28 (0.4%) | 163 (56.8%) | 261 (0.1%) |
| Yes | 42 (0.0%) | 9 (0.0%) | 45 (0.7%) | 77 (26.8%) | 173 (0.0%) |
| Ignored | 25 (0.0%) | 2 (0.0%) | 5 (0.1%) | 47 (16.4%) | 79 (0.0%) |
| Missing | 209,792 (100.0%) | 284,086 (100.0%) | 6,486 (98.8%) | 0 (0%) | 500,364 (99.9%) |
| **Cold extremities** |  |  |  |  |  |
| No | 48 (0.0%) | 41 (0.0%) | 66 (1.0%) | 162 (56.4%) | 317 (0.1%) |
| Yes | 30 (0.0%) | 4 (0.0%) | 11 (0.2%) | 82 (28.6%) | 127 (0.0%) |
| Ignored | 23 (0.0%) | 2 (0.0%) | 5 (0.1%) | 43 (15.0%) | 73 (0.0%) |
| Missing | 209,792 (100.0%) | 284,086 (100.0%) | 6,482 (98.8%) | 0 (0%) | 500,360 (99.9%) |
| **Late stage arterial hypotension** |  |  |  |  |  |
| No | 56 (0.0%) | 43 (0.0%) | 72 (1.1%) | 153 (53.3%) | 324 (0.1%) |
| Yes | 19 (0.0%) | 2 (0.0%) | 2 (0.0%) | 96 (33.4%) | 119 (0.0%) |
| Ignored | 26 (0.0%) | 2 (0.0%) | 7 (0.1%) | 38 (13.2%) | 73 (0.0%) |
| Missing | 209,792 (100.0%) | 284,086 (100.0%) | 6,483 (98.8%) | 0 (0%) | 500,361 (99.9%) |
| **Hematemesis** |  |  |  |  |  |
| No | 73 (0.0%) | 45 (0.0%) | 76 (1.2%) | 217 (75.6%) | 411 (0.1%) |
| Yes | 7 (0.0%) | 1 (0.0%) | 2 (0.0%) | 30 (10.5%) | 40 (0.0%) |
| Ignored | 21 (0.0%) | 1 (0.0%) | 2 (0.0%) | 40 (13.9%) | 64 (0.0%) |
| Missing | 209,792 (100.0%) | 284,086 (100.0%) | 6,484 (98.8%) | 0 (0%) | 500,362 (99.9%) |
| **Melena** |  |  |  |  |  |
| No | 71 (0.0%) | 45 (0.0%) | 73 (1.1%) | 217 (75.6%) | 406 (0.1%) |
| Yes | 8 (0.0%) | 1 (0.0%) | 4 (0.1%) | 28 (9.8%) | 41 (0.0%) |
| Ignored | 22 (0.0%) | 1 (0.0%) | 2 (0.0%) | 42 (14.6%) | 67 (0.0%) |
| Missing | 209,792 (100.0%) | 284,086 (100.0%) | 6,485 (98.8%) | 0 (0%) | 500,363 (99.9%) |
| **Voluminous metrorrhagia** |  |  |  |  |  |
| No | 79 (0.0%) | 46 (0.0%) | 77 (1.2%) | 238 (82.9%) | 440 (0.1%) |
| Yes | 5 (0.0%) | 0 (0%) | 0 (0%) | 12 (4.2%) | 17 (0.0%) |
| Ignored | 17 (0.0%) | 1 (0.0%) | 5 (0.1%) | 37 (12.9%) | 60 (0.0%) |
| Missing | 209,792 (100.0%) | 284,086 (100.0%) | 6,482 (98.8%) | 0 (0%) | 500,360 (99.9%) |
| **CNS bleeding** |  |  |  |  |  |
| No | 72 (0.0%) | 45 (0.0%) | 72 (1.1%) | 221 (77.0%) | 410 (0.1%) |
| Yes | 4 (0.0%) | 0 (0%) | 0 (0%) | 7 (2.4%) | 11 (0.0%) |
| Ignored | 25 (0.0%) | 2 (0.0%) | 7 (0.1%) | 59 (20.6%) | 93 (0.0%) |
| Missing | 209,792 (100.0%) | 284,086 (100.0%) | 6,485 (98.8%) | 0 (0%) | 500,363 (99.9%) |
| **AST/ALT > 1,000** |  |  |  |  |  |
| No | 68 (0.0%) | 41 (0.0%) | 63 (1.0%) | 223 (77.7%) | 395 (0.1%) |
| Yes | 8 (0.0%) | 1 (0.0%) | 1 (0.0%) | 15 (5.2%) | 25 (0.0%) |
| Ignored | 25 (0.0%) | 5 (0.0%) | 14 (0.2%) | 49 (17.1%) | 93 (0.0%) |
| Missing | 209,792 (100.0%) | 284,086 (100.0%) | 6,486 (98.8%) | 0 (0%) | 500,364 (99.9%) |
| **Myocarditis** |  |  |  |  |  |
| No | 64 (0.0%) | 43 (0.0%) | 71 (1.1%) | 225 (78.4%) | 403 (0.1%) |
| Yes | 5 (0.0%) | 0 (0%) | 0 (0%) | 8 (2.8%) | 13 (0.0%) |
| Ignored | 32 (0.0%) | 4 (0.0%) | 10 (0.2%) | 54 (18.8%) | 100 (0.0%) |
| Missing | 209,792 (100.0%) | 284,086 (100.0%) | 6,483 (98.8%) | 0 (0%) | 500,361 (99.9%) |
| **Altered consciousness** |  |  |  |  |  |
| No | 41 (0.0%) | 44 (0.0%) | 70 (1.1%) | 140 (48.8%) | 295 (0.1%) |
| Yes | 44 (0.0%) | 0 (0%) | 4 (0.1%) | 108 (37.6%) | 156 (0.0%) |
| Ignored | 16 (0.0%) | 3 (0.0%) | 5 (0.1%) | 39 (13.6%) | 63 (0.0%) |
| Missing | 209,792 (100.0%) | 284,086 (100.0%) | 6,485 (98.8%) | 0 (0%) | 500,363 (99.9%) |
| **Other severe organs involvement** |  |  |  |  |  |
| No | 51 (0.0%) | 43 (0.0%) | 70 (1.1%) | 154 (53.7%) | 318 (0.1%) |
| Yes | 31 (0.0%) | 1 (0.0%) | 0 (0%) | 85 (29.6%) | 117 (0.0%) |
| Ignored | 19 (0.0%) | 3 (0.0%) | 8 (0.1%) | 48 (16.7%) | 78 (0.0%) |
| Missing | 209,792 (100.0%) | 284,086 (100.0%) | 6,486 (98.8%) | 0 (0%) | 500,364 (99.9%) |
| IQR = interquartile range; SD = Standard deviation; CNS number = cartão nacional do SUS (used as notification ID); CNS = central nervous system; AST = aspartate aminotransferase; ALT = alanine aminotransferase; RT-PCR = reverse transcription polymerase chain reaction. | | | | | |

**Table S 2** Chikungunya dataset variables from the original notification form (including epidemiological, clinical at presentations and disease progressions) counts and frequencies by case diagnostic investigation outcome.

|  | **Not Chikungunya (N=25782)** | **Chikungunya (N=19506)** | **Overall (N=45288)** |
| --- | --- | --- | --- |
| **Valid CNS number** |  |  |  |
| Yes | 25,654 (99.5%) | 19,306 (99.0%) | 44,960 (99.3%) |
| No | 128 (0.5%) | 200 (1.0%) | 328 (0.7%) |
| **Age at notification** |  |  |  |
| Mean (SD) | 37.3 (18.8) | 44.7 (18.6) | 40.5 (19.1) |
| Median [Min, Max] | 36.0 [0, 99.0] | 45.0 [0, 100] | 40.0 [0, 100] |
| **Age at notification** |  |  |  |
| [0,1] | 203 (0.8%) | 50 (0.3%) | 253 (0.6%) |
| (1,12] | 2,102 (8.2%) | 878 (4.5%) | 2,980 (6.6%) |
| (12,18] | 2,232 (8.7%) | 934 (4.8%) | 3,166 (7.0%) |
| (18,25] | 3,370 (13.1%) | 1,528 (7.8%) | 4,898 (10.8%) |
| (25,35] | 4,515 (17.5%) | 2,784 (14.3%) | 7,299 (16.1%) |
| (35,45] | 4,914 (19.1%) | 3,719 (19.1%) | 8,633 (19.1%) |
| (45,55] | 3,695 (14.3%) | 3,629 (18.6%) | 7,324 (16.2%) |
| (55,65] | 2,645 (10.3%) | 3,262 (16.7%) | 5,907 (13.0%) |
| (65,75] | 1,520 (5.9%) | 1,894 (9.7%) | 3,414 (7.5%) |
| (75,85] | 473 (1.8%) | 712 (3.7%) | 1,185 (2.6%) |
| (85,95] | 109 (0.4%) | 112 (0.6%) | 221 (0.5%) |
| Missing | 4 (0.0%) | 4 (0.0%) | 8 (0.0%) |
| **Sex at notification** |  |  |  |
| Female | 15,143 (58.7%) | 12,502 (64.1%) | 27,645 (61.0%) |
| Male | 10,638 (41.3%) | 7,003 (35.9%) | 17,641 (39.0%) |
| Ignored | 1 (0.0%) | 1 (0.0%) | 2 (0.0%) |
| **Pregnancy** |  |  |  |
| 1st trimester | 152 (0.6%) | 70 (0.4%) | 222 (0.5%) |
| 2nd trimester | 222 (0.9%) | 88 (0.5%) | 310 (0.7%) |
| 3rd trimester | 185 (0.7%) | 96 (0.5%) | 281 (0.6%) |
| Gestational age unknown | 86 (0.3%) | 70 (0.4%) | 156 (0.3%) |
| No | 11,902 (46.2%) | 9,390 (48.1%) | 21,292 (47.0%) |
| Not applicable | 11,585 (44.9%) | 6,728 (34.5%) | 18,313 (40.4%) |
| Ignored/Unknown | 1,650 (6.4%) | 3,064 (15.7%) | 4,714 (10.4%) |
| **Race or skin color** |  |  |  |
| White | 7,198 (27.9%) | 5,360 (27.5%) | 12,558 (27.7%) |
| Black | 1,337 (5.2%) | 1,662 (8.5%) | 2,999 (6.6%) |
| Yellow | 2,499 (9.7%) | 1,902 (9.8%) | 4,401 (9.7%) |
| Brown | 13,096 (50.8%) | 9,861 (50.6%) | 22,957 (50.7%) |
| Indigenous | 43 (0.2%) | 15 (0.1%) | 58 (0.1%) |
| Ignored/Unknown | 1,609 (6.2%) | 706 (3.6%) | 2,315 (5.1%) |
| **Indigenous ethnicity** |  |  |  |
| Tupiniquim | 0 (0%) | 0 (0%) | 0 (0%) |
| Guarani | 0 (0%) | 0 (0%) | 0 (0%) |
| Ignored/Unknown | 0 (0%) | 0 (0%) | 0 (0%) |
| Missing | 25,782 (100%) | 19,506 (100%) | 45,288 (100%) |
| **Person with special needs** |  |  |  |
| Yes | 20 (0.1%) | 49 (0.3%) | 69 (0.2%) |
| No | 25,762 (99.9%) | 19,457 (99.7%) | 45,219 (99.8%) |
| **Homeless** |  |  |  |
| Yes | 3 (0.0%) | 8 (0.0%) | 11 (0.0%) |
| No | 25,779 (100.0%) | 19,498 (100.0%) | 45,277 (100.0%) |
| **Education** |  |  |  |
| Illiterate | 163 (0.6%) | 122 (0.6%) | 285 (0.6%) |
| Elementary School Incomplete | 1,224 (4.7%) | 836 (4.3%) | 2,060 (4.5%) |
| Elementary School Complete | 642 (2.5%) | 460 (2.4%) | 1,102 (2.4%) |
| Middle School Incomplete | 2,380 (9.2%) | 1,473 (7.6%) | 3,853 (8.5%) |
| Middle School Complete | 1,120 (4.3%) | 666 (3.4%) | 1,786 (3.9%) |
| High School Incomplete | 1,489 (5.8%) | 834 (4.3%) | 2,323 (5.1%) |
| High School Complete | 4,822 (18.7%) | 3,255 (16.7%) | 8,077 (17.8%) |
| Higher Education Incomplete | 453 (1.8%) | 358 (1.8%) | 811 (1.8%) |
| Higher Education Complete | 1,615 (6.3%) | 1,275 (6.5%) | 2,890 (6.4%) |
| Ignored/Unknown | 10,731 (41.6%) | 9,329 (47.8%) | 20,060 (44.3%) |
| Not applicable | 1,143 (4.4%) | 898 (4.6%) | 2,041 (4.5%) |
| **Home address** |  |  |  |
| Urban | 18,873 (73.2%) | 17,861 (91.6%) | 36,734 (81.1%) |
| Rural | 3,542 (13.7%) | 759 (3.9%) | 4,301 (9.5%) |
| Peri-urban | 169 (0.7%) | 15 (0.1%) | 184 (0.4%) |
| Unknown | 3,198 (12.4%) | 870 (4.5%) | 4,068 (9.0%) |
| Missing | 0 (0%) | 1 (0.0%) | 1 (0.0%) |
| **Autochthonous case** |  |  |  |
| No | 54 (0.2%) | 63 (0.3%) | 117 (0.3%) |
| Yes | 25,577 (99.2%) | 19,407 (99.5%) | 44,984 (99.3%) |
| Unknown | 60 (0.2%) | 31 (0.2%) | 91 (0.2%) |
| Missing | 91 (0.4%) | 5 (0.0%) | 96 (0.2%) |
| **Fever** |  |  |  |
| No | 7,281 (28.2%) | 8,128 (41.7%) | 15,409 (34.0%) |
| Yes | 18,501 (71.8%) | 11,378 (58.3%) | 29,879 (66.0%) |
| **Headache** |  |  |  |
| No | 6,955 (27.0%) | 9,671 (49.6%) | 16,626 (36.7%) |
| Yes | 18,827 (73.0%) | 9,835 (50.4%) | 28,662 (63.3%) |
| **Vomiting** |  |  |  |
| No | 20,155 (78.2%) | 17,327 (88.8%) | 37,482 (82.8%) |
| Yes | 5,627 (21.8%) | 2,179 (11.2%) | 7,806 (17.2%) |
| **Back pain** |  |  |  |
| No | 19,871 (77.1%) | 15,069 (77.3%) | 34,940 (77.2%) |
| Yes | 5,911 (22.9%) | 4,437 (22.7%) | 10,348 (22.8%) |
| **Arthritis** |  |  |  |
| No | 22,763 (88.3%) | 15,132 (77.6%) | 37,895 (83.7%) |
| Yes | 3,019 (11.7%) | 4,374 (22.4%) | 7,393 (16.3%) |
| **Petechiae** |  |  |  |
| No | 24,044 (93.3%) | 17,795 (91.2%) | 41,839 (92.4%) |
| Yes | 1,738 (6.7%) | 1,711 (8.8%) | 3,449 (7.6%) |
| **Tourniquet test** |  |  |  |
| No | 25,500 (98.9%) | 19,204 (98.5%) | 44,704 (98.7%) |
| Yes | 282 (1.1%) | 302 (1.5%) | 584 (1.3%) |
| **Myalgia** |  |  |  |
| No | 7,662 (29.7%) | 8,544 (43.8%) | 16,206 (35.8%) |
| Yes | 18,120 (70.3%) | 10,962 (56.2%) | 29,082 (64.2%) |
| **Rash** |  |  |  |
| No | 24,270 (94.1%) | 17,289 (88.6%) | 41,559 (91.8%) |
| Yes | 1,512 (5.9%) | 2,217 (11.4%) | 3,729 (8.2%) |
| **Nausea** |  |  |  |
| No | 16,733 (64.9%) | 13,680 (70.1%) | 30,413 (67.2%) |
| Yes | 9,049 (35.1%) | 5,826 (29.9%) | 14,875 (32.8%) |
| **Conjunctivitis** |  |  |  |
| No | 25,438 (98.7%) | 19,030 (97.6%) | 44,468 (98.2%) |
| Yes | 344 (1.3%) | 476 (2.4%) | 820 (1.8%) |
| **Intense arthralgia** |  |  |  |
| No | 17,952 (69.6%) | 7,657 (39.3%) | 25,609 (56.5%) |
| Yes | 7,830 (30.4%) | 11,849 (60.7%) | 19,679 (43.5%) |
| **Leukopenia** |  |  |  |
| No | 25,291 (98.1%) | 19,060 (97.7%) | 44,351 (97.9%) |
| Yes | 491 (1.9%) | 446 (2.3%) | 937 (2.1%) |
| **Retro-orbital pain** |  |  |  |
| No | 18,197 (70.6%) | 14,045 (72.0%) | 32,242 (71.2%) |
| Yes | 7,585 (29.4%) | 5,461 (28.0%) | 13,046 (28.8%) |
| **Diabetes** |  |  |  |
| No | 24,867 (96.5%) | 18,058 (92.6%) | 42,925 (94.8%) |
| Yes | 862 (3.3%) | 1,306 (6.7%) | 2,168 (4.8%) |
| Ignored | 53 (0.2%) | 142 (0.7%) | 195 (0.4%) |
| **Hepatopathies** |  |  |  |
| No | 25,691 (99.6%) | 19,298 (98.9%) | 44,989 (99.3%) |
| Yes | 33 (0.1%) | 53 (0.3%) | 86 (0.2%) |
| Ignored | 58 (0.2%) | 155 (0.8%) | 213 (0.5%) |
| **Arterial hypertension** |  |  |  |
| No | 23,581 (91.5%) | 16,414 (84.1%) | 39,995 (88.3%) |
| Yes | 2,146 (8.3%) | 2,959 (15.2%) | 5,105 (11.3%) |
| Ignored | 55 (0.2%) | 133 (0.7%) | 188 (0.4%) |
| **Autoimmune diseases** |  |  |  |
| No | 25,584 (99.2%) | 19,200 (98.4%) | 44,784 (98.9%) |
| Yes | 138 (0.5%) | 141 (0.7%) | 279 (0.6%) |
| Ignored | 60 (0.2%) | 165 (0.8%) | 225 (0.5%) |
| **Hematological diseases** |  |  |  |
| No | 25,698 (99.7%) | 19,315 (99.0%) | 45,013 (99.4%) |
| Yes | 28 (0.1%) | 32 (0.2%) | 60 (0.1%) |
| Ignored | 56 (0.2%) | 159 (0.8%) | 215 (0.5%) |
| **Chronic kidney disease** |  |  |  |
| No | 25,663 (99.5%) | 19,299 (98.9%) | 44,962 (99.3%) |
| Yes | 63 (0.2%) | 46 (0.2%) | 109 (0.2%) |
| Ignored | 56 (0.2%) | 161 (0.8%) | 217 (0.5%) |
| **Peptic acid disease** |  |  |  |
| No | 25,708 (99.7%) | 19,314 (99.0%) | 45,022 (99.4%) |
| Yes | 18 (0.1%) | 35 (0.2%) | 53 (0.1%) |
| Ignored | 56 (0.2%) | 157 (0.8%) | 213 (0.5%) |
| **First sample serology result (Chikungunya)** |  |  |  |
| Negative | 9,634 (37.4%) | 698 (3.6%) | 10,332 (22.8%) |
| Positive | 80 (0.3%) | 7,202 (36.9%) | 7,282 (16.1%) |
| Inconclusive | 112 (0.4%) | 44 (0.2%) | 156 (0.3%) |
| Not performed | 166 (0.6%) | 60 (0.3%) | 226 (0.5%) |
| Waiting results | 136 (0.5%) | 107 (0.5%) | 243 (0.5%) |
| Missing | 15,654 (60.7%) | 11,395 (58.4%) | 27,049 (59.7%) |
| **Second sample serology result (Chikungunya)** |  |  |  |
| Negative | 678 (2.6%) | 102 (0.5%) | 780 (1.7%) |
| Positive | 2 (0.0%) | 133 (0.7%) | 135 (0.3%) |
| Inconclusive | 4 (0.0%) | 6 (0.0%) | 10 (0.0%) |
| Not performed | 8 (0.0%) | 6 (0.0%) | 14 (0.0%) |
| Waiting results | 28 (0.1%) | 8 (0.0%) | 36 (0.1%) |
| Missing | 25,062 (97.2%) | 19,251 (98.7%) | 44,313 (97.8%) |
| **Chikungunya PRNT result** |  |  |  |
| Negative | 81 (0.3%) | 3 (0.0%) | 84 (0.2%) |
| Positive | 2 (0.0%) | 19 (0.1%) | 21 (0.0%) |
| Inconclusive | 2 (0.0%) | 3 (0.0%) | 5 (0.0%) |
| Not performed | 4 (0.0%) | 6 (0.0%) | 10 (0.0%) |
| Waiting results | 27 (0.1%) | 7 (0.0%) | 34 (0.1%) |
| Missing | 25,666 (99.6%) | 19,468 (99.8%) | 45,134 (99.7%) |
| **Dengue IgM result** |  |  |  |
| Negative | 389 (1.5%) | 159 (0.8%) | 548 (1.2%) |
| Positive | 192 (0.7%) | 16 (0.1%) | 208 (0.5%) |
| Inconclusive | 38 (0.1%) | 7 (0.0%) | 45 (0.1%) |
| Not performed | 6,508 (25.2%) | 2,209 (11.3%) | 8,717 (19.2%) |
| Missing | 18,655 (72.4%) | 17,115 (87.7%) | 35,770 (79.0%) |
| **Dengue NS1 result** |  |  |  |
| Negative | 44 (0.2%) | 30 (0.2%) | 74 (0.2%) |
| Positive | 45 (0.2%) | 1 (0.0%) | 46 (0.1%) |
| Inconclusive | 7 (0.0%) | 0 (0%) | 7 (0.0%) |
| Not performed | 7,077 (27.4%) | 2,366 (12.1%) | 9,443 (20.9%) |
| Missing | 18,609 (72.2%) | 17,109 (87.7%) | 35,718 (78.9%) |
| **Dengue viral isolation result** |  |  |  |
| Negative | 5 (0.0%) | 2 (0.0%) | 7 (0.0%) |
| Positive | 3 (0.0%) | 1 (0.0%) | 4 (0.0%) |
| Inconclusive | 2 (0.0%) | 0 (0%) | 2 (0.0%) |
| Not performed | 7,167 (27.8%) | 2,395 (12.3%) | 9,562 (21.1%) |
| Missing | 18,605 (72.2%) | 17,108 (87.7%) | 35,713 (78.9%) |
| **Dengue RT-PCR result** |  |  |  |
| Negative | 3,591 (13.9%) | 436 (2.2%) | 4,027 (8.9%) |
| Positive | 25 (0.1%) | 4,441 (22.8%) | 4,466 (9.9%) |
| Inconclusive | 28 (0.1%) | 8 (0.0%) | 36 (0.1%) |
| Not performed | 22,079 (85.6%) | 14,564 (74.7%) | 36,643 (80.9%) |
| Missing | 59 (0.2%) | 57 (0.3%) | 116 (0.3%) |
| **Dengue histopathology result** |  |  |  |
| Dengue compatible | 0 (0%) | 0 (0%) | 0 (0%) |
| Dengue not compatible | 1 (0.0%) | 0 (0%) | 1 (0.0%) |
| Inconclusive | 2 (0.0%) | 0 (0%) | 2 (0.0%) |
| Not performed | 320 (1.2%) | 99 (0.5%) | 419 (0.9%) |
| Missing | 25,459 (98.7%) | 19,407 (99.5%) | 44,866 (99.1%) |
| **Dengue immunohistochemistry result** |  |  |  |
| Negative | 2 (0.0%) | 0 (0%) | 2 (0.0%) |
| Positive | 0 (0%) | 0 (0%) | 0 (0%) |
| Inconclusive | 0 (0%) | 0 (0%) | 0 (0%) |
| Not performed | 320 (1.2%) | 98 (0.5%) | 418 (0.9%) |
| Missing | 25,460 (98.8%) | 19,408 (99.5%) | 44,868 (99.1%) |
| **Chikungunya classification criteria** |  |  |  |
| Laboratory | 12,898 (50.0%) | 11,845 (60.7%) | 24,743 (54.6%) |
| Clinical-epidemiological | 12,740 (49.4%) | 7,585 (38.9%) | 20,325 (44.9%) |
| Not concluded | 19 (0.1%) | 24 (0.1%) | 43 (0.1%) |
| Missing | 125 (0.5%) | 52 (0.3%) | 177 (0.4%) |
| **Case outcome** |  |  |  |
| Recovered | 10,255 (39.8%) | 18,635 (95.5%) | 28,890 (63.8%) |
| Death (arbovirus) | 0 (0%) | 11 (0.1%) | 11 (0.0%) |
| Death (other causes) | 66 (0.3%) | 2 (0.0%) | 68 (0.2%) |
| Death (investigating) | 0 (0%) | 0 (0%) | 0 (0%) |
| Ignored | 14,089 (54.6%) | 713 (3.7%) | 14,802 (32.7%) |
| Missing | 1,372 (5.3%) | 145 (0.7%) | 1,517 (3.3%) |
| **Hospitalization** |  |  |  |
| No | 24,854 (96.4%) | 19,079 (97.8%) | 43,933 (97.0%) |
| Yes | 924 (3.6%) | 418 (2.1%) | 1,342 (3.0%) |
| Ignored | 4 (0.0%) | 7 (0.0%) | 11 (0.0%) |
| Missing | 0 (0%) | 2 (0.0%) | 2 (0.0%) |
| **Disease timing at presentation** |  |  |  |
| Acute | 5,010 (19.4%) | 16,370 (83.9%) | 21,380 (47.2%) |
| Chronic | 331 (1.3%) | 758 (3.9%) | 1,089 (2.4%) |
| Missing | 20,441 (79.3%) | 2,378 (12.2%) | 22,819 (50.4%) |
| IQR = interquartile range; SD = Standard deviation; CNS number = cartão nacional do SUS (used as notification ID); PRNT = plaque reduction neutralization test: RT-PCR = reverse transcription polymerase chain reaction. | | | |

**Table S 3** Zika and Oropouche dataset variables from the original notification form (including epidemiological, clinical at presentations, and disease progressions) counts and frequencies by case diagnostic investigation outcome.

|  | **Discarded (N=18500)** | **Zika (N=1080)** | **Oropouche (N=12173)** | **Overall (N=31753)** |
| --- | --- | --- | --- | --- |
| **Valid CNS number** |  |  |  |  |
| Yes | 18,413 (99.5%) | 1,078 (99.8%) | 12,168 (100.0%) | 31,659 (99.7%) |
| No | 87 (0.5%) | 2 (0.2%) | 5 (0.0%) | 94 (0.3%) |
| **Age at notification** |  |  |  |  |
| Mean (SD) | 35.3 (18.5) | 39.7 (18.2) | 42.0 (18.2) | 38.0 (18.7) |
| Median [Min, Max] | 33.0 [0, 96.0] | 40.0 [0, 93.0] | 42.0 [0, 100] | 37.0 [0, 100] |
| **Age at notification** |  |  |  |  |
| [0,1] | 222 (1.2%) | 8 (0.7%) | 12 (0.1%) | 242 (0.8%) |
| (1,12] | 1,545 (8.4%) | 68 (6.3%) | 462 (3.8%) | 2,075 (6.5%) |
| (12,18] | 1,810 (9.8%) | 91 (8.4%) | 787 (6.5%) | 2,688 (8.5%) |
| (18,25] | 2,888 (15.6%) | 100 (9.3%) | 1,304 (10.7%) | 4,292 (13.5%) |
| (25,35] | 3,533 (19.1%) | 170 (15.7%) | 2,103 (17.3%) | 5,806 (18.3%) |
| (35,45] | 3,254 (17.6%) | 229 (21.2%) | 2,405 (19.8%) | 5,888 (18.5%) |
| (45,55] | 2,306 (12.5%) | 178 (16.5%) | 2,038 (16.7%) | 4,522 (14.2%) |
| (55,65] | 1,638 (8.9%) | 154 (14.3%) | 1,684 (13.8%) | 3,476 (10.9%) |
| (65,75] | 945 (5.1%) | 63 (5.8%) | 963 (7.9%) | 1,971 (6.2%) |
| (75,85] | 289 (1.6%) | 16 (1.5%) | 340 (2.8%) | 645 (2.0%) |
| (85,95] | 68 (0.4%) | 3 (0.3%) | 72 (0.6%) | 143 (0.5%) |
| Missing | 2 (0.0%) | 0 (0%) | 3 (0.0%) | 5 (0.0%) |
| **Sex at notification** |  |  |  |  |
| Female | 10,842 (58.6%) | 614 (56.9%) | 5,649 (46.4%) | 17,105 (53.9%) |
| Male | 7,658 (41.4%) | 466 (43.1%) | 6,524 (53.6%) | 14,648 (46.1%) |
| Ignored | 0 (0%) | 0 (0%) | 0 (0%) | 0 (0%) |
| **Pregnancy** |  |  |  |  |
| 1st trimester | 289 (1.6%) | 9 (0.8%) | 55 (0.5%) | 353 (1.1%) |
| 2nd trimester | 368 (2.0%) | 11 (1.0%) | 93 (0.8%) | 472 (1.5%) |
| 3rd trimester | 342 (1.8%) | 13 (1.2%) | 98 (0.8%) | 453 (1.4%) |
| Gestational age unknown | 79 (0.4%) | 10 (0.9%) | 0 (0%) | 89 (0.3%) |
| No | 7,608 (41.1%) | 505 (46.8%) | 4,838 (39.7%) | 12,951 (40.8%) |
| Not applicable | 8,748 (47.3%) | 493 (45.6%) | 6,731 (55.3%) | 15,972 (50.3%) |
| Ignored/Unknown | 1,066 (5.8%) | 39 (3.6%) | 358 (2.9%) | 1,463 (4.6%) |
| **Race or skin color** |  |  |  |  |
| White | 5,413 (29.3%) | 453 (41.9%) | 6,846 (56.2%) | 12,712 (40.0%) |
| Black | 812 (4.4%) | 49 (4.5%) | 660 (5.4%) | 1,521 (4.8%) |
| Yellow | 1,934 (10.5%) | 116 (10.7%) | 1,428 (11.7%) | 3,478 (11.0%) |
| Brown | 8,623 (46.6%) | 358 (33.1%) | 3,232 (26.6%) | 12,213 (38.5%) |
| Indigenous | 27 (0.1%) | 0 (0%) | 7 (0.1%) | 34 (0.1%) |
| Ignored/Unknown | 1,691 (9.1%) | 104 (9.6%) | 0 (0%) | 1,795 (5.7%) |
| **Indigenous ethnicity** |  |  |  |  |
| Tupiniquim | 10 (0.1%) | 0 (0%) | 1 (0.0%) | 11 (0.0%) |
| Guarani | 1 (0.0%) | 0 (0%) | 0 (0%) | 1 (0.0%) |
| Ignored/Unknown | 16 (0.1%) | 0 (0%) | 6 (0.0%) | 22 (0.1%) |
| Missing | 18,473 (99.9%) | 1,080 (100%) | 12,166 (99.9%) | 31,719 (99.9%) |
| **Person with special needs** |  |  |  |  |
| Yes | 10 (0.1%) | 1 (0.1%) | 0 (0%) | 11 (0.0%) |
| No | 18,490 (99.9%) | 1,079 (99.9%) | 12,173 (100%) | 31,742 (100.0%) |
| **Homeless** |  |  |  |  |
| Yes | 3 (0.0%) | 0 (0%) | 0 (0%) | 3 (0.0%) |
| No | 18,497 (100.0%) | 1,080 (100%) | 12,173 (100%) | 31,750 (100.0%) |
| **Education** |  |  |  |  |
| Illiterate | 101 (0.5%) | 7 (0.6%) | 101 (0.8%) | 209 (0.7%) |
| Elementary School Incomplete | 849 (4.6%) | 69 (6.4%) | 831 (6.8%) | 1,749 (5.5%) |
| Elementary School Complete | 459 (2.5%) | 34 (3.1%) | 706 (5.8%) | 1,199 (3.8%) |
| Middle School Incomplete | 1,681 (9.1%) | 121 (11.2%) | 1,336 (11.0%) | 3,138 (9.9%) |
| Middle School Complete | 840 (4.5%) | 59 (5.5%) | 736 (6.0%) | 1,635 (5.1%) |
| High School Incomplete | 1,097 (5.9%) | 68 (6.3%) | 832 (6.8%) | 1,997 (6.3%) |
| High School Complete | 3,753 (20.3%) | 237 (21.9%) | 2,883 (23.7%) | 6,873 (21.6%) |
| Higher Education Incomplete | 324 (1.8%) | 35 (3.2%) | 161 (1.3%) | 520 (1.6%) |
| Higher Education Complete | 1,147 (6.2%) | 125 (11.6%) | 744 (6.1%) | 2,016 (6.3%) |
| Ignored/Unknown | 7,433 (40.2%) | 275 (25.5%) | 3,709 (30.5%) | 11,417 (36.0%) |
| Not applicable | 816 (4.4%) | 50 (4.6%) | 134 (1.1%) | 1,000 (3.1%) |
| **Home address** |  |  |  |  |
| Urban | 12,776 (69.1%) | 744 (68.9%) | 5,833 (47.9%) | 19,353 (60.9%) |
| Rural | 3,083 (16.7%) | 204 (18.9%) | 5,475 (45.0%) | 8,762 (27.6%) |
| Peri-urban | 144 (0.8%) | 9 (0.8%) | 68 (0.6%) | 221 (0.7%) |
| Unknown | 2,497 (13.5%) | 123 (11.4%) | 794 (6.5%) | 3,414 (10.8%) |
| Missing | 0 (0%) | 0 (0%) | 3 (0.0%) | 3 (0.0%) |
| **Autochthonous case** |  |  |  |  |
| No | 43 (0.2%) | 6 (0.6%) | 311 (2.6%) | 360 (1.1%) |
| Yes | 18,345 (99.2%) | 1,061 (98.2%) | 11,705 (96.2%) | 31,111 (98.0%) |
| Unknown | 112 (0.6%) | 13 (1.2%) | 157 (1.3%) | 282 (0.9%) |
| **Case classification criteria** |  |  |  |  |
| Laboratory | 6,410 (34.6%) | 860 (79.6%) | 12,006 (98.6%) | 19,276 (60.7%) |
| Clinical-epidemiological | 11,959 (64.6%) | 212 (19.6%) | 0 (0%) | 12,171 (38.3%) |
| Missing | 131 (0.7%) | 8 (0.7%) | 167 (1.4%) | 306 (1.0%) |
| **Case outcome** |  |  |  |  |
| Recovered | 6,702 (36.2%) | 995 (92.1%) | 10,905 (89.6%) | 18,602 (58.6%) |
| Death (arbovirus) | 0 (0%) | 0 (0%) | 2 (0.0%) | 2 (0.0%) |
| Death (other causes) | 47 (0.3%) | 0 (0%) | 8 (0.1%) | 55 (0.2%) |
| Ignored | 9,408 (50.9%) | 47 (4.4%) | 284 (2.3%) | 9,739 (30.7%) |
| Missing | 2,343 (12.7%) | 38 (3.5%) | 974 (8.0%) | 3,355 (10.6%) |
| IQR = interquartile range; SD = Standard deviation; CNS number = cartão nacional do SUS (used as notification ID). | | | | |

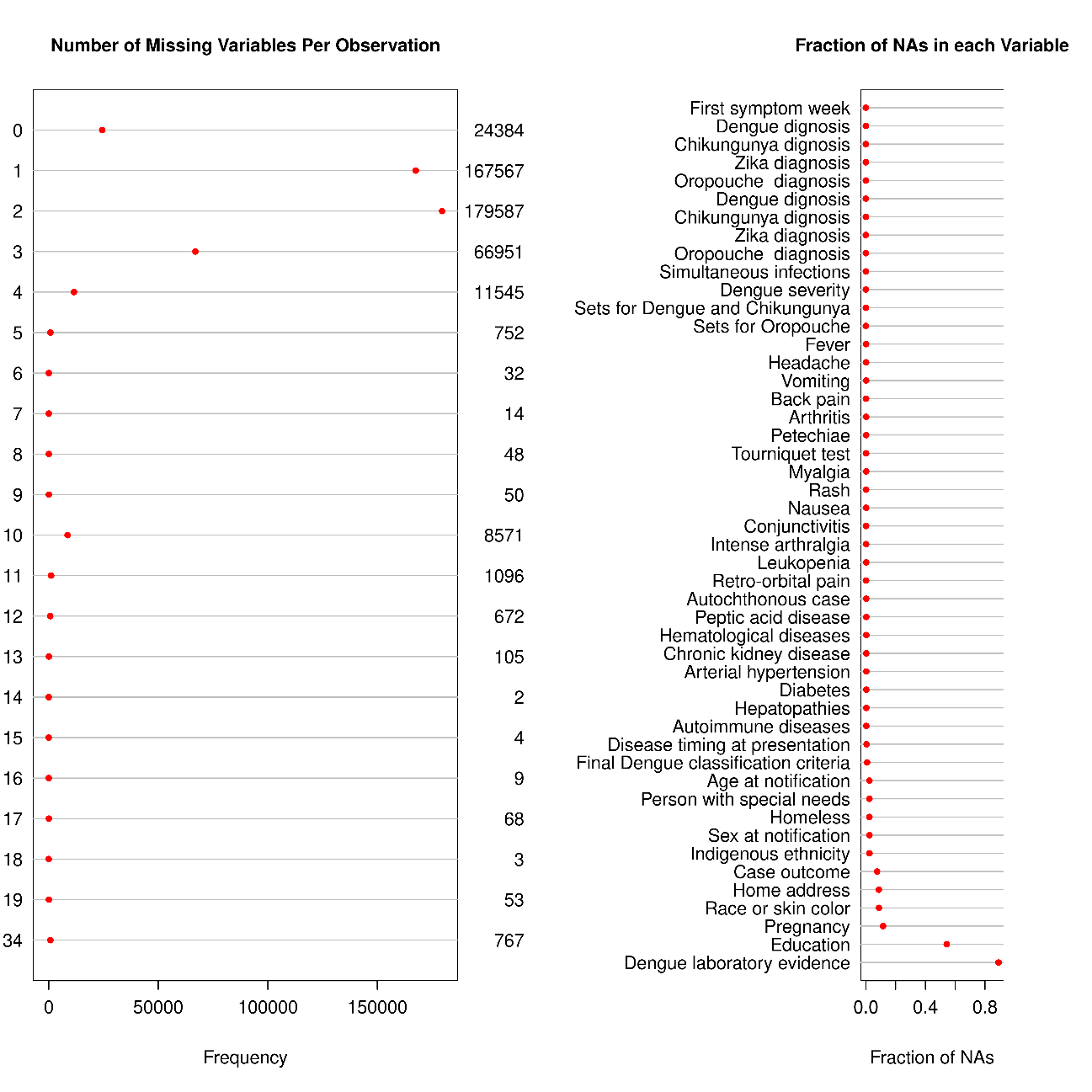

**Figure S 3:** Missing pattern per variable in the whole unified clean dataset.

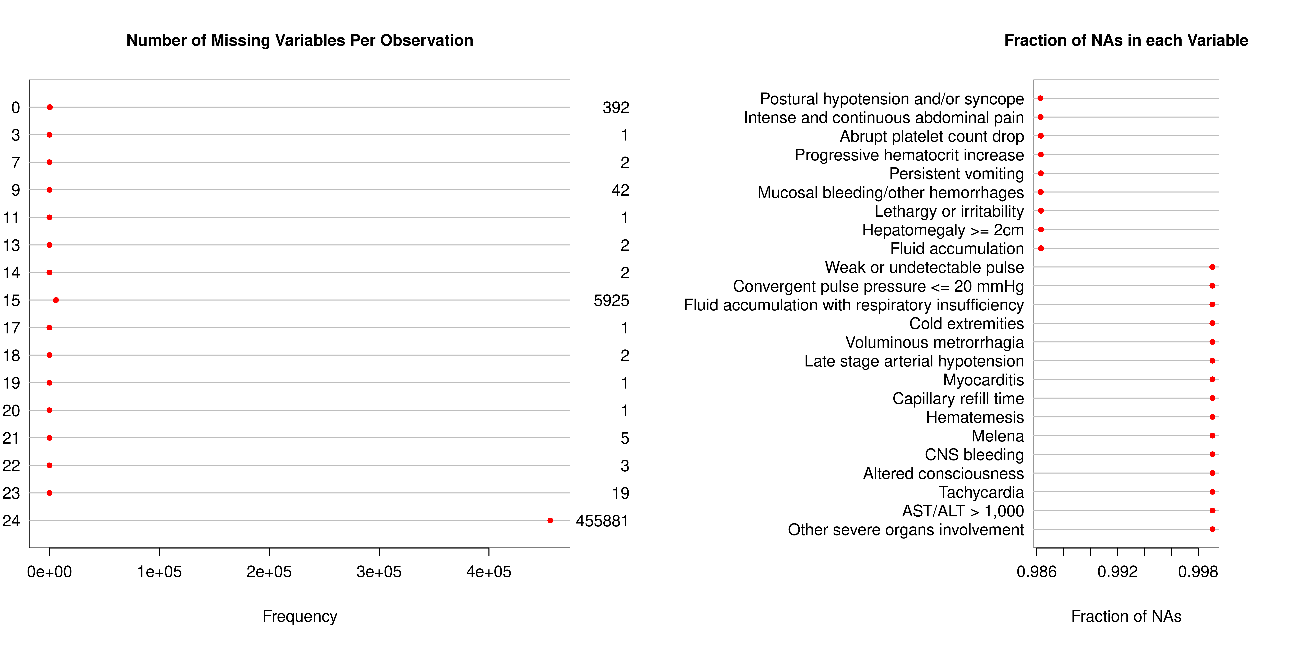

**Figure S 4:** Missing pattern per variables of prognosis chunk, in the whole unified clean dataset.

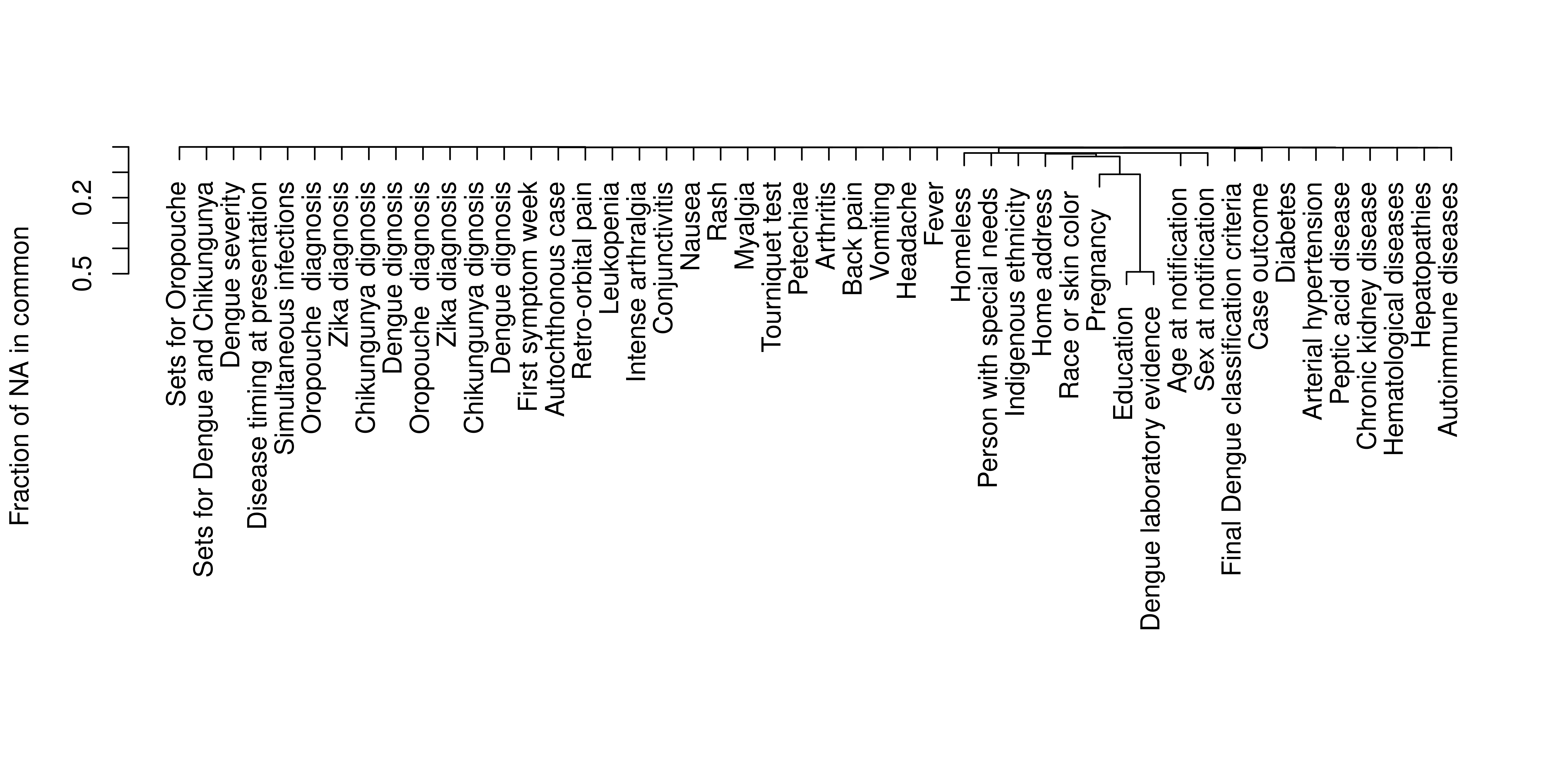

**Figure S 5:** Missing pattern (fractions) in common in the whole unified clean dataset.

**Table S 4** Final dataset variables (including epidemiological, clinical at presentations and disease progressions) counts and frequencies by dengue diagnostic investigation outcome in each dataset split.

|  | **No** | | **Yes** | | **Overall** | |
| --- | --- | --- | --- | --- | --- | --- |
|  | **Train (N=133658)** | **Test (N=66931)** | **Train (N=174357)** | **Test (N=87334)** | **Train (N=308015)** | **Test (N=154265)** |
| **Age at notification** |  |  |  |  |  |  |
| Mean (SD) | 34.9 (19.7) | 34.8 (19.6) | 35.5 (18.6) | 35.3 (18.6) | 35.2 (19.1) | 35.1 (19.0) |
| Median [Min, Max] | 33.0 [0, 106] | 33.0 [0, 104] | 33.0 [0, 104] | 33.0 [0, 102] | 33.0 [0, 106] | 33.0 [0, 104] |
| Missing | 6,949 (5.2%) | 3,557 (5.3%) | 359 (0.2%) | 169 (0.2%) | 7,308 (2.4%) | 3,726 (2.4%) |
| **Age at notification** |  |  |  |  |  |  |
| [0,1] | 2,249 (1.7%) | 1,171 (1.7%) | 1,244 (0.7%) | 656 (0.8%) | 3,493 (1.1%) | 1,827 (1.2%) |
| (1,12] | 14,411 (10.8%) | 7,098 (10.6%) | 15,480 (8.9%) | 7,968 (9.1%) | 29,891 (9.7%) | 15,066 (9.8%) |
| (12,18] | 11,639 (8.7%) | 5,979 (8.9%) | 16,719 (9.6%) | 8,322 (9.5%) | 28,358 (9.2%) | 14,301 (9.3%) |
| (18,25] | 17,584 (13.2%) | 8,797 (13.1%) | 27,399 (15.7%) | 13,908 (15.9%) | 44,983 (14.6%) | 22,705 (14.7%) |
| (25,35] | 22,756 (17.0%) | 11,470 (17.1%) | 32,683 (18.7%) | 16,411 (18.8%) | 55,439 (18.0%) | 27,881 (18.1%) |
| (35,45] | 21,139 (15.8%) | 10,647 (15.9%) | 30,063 (17.2%) | 14,962 (17.1%) | 51,202 (16.6%) | 25,609 (16.6%) |
| (45,55] | 15,816 (11.8%) | 7,795 (11.6%) | 22,048 (12.6%) | 10,947 (12.5%) | 37,864 (12.3%) | 18,742 (12.1%) |
| (55,65] | 11,172 (8.4%) | 5,623 (8.4%) | 15,894 (9.1%) | 7,772 (8.9%) | 27,066 (8.8%) | 13,395 (8.7%) |
| (65,75] | 6,633 (5.0%) | 3,149 (4.7%) | 8,724 (5.0%) | 4,368 (5.0%) | 15,357 (5.0%) | 7,517 (4.9%) |
| (75,85] | 2,604 (1.9%) | 1,286 (1.9%) | 3,046 (1.7%) | 1,482 (1.7%) | 5,650 (1.8%) | 2,768 (1.8%) |
| (85,95] | 676 (0.5%) | 336 (0.5%) | 668 (0.4%) | 347 (0.4%) | 1,344 (0.4%) | 683 (0.4%) |
| Missing | 6,979 (5.2%) | 3,580 (5.3%) | 389 (0.2%) | 191 (0.2%) | 7,368 (2.4%) | 3,771 (2.4%) |
| **Sex at notification** |  |  |  |  |  |  |
| Female | 67,774 (50.7%) | 33,967 (50.7%) | 94,998 (54.5%) | 47,644 (54.6%) | 162,772 (52.8%) | 81,611 (52.9%) |
| Male | 58,927 (44.1%) | 29,405 (43.9%) | 78,996 (45.3%) | 39,518 (45.2%) | 137,923 (44.8%) | 68,923 (44.7%) |
| Ignored | 8 (0.0%) | 2 (0.0%) | 4 (0.0%) | 3 (0.0%) | 12 (0.0%) | 5 (0.0%) |
| Missing | 6,949 (5.2%) | 3,557 (5.3%) | 359 (0.2%) | 169 (0.2%) | 7,308 (2.4%) | 3,726 (2.4%) |
| **Pregnancy** |  |  |  |  |  |  |
| No | 116,821 (87.4%) | 58,399 (87.3%) | 152,424 (87.4%) | 76,135 (87.2%) | 269,245 (87.4%) | 134,534 (87.2%) |
| Yes | 1,671 (1.3%) | 860 (1.3%) | 1,680 (1.0%) | 895 (1.0%) | 3,351 (1.1%) | 1,755 (1.1%) |
| Ignored/Unknown | 8,217 (6.1%) | 4,115 (6.1%) | 19,894 (11.4%) | 10,135 (11.6%) | 28,111 (9.1%) | 14,250 (9.2%) |
| Missing | 6,949 (5.2%) | 3,557 (5.3%) | 359 (0.2%) | 169 (0.2%) | 7,308 (2.4%) | 3,726 (2.4%) |
| **Race or skin color** |  |  |  |  |  |  |
| White | 46,943 (35.1%) | 23,414 (35.0%) | 45,687 (26.2%) | 22,981 (26.3%) | 92,630 (30.1%) | 46,395 (30.1%) |
| Black | 63,587 (47.6%) | 31,887 (47.6%) | 97,045 (55.7%) | 48,560 (55.6%) | 160,632 (52.2%) | 80,447 (52.1%) |
| Yellow | 11,908 (8.9%) | 5,934 (8.9%) | 15,576 (8.9%) | 7,823 (9.0%) | 27,484 (8.9%) | 13,757 (8.9%) |
| Indigenous | 146 (0.1%) | 73 (0.1%) | 135 (0.1%) | 75 (0.1%) | 281 (0.1%) | 148 (0.1%) |
| Ignored/Unknown | 4,125 (3.1%) | 2,066 (3.1%) | 15,555 (8.9%) | 7,726 (8.8%) | 19,680 (6.4%) | 9,792 (6.3%) |
| Missing | 6,949 (5.2%) | 3,557 (5.3%) | 359 (0.2%) | 169 (0.2%) | 7,308 (2.4%) | 3,726 (2.4%) |
| **Indigenous ethnicity** |  |  |  |  |  |  |
| Tupiniquim | 84 (0.1%) | 34 (0.1%) | 29 (0.0%) | 15 (0.0%) | 113 (0.0%) | 49 (0.0%) |
| Guarani | 11 (0.0%) | 9 (0.0%) | 14 (0.0%) | 9 (0.0%) | 25 (0.0%) | 18 (0.0%) |
| Ignored/Unknown | 51 (0.0%) | 30 (0.0%) | 92 (0.1%) | 51 (0.1%) | 143 (0.0%) | 81 (0.1%) |
| Not applicable | 126,563 (94.7%) | 63,301 (94.6%) | 173,863 (99.7%) | 87,090 (99.7%) | 300,426 (97.5%) | 150,391 (97.5%) |
| Missing | 6,949 (5.2%) | 3,557 (5.3%) | 359 (0.2%) | 169 (0.2%) | 7,308 (2.4%) | 3,726 (2.4%) |
| **Person with special needs** |  |  |  |  |  |  |
| Yes | 71 (0.1%) | 20 (0.0%) | 236 (0.1%) | 138 (0.2%) | 307 (0.1%) | 158 (0.1%) |
| No | 126,638 (94.7%) | 63,354 (94.7%) | 173,762 (99.7%) | 87,027 (99.6%) | 300,400 (97.5%) | 150,381 (97.5%) |
| Missing | 6,949 (5.2%) | 3,557 (5.3%) | 359 (0.2%) | 169 (0.2%) | 7,308 (2.4%) | 3,726 (2.4%) |
| **Homeless** |  |  |  |  |  |  |
| Yes | 18 (0.0%) | 6 (0.0%) | 84 (0.0%) | 40 (0.0%) | 102 (0.0%) | 46 (0.0%) |
| No | 126,691 (94.8%) | 63,368 (94.7%) | 173,914 (99.7%) | 87,125 (99.8%) | 300,605 (97.6%) | 150,493 (97.6%) |
| Missing | 6,949 (5.2%) | 3,557 (5.3%) | 359 (0.2%) | 169 (0.2%) | 7,308 (2.4%) | 3,726 (2.4%) |
| **Education** |  |  |  |  |  |  |
| Illiterate | 7,393 (5.5%) | 3,663 (5.5%) | 6,673 (3.8%) | 3,233 (3.7%) | 14,066 (4.6%) | 6,896 (4.5%) |
| Elementary School Complete | 16,158 (12.1%) | 8,004 (12.0%) | 14,983 (8.6%) | 7,540 (8.6%) | 31,141 (10.1%) | 15,544 (10.1%) |
| Middle School Complete | 14,058 (10.5%) | 6,966 (10.4%) | 14,346 (8.2%) | 7,186 (8.2%) | 28,404 (9.2%) | 14,152 (9.2%) |
| High School Complete | 23,577 (17.6%) | 12,022 (18.0%) | 29,146 (16.7%) | 14,407 (16.5%) | 52,723 (17.1%) | 26,429 (17.1%) |
| Higher Education Complete | 6,554 (4.9%) | 3,172 (4.7%) | 8,067 (4.6%) | 4,097 (4.7%) | 14,621 (4.7%) | 7,269 (4.7%) |
| Ignored/Unknown | 58,969 (44.1%) | 29,547 (44.1%) | 100,783 (57.8%) | 50,702 (58.1%) | 159,752 (51.9%) | 80,249 (52.0%) |
| Missing | 6,949 (5.2%) | 3,557 (5.3%) | 359 (0.2%) | 169 (0.2%) | 7,308 (2.4%) | 3,726 (2.4%) |
| **Home address** |  |  |  |  |  |  |
| Urban | 85,392 (63.9%) | 42,537 (63.6%) | 149,354 (85.7%) | 74,762 (85.6%) | 234,746 (76.2%) | 117,299 (76.0%) |
| Rural | 28,944 (21.7%) | 14,626 (21.9%) | 16,659 (9.6%) | 8,319 (9.5%) | 45,603 (14.8%) | 22,945 (14.9%) |
| Peri-urban | 308 (0.2%) | 159 (0.2%) | 629 (0.4%) | 333 (0.4%) | 937 (0.3%) | 492 (0.3%) |
| Unknown | 12,065 (9.0%) | 6,052 (9.0%) | 7,353 (4.2%) | 3,748 (4.3%) | 19,418 (6.3%) | 9,800 (6.4%) |
| Missing | 6,949 (5.2%) | 3,557 (5.3%) | 362 (0.2%) | 172 (0.2%) | 7,311 (2.4%) | 3,729 (2.4%) |
| **Autochthonous case** |  |  |  |  |  |  |
| No | 283 (0.2%) | 114 (0.2%) | 231 (0.1%) | 104 (0.1%) | 514 (0.2%) | 218 (0.1%) |
| Yes | 132,529 (99.2%) | 66,387 (99.2%) | 173,898 (99.7%) | 87,115 (99.7%) | 306,427 (99.5%) | 153,502 (99.5%) |
| Unknown | 132 (0.1%) | 82 (0.1%) | 204 (0.1%) | 102 (0.1%) | 336 (0.1%) | 184 (0.1%) |
| Missing | 714 (0.5%) | 348 (0.5%) | 24 (0.0%) | 13 (0.0%) | 738 (0.2%) | 361 (0.2%) |
| **Fever** |  |  |  |  |  |  |
| No | 41,131 (30.8%) | 20,648 (30.8%) | 30,261 (17.4%) | 15,120 (17.3%) | 71,392 (23.2%) | 35,768 (23.2%) |
| Yes | 92,017 (68.8%) | 46,026 (68.8%) | 144,096 (82.6%) | 72,214 (82.7%) | 236,113 (76.7%) | 118,240 (76.6%) |
| Missing | 510 (0.4%) | 257 (0.4%) | 0 (0%) | 0 (0%) | 510 (0.2%) | 257 (0.2%) |
| **Headache** |  |  |  |  |  |  |
| No | 39,689 (29.7%) | 19,752 (29.5%) | 34,148 (19.6%) | 17,150 (19.6%) | 73,837 (24.0%) | 36,902 (23.9%) |
| Yes | 93,459 (69.9%) | 46,922 (70.1%) | 140,209 (80.4%) | 70,184 (80.4%) | 233,668 (75.9%) | 117,106 (75.9%) |
| Missing | 510 (0.4%) | 257 (0.4%) | 0 (0%) | 0 (0%) | 510 (0.2%) | 257 (0.2%) |
| **Vomiting** |  |  |  |  |  |  |
| No | 106,267 (79.5%) | 53,207 (79.5%) | 137,472 (78.8%) | 68,996 (79.0%) | 243,739 (79.1%) | 122,203 (79.2%) |
| Yes | 26,881 (20.1%) | 13,467 (20.1%) | 36,885 (21.2%) | 18,338 (21.0%) | 63,766 (20.7%) | 31,805 (20.6%) |
| Missing | 510 (0.4%) | 257 (0.4%) | 0 (0%) | 0 (0%) | 510 (0.2%) | 257 (0.2%) |
| **Back pain** |  |  |  |  |  |  |
| No | 106,291 (79.5%) | 53,208 (79.5%) | 136,230 (78.1%) | 68,278 (78.2%) | 242,521 (78.7%) | 121,486 (78.8%) |
| Yes | 26,857 (20.1%) | 13,466 (20.1%) | 38,127 (21.9%) | 19,056 (21.8%) | 64,984 (21.1%) | 32,522 (21.1%) |
| Missing | 510 (0.4%) | 257 (0.4%) | 0 (0%) | 0 (0%) | 510 (0.2%) | 257 (0.2%) |
| **Arthritis** |  |  |  |  |  |  |
| No | 124,040 (92.8%) | 62,075 (92.7%) | 162,492 (93.2%) | 81,577 (93.4%) | 286,532 (93.0%) | 143,652 (93.1%) |
| Yes | 9,108 (6.8%) | 4,599 (6.9%) | 11,865 (6.8%) | 5,757 (6.6%) | 20,973 (6.8%) | 10,356 (6.7%) |
| Missing | 510 (0.4%) | 257 (0.4%) | 0 (0%) | 0 (0%) | 510 (0.2%) | 257 (0.2%) |
| **Petechiae** |  |  |  |  |  |  |
| No | 128,227 (95.9%) | 64,209 (95.9%) | 165,044 (94.7%) | 82,870 (94.9%) | 293,271 (95.2%) | 147,079 (95.3%) |
| Yes | 4,921 (3.7%) | 2,465 (3.7%) | 9,313 (5.3%) | 4,464 (5.1%) | 14,234 (4.6%) | 6,929 (4.5%) |
| Missing | 510 (0.4%) | 257 (0.4%) | 0 (0%) | 0 (0%) | 510 (0.2%) | 257 (0.2%) |
| **Tourniquet test** |  |  |  |  |  |  |
| No | 132,211 (98.9%) | 66,255 (99.0%) | 171,372 (98.3%) | 85,916 (98.4%) | 303,583 (98.6%) | 152,171 (98.6%) |
| Yes | 937 (0.7%) | 419 (0.6%) | 2,985 (1.7%) | 1,418 (1.6%) | 3,922 (1.3%) | 1,837 (1.2%) |
| Missing | 510 (0.4%) | 257 (0.4%) | 0 (0%) | 0 (0%) | 510 (0.2%) | 257 (0.2%) |
| **Myalgia** |  |  |  |  |  |  |
| No | 46,106 (34.5%) | 23,146 (34.6%) | 38,681 (22.2%) | 19,433 (22.3%) | 84,787 (27.5%) | 42,579 (27.6%) |
| Yes | 87,042 (65.1%) | 43,528 (65.0%) | 135,676 (77.8%) | 67,901 (77.7%) | 222,718 (72.3%) | 111,429 (72.2%) |
| Missing | 510 (0.4%) | 257 (0.4%) | 0 (0%) | 0 (0%) | 510 (0.2%) | 257 (0.2%) |
| **Rash** |  |  |  |  |  |  |
| No | 128,669 (96.3%) | 64,412 (96.2%) | 165,407 (94.9%) | 82,832 (94.8%) | 294,076 (95.5%) | 147,244 (95.4%) |
| Yes | 4,479 (3.4%) | 2,262 (3.4%) | 8,950 (5.1%) | 4,502 (5.2%) | 13,429 (4.4%) | 6,764 (4.4%) |
| Missing | 510 (0.4%) | 257 (0.4%) | 0 (0%) | 0 (0%) | 510 (0.2%) | 257 (0.2%) |
| **Nausea** |  |  |  |  |  |  |
| No | 89,636 (67.1%) | 45,079 (67.4%) | 110,662 (63.5%) | 55,349 (63.4%) | 200,298 (65.0%) | 100,428 (65.1%) |
| Yes | 43,512 (32.6%) | 21,595 (32.3%) | 63,695 (36.5%) | 31,985 (36.6%) | 107,207 (34.8%) | 53,580 (34.7%) |
| Missing | 510 (0.4%) | 257 (0.4%) | 0 (0%) | 0 (0%) | 510 (0.2%) | 257 (0.2%) |
| **Conjunctivitis** |  |  |  |  |  |  |
| No | 132,136 (98.9%) | 66,207 (98.9%) | 172,262 (98.8%) | 86,297 (98.8%) | 304,398 (98.8%) | 152,504 (98.9%) |
| Yes | 1,012 (0.8%) | 467 (0.7%) | 2,095 (1.2%) | 1,037 (1.2%) | 3,107 (1.0%) | 1,504 (1.0%) |
| Missing | 510 (0.4%) | 257 (0.4%) | 0 (0%) | 0 (0%) | 510 (0.2%) | 257 (0.2%) |
| **Intense arthralgia** |  |  |  |  |  |  |
| No | 109,651 (82.0%) | 54,830 (81.9%) | 135,271 (77.6%) | 67,904 (77.8%) | 244,922 (79.5%) | 122,734 (79.6%) |
| Yes | 23,497 (17.6%) | 11,844 (17.7%) | 39,086 (22.4%) | 19,430 (22.2%) | 62,583 (20.3%) | 31,274 (20.3%) |
| Missing | 510 (0.4%) | 257 (0.4%) | 0 (0%) | 0 (0%) | 510 (0.2%) | 257 (0.2%) |
| **Leukopenia** |  |  |  |  |  |  |
| No | 131,636 (98.5%) | 65,879 (98.4%) | 170,284 (97.7%) | 85,315 (97.7%) | 301,920 (98.0%) | 151,194 (98.0%) |
| Yes | 1,512 (1.1%) | 795 (1.2%) | 4,073 (2.3%) | 2,019 (2.3%) | 5,585 (1.8%) | 2,814 (1.8%) |
| Missing | 510 (0.4%) | 257 (0.4%) | 0 (0%) | 0 (0%) | 510 (0.2%) | 257 (0.2%) |
| **Retro-orbital pain** |  |  |  |  |  |  |
| No | 99,975 (74.8%) | 49,991 (74.7%) | 114,260 (65.5%) | 57,169 (65.5%) | 214,235 (69.6%) | 107,160 (69.5%) |
| Yes | 33,173 (24.8%) | 16,683 (24.9%) | 60,097 (34.5%) | 30,165 (34.5%) | 93,270 (30.3%) | 46,848 (30.4%) |
| Missing | 510 (0.4%) | 257 (0.4%) | 0 (0%) | 0 (0%) | 510 (0.2%) | 257 (0.2%) |
| **Diabetes** |  |  |  |  |  |  |
| No | 129,532 (96.9%) | 64,844 (96.9%) | 168,801 (96.8%) | 84,616 (96.9%) | 298,333 (96.9%) | 149,460 (96.9%) |
| Yes | 3,380 (2.5%) | 1,722 (2.6%) | 5,389 (3.1%) | 2,654 (3.0%) | 8,769 (2.8%) | 4,376 (2.8%) |
| Ignored | 236 (0.2%) | 108 (0.2%) | 167 (0.1%) | 64 (0.1%) | 403 (0.1%) | 172 (0.1%) |
| Missing | 510 (0.4%) | 257 (0.4%) | 0 (0%) | 0 (0%) | 510 (0.2%) | 257 (0.2%) |
| **Hepatopathies** |  |  |  |  |  |  |
| No | 132,782 (99.3%) | 66,500 (99.4%) | 173,973 (99.8%) | 87,146 (99.8%) | 306,755 (99.6%) | 153,646 (99.6%) |
| Yes | 130 (0.1%) | 62 (0.1%) | 220 (0.1%) | 123 (0.1%) | 350 (0.1%) | 185 (0.1%) |
| Ignored | 236 (0.2%) | 112 (0.2%) | 164 (0.1%) | 65 (0.1%) | 400 (0.1%) | 177 (0.1%) |
| Missing | 510 (0.4%) | 257 (0.4%) | 0 (0%) | 0 (0%) | 510 (0.2%) | 257 (0.2%) |
| **Arterial hypertension** |  |  |  |  |  |  |
| No | 123,749 (92.6%) | 61,986 (92.6%) | 160,476 (92.0%) | 80,562 (92.2%) | 284,225 (92.3%) | 142,548 (92.4%) |
| Yes | 9,166 (6.9%) | 4,581 (6.8%) | 13,716 (7.9%) | 6,707 (7.7%) | 22,882 (7.4%) | 11,288 (7.3%) |
| Ignored | 233 (0.2%) | 107 (0.2%) | 165 (0.1%) | 65 (0.1%) | 398 (0.1%) | 172 (0.1%) |
| Missing | 510 (0.4%) | 257 (0.4%) | 0 (0%) | 0 (0%) | 510 (0.2%) | 257 (0.2%) |
| **Autoimmune diseases** |  |  |  |  |  |  |
| No | 132,548 (99.2%) | 66,376 (99.2%) | 173,590 (99.6%) | 86,978 (99.6%) | 306,138 (99.4%) | 153,354 (99.4%) |
| Yes | 360 (0.3%) | 189 (0.3%) | 600 (0.3%) | 284 (0.3%) | 960 (0.3%) | 473 (0.3%) |
| Ignored | 240 (0.2%) | 109 (0.2%) | 167 (0.1%) | 72 (0.1%) | 407 (0.1%) | 181 (0.1%) |
| Missing | 510 (0.4%) | 257 (0.4%) | 0 (0%) | 0 (0%) | 510 (0.2%) | 257 (0.2%) |
| **Hematological diseases** |  |  |  |  |  |  |
| No | 132,817 (99.4%) | 66,504 (99.4%) | 174,010 (99.8%) | 87,172 (99.8%) | 306,827 (99.6%) | 153,676 (99.6%) |
| Yes | 100 (0.1%) | 61 (0.1%) | 186 (0.1%) | 99 (0.1%) | 286 (0.1%) | 160 (0.1%) |
| Ignored | 231 (0.2%) | 109 (0.2%) | 161 (0.1%) | 63 (0.1%) | 392 (0.1%) | 172 (0.1%) |
| Missing | 510 (0.4%) | 257 (0.4%) | 0 (0%) | 0 (0%) | 510 (0.2%) | 257 (0.2%) |
| **Chronic kidney disease** |  |  |  |  |  |  |
| No | 132,721 (99.3%) | 66,468 (99.3%) | 173,969 (99.8%) | 87,164 (99.8%) | 306,690 (99.6%) | 153,632 (99.6%) |
| Yes | 197 (0.1%) | 97 (0.1%) | 228 (0.1%) | 105 (0.1%) | 425 (0.1%) | 202 (0.1%) |
| Ignored | 230 (0.2%) | 109 (0.2%) | 160 (0.1%) | 65 (0.1%) | 390 (0.1%) | 174 (0.1%) |
| Missing | 510 (0.4%) | 257 (0.4%) | 0 (0%) | 0 (0%) | 510 (0.2%) | 257 (0.2%) |
| **Peptic acid disease** |  |  |  |  |  |  |
| No | 132,875 (99.4%) | 66,538 (99.4%) | 174,081 (99.8%) | 87,191 (99.8%) | 306,956 (99.7%) | 153,729 (99.7%) |
| Yes | 39 (0.0%) | 28 (0.0%) | 121 (0.1%) | 79 (0.1%) | 160 (0.1%) | 107 (0.1%) |
| Ignored | 234 (0.2%) | 108 (0.2%) | 155 (0.1%) | 64 (0.1%) | 389 (0.1%) | 172 (0.1%) |
| Missing | 510 (0.4%) | 257 (0.4%) | 0 (0%) | 0 (0%) | 510 (0.2%) | 257 (0.2%) |
| **Final Dengue classification criteria** |  |  |  |  |  |  |
| Laboratory | 69,005 (51.6%) | 34,525 (51.6%) | 43,855 (25.2%) | 22,013 (25.2%) | 112,860 (36.6%) | 56,538 (36.7%) |
| Clinical-epideiological | 62,711 (46.9%) | 31,430 (47.0%) | 128,994 (74.0%) | 64,579 (73.9%) | 191,705 (62.2%) | 96,009 (62.2%) |
| Not concluded | 201 (0.2%) | 99 (0.1%) | 662 (0.4%) | 350 (0.4%) | 863 (0.3%) | 449 (0.3%) |
| Missing | 1,741 (1.3%) | 877 (1.3%) | 846 (0.5%) | 392 (0.4%) | 2,587 (0.8%) | 1,269 (0.8%) |
| **Case outcome** |  |  |  |  |  |  |
| Recovered | 37,270 (27.9%) | 18,576 (27.8%) | 146,188 (83.8%) | 73,034 (83.6%) | 183,458 (59.6%) | 91,610 (59.4%) |
| Death (arbovirus) | 0 (0%) | 0 (0%) | 79 (0.0%) | 37 (0.0%) | 79 (0.0%) | 37 (0.0%) |
| Death (other causes) | 204 (0.2%) | 101 (0.2%) | 29 (0.0%) | 11 (0.0%) | 233 (0.1%) | 112 (0.1%) |
| Death (investigating) | 1 (0.0%) | 0 (0%) | 0 (0%) | 3 (0.0%) | 1 (0.0%) | 3 (0.0%) |
| Ignored | 76,560 (57.3%) | 38,460 (57.5%) | 24,504 (14.1%) | 12,455 (14.3%) | 101,064 (32.8%) | 50,915 (33.0%) |
| Missing | 19,623 (14.7%) | 9,794 (14.6%) | 3,557 (2.0%) | 1,794 (2.1%) | 23,180 (7.5%) | 11,588 (7.5%) |
| **Hospitalization** |  |  |  |  |  |  |
| No | 130,925 (98.0%) | 65,573 (98.0%) | 170,329 (97.7%) | 85,360 (97.7%) | 301,254 (97.8%) | 150,933 (97.8%) |
| Yes | 2,189 (1.6%) | 1,087 (1.6%) | 3,986 (2.3%) | 1,959 (2.2%) | 6,175 (2.0%) | 3,046 (2.0%) |
| Ignored | 25 (0.0%) | 10 (0.0%) | 35 (0.0%) | 14 (0.0%) | 60 (0.0%) | 24 (0.0%) |
| Missing | 519 (0.4%) | 261 (0.4%) | 7 (0.0%) | 1 (0.0%) | 526 (0.2%) | 262 (0.2%) |
| **Postural hypotension and/or syncope** |  |  |  |  |  |  |
| No | 256 (0.2%) | 116 (0.2%) | 2,969 (1.7%) | 1,461 (1.7%) | 3,225 (1.0%) | 1,577 (1.0%) |
| Yes | 93 (0.1%) | 37 (0.1%) | 724 (0.4%) | 339 (0.4%) | 817 (0.3%) | 376 (0.2%) |
| Ignored | 23 (0.0%) | 18 (0.0%) | 206 (0.1%) | 104 (0.1%) | 229 (0.1%) | 122 (0.1%) |
| Missing | 133,286 (99.7%) | 66,760 (99.7%) | 170,458 (97.8%) | 85,430 (97.8%) | 303,744 (98.6%) | 152,190 (98.7%) |
| **Abrupt platelet count drop** |  |  |  |  |  |  |
| No | 189 (0.1%) | 80 (0.1%) | 1,562 (0.9%) | 737 (0.8%) | 1,751 (0.6%) | 817 (0.5%) |
| Yes | 144 (0.1%) | 72 (0.1%) | 2,071 (1.2%) | 1,026 (1.2%) | 2,215 (0.7%) | 1,098 (0.7%) |
| Ignored | 37 (0.0%) | 20 (0.0%) | 263 (0.2%) | 133 (0.2%) | 300 (0.1%) | 153 (0.1%) |
| Missing | 133,288 (99.7%) | 66,759 (99.7%) | 170,461 (97.8%) | 85,438 (97.8%) | 303,749 (98.6%) | 152,197 (98.7%) |
| **Persistent vomiting** |  |  |  |  |  |  |
| No | 276 (0.2%) | 122 (0.2%) | 2,978 (1.7%) | 1,483 (1.7%) | 3,254 (1.1%) | 1,605 (1.0%) |
| Yes | 69 (0.1%) | 35 (0.1%) | 709 (0.4%) | 304 (0.3%) | 778 (0.3%) | 339 (0.2%) |
| Ignored | 24 (0.0%) | 14 (0.0%) | 208 (0.1%) | 110 (0.1%) | 232 (0.1%) | 124 (0.1%) |
| Missing | 133,289 (99.7%) | 66,760 (99.7%) | 170,462 (97.8%) | 85,437 (97.8%) | 303,751 (98.6%) | 152,197 (98.7%) |
| **Intense and continuous abdominal pain** |  |  |  |  |  |  |
| No | 217 (0.2%) | 95 (0.1%) | 2,398 (1.4%) | 1,201 (1.4%) | 2,615 (0.8%) | 1,296 (0.8%) |
| Yes | 130 (0.1%) | 59 (0.1%) | 1,334 (0.8%) | 608 (0.7%) | 1,464 (0.5%) | 667 (0.4%) |
| Ignored | 24 (0.0%) | 17 (0.0%) | 169 (0.1%) | 87 (0.1%) | 193 (0.1%) | 104 (0.1%) |
| Missing | 133,287 (99.7%) | 66,760 (99.7%) | 170,456 (97.8%) | 85,438 (97.8%) | 303,743 (98.6%) | 152,198 (98.7%) |
| **Lethargy or irritability** |  |  |  |  |  |  |
| No | 277 (0.2%) | 117 (0.2%) | 2,890 (1.7%) | 1,385 (1.6%) | 3,167 (1.0%) | 1,502 (1.0%) |
| Yes | 60 (0.0%) | 35 (0.1%) | 759 (0.4%) | 391 (0.4%) | 819 (0.3%) | 426 (0.3%) |
| Ignored | 33 (0.0%) | 19 (0.0%) | 243 (0.1%) | 118 (0.1%) | 276 (0.1%) | 137 (0.1%) |
| Missing | 133,288 (99.7%) | 66,760 (99.7%) | 170,465 (97.8%) | 85,440 (97.8%) | 303,753 (98.6%) | 152,200 (98.7%) |
| **Mucosal bleeding/other hemorrhages** |  |  |  |  |  |  |
| No | 253 (0.2%) | 116 (0.2%) | 2,965 (1.7%) | 1,391 (1.6%) | 3,218 (1.0%) | 1,507 (1.0%) |
| Yes | 92 (0.1%) | 39 (0.1%) | 713 (0.4%) | 406 (0.5%) | 805 (0.3%) | 445 (0.3%) |
| Ignored | 24 (0.0%) | 16 (0.0%) | 215 (0.1%) | 100 (0.1%) | 239 (0.1%) | 116 (0.1%) |
| Missing | 133,289 (99.7%) | 66,760 (99.7%) | 170,464 (97.8%) | 85,437 (97.8%) | 303,753 (98.6%) | 152,197 (98.7%) |
| **Progressive hematocrit increase** |  |  |  |  |  |  |
| No | 309 (0.2%) | 132 (0.2%) | 3,226 (1.9%) | 1,544 (1.8%) | 3,535 (1.1%) | 1,676 (1.1%) |
| Yes | 14 (0.0%) | 4 (0.0%) | 240 (0.1%) | 116 (0.1%) | 254 (0.1%) | 120 (0.1%) |
| Ignored | 47 (0.0%) | 35 (0.1%) | 431 (0.2%) | 235 (0.3%) | 478 (0.2%) | 270 (0.2%) |
| Missing | 133,288 (99.7%) | 66,760 (99.7%) | 170,460 (97.8%) | 85,439 (97.8%) | 303,748 (98.6%) | 152,199 (98.7%) |
| **Hepatomegaly >= 2cm** |  |  |  |  |  |  |
| No | 307 (0.2%) | 139 (0.2%) | 3,308 (1.9%) | 1,603 (1.8%) | 3,615 (1.2%) | 1,742 (1.1%) |
| Yes | 13 (0.0%) | 5 (0.0%) | 79 (0.0%) | 36 (0.0%) | 92 (0.0%) | 41 (0.0%) |
| Ignored | 49 (0.0%) | 27 (0.0%) | 505 (0.3%) | 255 (0.3%) | 554 (0.2%) | 282 (0.2%) |
| Missing | 133,289 (99.7%) | 66,760 (99.7%) | 170,465 (97.8%) | 85,440 (97.8%) | 303,754 (98.6%) | 152,200 (98.7%) |
| **Fluid accumulation** |  |  |  |  |  |  |
| No | 304 (0.2%) | 134 (0.2%) | 3,365 (1.9%) | 1,645 (1.9%) | 3,669 (1.2%) | 1,779 (1.2%) |
| Yes | 26 (0.0%) | 17 (0.0%) | 142 (0.1%) | 62 (0.1%) | 168 (0.1%) | 79 (0.1%) |
| Ignored | 38 (0.0%) | 20 (0.0%) | 386 (0.2%) | 187 (0.2%) | 424 (0.1%) | 207 (0.1%) |
| Missing | 133,290 (99.7%) | 66,760 (99.7%) | 170,464 (97.8%) | 85,440 (97.8%) | 303,754 (98.6%) | 152,200 (98.7%) |
| **Weak or undetectable pulse** |  |  |  |  |  |  |
| No | 33 (0.0%) | 7 (0.0%) | 165 (0.1%) | 90 (0.1%) | 198 (0.1%) | 97 (0.1%) |
| Yes | 15 (0.0%) | 6 (0.0%) | 42 (0.0%) | 19 (0.0%) | 57 (0.0%) | 25 (0.0%) |
| Ignored | 13 (0.0%) | 8 (0.0%) | 29 (0.0%) | 15 (0.0%) | 42 (0.0%) | 23 (0.0%) |
| Missing | 133,597 (100.0%) | 66,910 (100.0%) | 174,121 (99.9%) | 87,210 (99.9%) | 307,718 (99.9%) | 154,120 (99.9%) |
| **Convergent pulse pressure <= 20 mmHg** |  |  |  |  |  |  |
| No | 39 (0.0%) | 8 (0.0%) | 180 (0.1%) | 104 (0.1%) | 219 (0.1%) | 112 (0.1%) |
| Yes | 4 (0.0%) | 3 (0.0%) | 17 (0.0%) | 5 (0.0%) | 21 (0.0%) | 8 (0.0%) |
| Ignored | 18 (0.0%) | 10 (0.0%) | 36 (0.0%) | 15 (0.0%) | 54 (0.0%) | 25 (0.0%) |
| Missing | 133,597 (100.0%) | 66,910 (100.0%) | 174,124 (99.9%) | 87,210 (99.9%) | 307,721 (99.9%) | 154,120 (99.9%) |
| **Capillary refill time** |  |  |  |  |  |  |
| No | 27 (0.0%) | 7 (0.0%) | 143 (0.1%) | 66 (0.1%) | 170 (0.1%) | 73 (0.0%) |
| Yes | 12 (0.0%) | 4 (0.0%) | 45 (0.0%) | 35 (0.0%) | 57 (0.0%) | 39 (0.0%) |
| Ignored | 22 (0.0%) | 10 (0.0%) | 43 (0.0%) | 22 (0.0%) | 65 (0.0%) | 32 (0.0%) |
| Missing | 133,597 (100.0%) | 66,910 (100.0%) | 174,126 (99.9%) | 87,211 (99.9%) | 307,723 (99.9%) | 154,121 (99.9%) |
| **Fluid accumulation with respiratory insufficiency** |  |  |  |  |  |  |
| No | 32 (0.0%) | 5 (0.0%) | 153 (0.1%) | 88 (0.1%) | 185 (0.1%) | 93 (0.1%) |
| Yes | 13 (0.0%) | 7 (0.0%) | 47 (0.0%) | 22 (0.0%) | 60 (0.0%) | 29 (0.0%) |
| Ignored | 16 (0.0%) | 9 (0.0%) | 33 (0.0%) | 13 (0.0%) | 49 (0.0%) | 22 (0.0%) |
| Missing | 133,597 (100.0%) | 66,910 (100.0%) | 174,124 (99.9%) | 87,211 (99.9%) | 307,721 (99.9%) | 154,121 (99.9%) |
| **Tachycardia** |  |  |  |  |  |  |
| No | 22 (0.0%) | 6 (0.0%) | 113 (0.1%) | 72 (0.1%) | 135 (0.0%) | 78 (0.1%) |
| Yes | 24 (0.0%) | 6 (0.0%) | 82 (0.0%) | 36 (0.0%) | 106 (0.0%) | 42 (0.0%) |
| Ignored | 15 (0.0%) | 9 (0.0%) | 35 (0.0%) | 14 (0.0%) | 50 (0.0%) | 23 (0.0%) |
| Missing | 133,597 (100.0%) | 66,910 (100.0%) | 174,127 (99.9%) | 87,212 (99.9%) | 307,724 (99.9%) | 154,122 (99.9%) |
| **Cold extremities** |  |  |  |  |  |  |
| No | 28 (0.0%) | 8 (0.0%) | 151 (0.1%) | 76 (0.1%) | 179 (0.1%) | 84 (0.1%) |
| Yes | 19 (0.0%) | 5 (0.0%) | 54 (0.0%) | 31 (0.0%) | 73 (0.0%) | 36 (0.0%) |
| Ignored | 14 (0.0%) | 8 (0.0%) | 28 (0.0%) | 16 (0.0%) | 42 (0.0%) | 24 (0.0%) |
| Missing | 133,597 (100.0%) | 66,910 (100.0%) | 174,124 (99.9%) | 87,211 (99.9%) | 307,721 (99.9%) | 154,121 (99.9%) |
| **Late stage arterial hypotension** |  |  |  |  |  |  |
| No | 33 (0.0%) | 9 (0.0%) | 149 (0.1%) | 83 (0.1%) | 182 (0.1%) | 92 (0.1%) |
| Yes | 13 (0.0%) | 2 (0.0%) | 52 (0.0%) | 29 (0.0%) | 65 (0.0%) | 31 (0.0%) |
| Ignored | 15 (0.0%) | 10 (0.0%) | 31 (0.0%) | 11 (0.0%) | 46 (0.0%) | 21 (0.0%) |
| Missing | 133,597 (100.0%) | 66,910 (100.0%) | 174,125 (99.9%) | 87,211 (99.9%) | 307,722 (99.9%) | 154,121 (99.9%) |
| **Hematemesis** |  |  |  |  |  |  |
| No | 47 (0.0%) | 10 (0.0%) | 187 (0.1%) | 101 (0.1%) | 234 (0.1%) | 111 (0.1%) |
| Yes | 3 (0.0%) | 3 (0.0%) | 16 (0.0%) | 11 (0.0%) | 19 (0.0%) | 14 (0.0%) |
| Ignored | 11 (0.0%) | 8 (0.0%) | 28 (0.0%) | 11 (0.0%) | 39 (0.0%) | 19 (0.0%) |
| Missing | 133,597 (100.0%) | 66,910 (100.0%) | 174,126 (99.9%) | 87,211 (99.9%) | 307,723 (99.9%) | 154,121 (99.9%) |
| **Melena** |  |  |  |  |  |  |
| No | 45 (0.0%) | 10 (0.0%) | 179 (0.1%) | 104 (0.1%) | 224 (0.1%) | 114 (0.1%) |
| Yes | 5 (0.0%) | 2 (0.0%) | 22 (0.0%) | 8 (0.0%) | 27 (0.0%) | 10 (0.0%) |
| Ignored | 11 (0.0%) | 9 (0.0%) | 29 (0.0%) | 11 (0.0%) | 40 (0.0%) | 20 (0.0%) |
| Missing | 133,597 (100.0%) | 66,910 (100.0%) | 174,127 (99.9%) | 87,211 (99.9%) | 307,724 (99.9%) | 154,121 (99.9%) |
| **Voluminous metrorrhagia** |  |  |  |  |  |  |
| No | 49 (0.0%) | 13 (0.0%) | 200 (0.1%) | 109 (0.1%) | 249 (0.1%) | 122 (0.1%) |
| Yes | 3 (0.0%) | 2 (0.0%) | 7 (0.0%) | 3 (0.0%) | 10 (0.0%) | 5 (0.0%) |
| Ignored | 9 (0.0%) | 6 (0.0%) | 26 (0.0%) | 11 (0.0%) | 35 (0.0%) | 17 (0.0%) |
| Missing | 133,597 (100.0%) | 66,910 (100.0%) | 174,124 (99.9%) | 87,211 (99.9%) | 307,721 (99.9%) | 154,121 (99.9%) |
| **CNS bleeding** |  |  |  |  |  |  |
| No | 45 (0.0%) | 11 (0.0%) | 181 (0.1%) | 101 (0.1%) | 226 (0.1%) | 112 (0.1%) |
| Yes | 2 (0.0%) | 1 (0.0%) | 3 (0.0%) | 5 (0.0%) | 5 (0.0%) | 6 (0.0%) |
| Ignored | 14 (0.0%) | 9 (0.0%) | 46 (0.0%) | 17 (0.0%) | 60 (0.0%) | 26 (0.0%) |
| Missing | 133,597 (100.0%) | 66,910 (100.0%) | 174,127 (99.9%) | 87,211 (99.9%) | 307,724 (99.9%) | 154,121 (99.9%) |
| **AST/ALT > 1,000** |  |  |  |  |  |  |
| No | 43 (0.0%) | 9 (0.0%) | 177 (0.1%) | 98 (0.1%) | 220 (0.1%) | 107 (0.1%) |
| Yes | 5 (0.0%) | 2 (0.0%) | 11 (0.0%) | 4 (0.0%) | 16 (0.0%) | 6 (0.0%) |
| Ignored | 13 (0.0%) | 10 (0.0%) | 42 (0.0%) | 20 (0.0%) | 55 (0.0%) | 30 (0.0%) |
| Missing | 133,597 (100.0%) | 66,910 (100.0%) | 174,127 (99.9%) | 87,212 (99.9%) | 307,724 (99.9%) | 154,122 (99.9%) |
| **Myocarditis** |  |  |  |  |  |  |
| No | 38 (0.0%) | 11 (0.0%) | 186 (0.1%) | 102 (0.1%) | 224 (0.1%) | 113 (0.1%) |
| Yes | 3 (0.0%) | 1 (0.0%) | 3 (0.0%) | 1 (0.0%) | 6 (0.0%) | 2 (0.0%) |
| Ignored | 20 (0.0%) | 9 (0.0%) | 43 (0.0%) | 20 (0.0%) | 63 (0.0%) | 29 (0.0%) |
| Missing | 133,597 (100.0%) | 66,910 (100.0%) | 174,125 (99.9%) | 87,211 (99.9%) | 307,722 (99.9%) | 154,121 (99.9%) |
| **Altered consciousness** |  |  |  |  |  |  |
| No | 25 (0.0%) | 2 (0.0%) | 147 (0.1%) | 75 (0.1%) | 172 (0.1%) | 77 (0.1%) |
| Yes | 24 (0.0%) | 16 (0.0%) | 52 (0.0%) | 37 (0.0%) | 76 (0.0%) | 53 (0.0%) |
| Ignored | 12 (0.0%) | 3 (0.0%) | 31 (0.0%) | 11 (0.0%) | 43 (0.0%) | 14 (0.0%) |
| Missing | 133,597 (100.0%) | 66,910 (100.0%) | 174,127 (99.9%) | 87,211 (99.9%) | 307,724 (99.9%) | 154,121 (99.9%) |
| **Other severe organs involvement** |  |  |  |  |  |  |
| No | 32 (0.0%) | 5 (0.0%) | 142 (0.1%) | 88 (0.1%) | 174 (0.1%) | 93 (0.1%) |
| Yes | 17 (0.0%) | 11 (0.0%) | 49 (0.0%) | 21 (0.0%) | 66 (0.0%) | 32 (0.0%) |
| Ignored | 12 (0.0%) | 5 (0.0%) | 39 (0.0%) | 13 (0.0%) | 51 (0.0%) | 18 (0.0%) |
| Missing | 133,597 (100.0%) | 66,910 (100.0%) | 174,127 (99.9%) | 87,212 (99.9%) | 307,724 (99.9%) | 154,122 (99.9%) |
| IQR = interquartile range; SD = Standard deviation; CNS = central nervous system; AST = aspartate aminotransferase; ALT = alanine aminotransferase; RT-PCR = reverse transcription polymerase chain reaction. | | | | | | |

**Table S 5:** Final dataset variables (including epidemiological, clinical at presentations and disease progressions) counts and frequencies by Ccikungunya diagnostic investigation outcome in each dataset split.

|  | **No** | | **Yes** | | **Overall** | |
| --- | --- | --- | --- | --- | --- | --- |
|  | **Train (N=295548)** | **Test (N=148056)** | **Train (N=12467)** | **Test (N=6209)** | **Train (N=308015)** | **Test (N=154265)** |
| **Age at notification** |  |  |  |  |  |  |
| Mean (SD) | 35.0 (19.0) | 34.8 (19.0) | 43.0 (19.0) | 43.2 (18.8) | 35.2 (19.1) | 35.1 (19.0) |
| Median [Min, Max] | 33.0 [0, 106] | 33.0 [0, 104] | 43.0 [0, 98.0] | 43.0 [0, 95.0] | 33.0 [0, 106] | 33.0 [0, 104] |
| Missing | 2,950 (1.0%) | 1,515 (1.0%) | 4,358 (35.0%) | 2,211 (35.6%) | 7,308 (2.4%) | 3,726 (2.4%) |
| **Age at notification** |  |  |  |  |  |  |
| [0,1] | 3,479 (1.2%) | 1,821 (1.2%) | 14 (0.1%) | 6 (0.1%) | 3,493 (1.1%) | 1,827 (1.2%) |
| (1,12] | 29,453 (10.0%) | 14,841 (10.0%) | 438 (3.5%) | 225 (3.6%) | 29,891 (9.7%) | 15,066 (9.8%) |
| (12,18] | 27,882 (9.4%) | 14,071 (9.5%) | 476 (3.8%) | 230 (3.7%) | 28,358 (9.2%) | 14,301 (9.3%) |
| (18,25] | 44,226 (15.0%) | 22,357 (15.1%) | 757 (6.1%) | 348 (5.6%) | 44,983 (14.6%) | 22,705 (14.7%) |
| (25,35] | 54,204 (18.3%) | 27,287 (18.4%) | 1,235 (9.9%) | 594 (9.6%) | 55,439 (18.0%) | 27,881 (18.1%) |
| (35,45] | 49,727 (16.8%) | 24,842 (16.8%) | 1,475 (11.8%) | 767 (12.4%) | 51,202 (16.6%) | 25,609 (16.6%) |
| (45,55] | 36,438 (12.3%) | 18,052 (12.2%) | 1,426 (11.4%) | 690 (11.1%) | 37,864 (12.3%) | 18,742 (12.1%) |
| (55,65] | 25,815 (8.7%) | 12,766 (8.6%) | 1,251 (10.0%) | 629 (10.1%) | 27,066 (8.8%) | 13,395 (8.7%) |
| (65,75] | 14,643 (5.0%) | 7,156 (4.8%) | 714 (5.7%) | 361 (5.8%) | 15,357 (5.0%) | 7,517 (4.9%) |
| (75,85] | 5,370 (1.8%) | 2,639 (1.8%) | 280 (2.2%) | 129 (2.1%) | 5,650 (1.8%) | 2,768 (1.8%) |
| (85,95] | 1,302 (0.4%) | 664 (0.4%) | 42 (0.3%) | 19 (0.3%) | 1,344 (0.4%) | 683 (0.4%) |
| Missing | 3,009 (1.0%) | 1,560 (1.1%) | 4,359 (35.0%) | 2,211 (35.6%) | 7,368 (2.4%) | 3,771 (2.4%) |
| **Sex at notification** |  |  |  |  |  |  |
| Female | 157,698 (53.4%) | 79,150 (53.5%) | 5,074 (40.7%) | 2,461 (39.6%) | 162,772 (52.8%) | 81,611 (52.9%) |
| Male | 134,888 (45.6%) | 67,386 (45.5%) | 3,035 (24.3%) | 1,537 (24.8%) | 137,923 (44.8%) | 68,923 (44.7%) |
| Ignored | 12 (0.0%) | 5 (0.0%) | 0 (0%) | 0 (0%) | 12 (0.0%) | 5 (0.0%) |
| Missing | 2,950 (1.0%) | 1,515 (1.0%) | 4,358 (35.0%) | 2,211 (35.6%) | 7,308 (2.4%) | 3,726 (2.4%) |
| **Pregnancy** |  |  |  |  |  |  |
| No | 261,872 (88.6%) | 130,904 (88.4%) | 7,373 (59.1%) | 3,630 (58.5%) | 269,245 (87.4%) | 134,534 (87.2%) |
| Yes | 3,195 (1.1%) | 1,683 (1.1%) | 156 (1.3%) | 72 (1.2%) | 3,351 (1.1%) | 1,755 (1.1%) |
| Ignored/Unknown | 27,531 (9.3%) | 13,954 (9.4%) | 580 (4.7%) | 296 (4.8%) | 28,111 (9.1%) | 14,250 (9.2%) |
| Missing | 2,950 (1.0%) | 1,515 (1.0%) | 4,358 (35.0%) | 2,211 (35.6%) | 7,308 (2.4%) | 3,726 (2.4%) |
| **Race or skin color** |  |  |  |  |  |  |
| White | 90,555 (30.6%) | 45,366 (30.6%) | 2,075 (16.6%) | 1,029 (16.6%) | 92,630 (30.1%) | 46,395 (30.1%) |
| Black | 155,743 (52.7%) | 78,019 (52.7%) | 4,889 (39.2%) | 2,428 (39.1%) | 160,632 (52.2%) | 80,447 (52.1%) |
| Yellow | 26,744 (9.0%) | 13,416 (9.1%) | 740 (5.9%) | 341 (5.5%) | 27,484 (8.9%) | 13,757 (8.9%) |
| Indigenous | 276 (0.1%) | 143 (0.1%) | 5 (0.0%) | 5 (0.1%) | 281 (0.1%) | 148 (0.1%) |
| Ignored/Unknown | 19,280 (6.5%) | 9,597 (6.5%) | 400 (3.2%) | 195 (3.1%) | 19,680 (6.4%) | 9,792 (6.3%) |
| Missing | 2,950 (1.0%) | 1,515 (1.0%) | 4,358 (35.0%) | 2,211 (35.6%) | 7,308 (2.4%) | 3,726 (2.4%) |
| **Indigenous ethnicity** |  |  |  |  |  |  |
| Tupiniquim | 113 (0.0%) | 49 (0.0%) | 0 (0%) | 0 (0%) | 113 (0.0%) | 49 (0.0%) |
| Guarani | 25 (0.0%) | 18 (0.0%) | 0 (0%) | 0 (0%) | 25 (0.0%) | 18 (0.0%) |
| Ignored/Unknown | 138 (0.0%) | 76 (0.1%) | 5 (0.0%) | 5 (0.1%) | 143 (0.0%) | 81 (0.1%) |
| Not applicable | 292,322 (98.9%) | 146,398 (98.9%) | 8,104 (65.0%) | 3,993 (64.3%) | 300,426 (97.5%) | 150,391 (97.5%) |
| Missing | 2,950 (1.0%) | 1,515 (1.0%) | 4,358 (35.0%) | 2,211 (35.6%) | 7,308 (2.4%) | 3,726 (2.4%) |
| **Person with special needs** |  |  |  |  |  |  |
| Yes | 276 (0.1%) | 143 (0.1%) | 31 (0.2%) | 15 (0.2%) | 307 (0.1%) | 158 (0.1%) |
| No | 292,322 (98.9%) | 146,398 (98.9%) | 8,078 (64.8%) | 3,983 (64.1%) | 300,400 (97.5%) | 150,381 (97.5%) |
| Missing | 2,950 (1.0%) | 1,515 (1.0%) | 4,358 (35.0%) | 2,211 (35.6%) | 7,308 (2.4%) | 3,726 (2.4%) |
| **Homeless** |  |  |  |  |  |  |
| Yes | 97 (0.0%) | 43 (0.0%) | 5 (0.0%) | 3 (0.0%) | 102 (0.0%) | 46 (0.0%) |
| No | 292,501 (99.0%) | 146,498 (98.9%) | 8,104 (65.0%) | 3,995 (64.3%) | 300,605 (97.6%) | 150,493 (97.6%) |
| Missing | 2,950 (1.0%) | 1,515 (1.0%) | 4,358 (35.0%) | 2,211 (35.6%) | 7,308 (2.4%) | 3,726 (2.4%) |
| **Education** |  |  |  |  |  |  |
| Illiterate | 13,664 (4.6%) | 6,697 (4.5%) | 402 (3.2%) | 199 (3.2%) | 14,066 (4.6%) | 6,896 (4.5%) |
| Elementary School Complete | 30,221 (10.2%) | 15,076 (10.2%) | 920 (7.4%) | 468 (7.5%) | 31,141 (10.1%) | 15,544 (10.1%) |
| Middle School Complete | 27,712 (9.4%) | 13,808 (9.3%) | 692 (5.6%) | 344 (5.5%) | 28,404 (9.2%) | 14,152 (9.2%) |
| High School Complete | 51,018 (17.3%) | 25,589 (17.3%) | 1,705 (13.7%) | 840 (13.5%) | 52,723 (17.1%) | 26,429 (17.1%) |
| Higher Education Complete | 14,067 (4.8%) | 7,020 (4.7%) | 554 (4.4%) | 249 (4.0%) | 14,621 (4.7%) | 7,269 (4.7%) |
| Ignored/Unknown | 155,916 (52.8%) | 78,351 (52.9%) | 3,836 (30.8%) | 1,898 (30.6%) | 159,752 (51.9%) | 80,249 (52.0%) |
| Missing | 2,950 (1.0%) | 1,515 (1.0%) | 4,358 (35.0%) | 2,211 (35.6%) | 7,308 (2.4%) | 3,726 (2.4%) |
| **Home address** |  |  |  |  |  |  |
| Urban | 227,418 (76.9%) | 113,690 (76.8%) | 7,328 (58.8%) | 3,609 (58.1%) | 234,746 (76.2%) | 117,299 (76.0%) |
| Rural | 45,219 (15.3%) | 22,764 (15.4%) | 384 (3.1%) | 181 (2.9%) | 45,603 (14.8%) | 22,945 (14.9%) |
| Peri-urban | 929 (0.3%) | 487 (0.3%) | 8 (0.1%) | 5 (0.1%) | 937 (0.3%) | 492 (0.3%) |
| Unknown | 19,029 (6.4%) | 9,597 (6.5%) | 389 (3.1%) | 203 (3.3%) | 19,418 (6.3%) | 9,800 (6.4%) |
| Missing | 2,953 (1.0%) | 1,518 (1.0%) | 4,358 (35.0%) | 2,211 (35.6%) | 7,311 (2.4%) | 3,729 (2.4%) |
| **Autochthonous case** |  |  |  |  |  |  |
| No | 497 (0.2%) | 211 (0.1%) | 17 (0.1%) | 7 (0.1%) | 514 (0.2%) | 218 (0.1%) |
| Yes | 293,991 (99.5%) | 147,312 (99.5%) | 12,436 (99.8%) | 6,190 (99.7%) | 306,427 (99.5%) | 153,502 (99.5%) |
| Unknown | 324 (0.1%) | 173 (0.1%) | 12 (0.1%) | 11 (0.2%) | 336 (0.1%) | 184 (0.1%) |
| Missing | 736 (0.2%) | 360 (0.2%) | 2 (0.0%) | 1 (0.0%) | 738 (0.2%) | 361 (0.2%) |
| **Fever** |  |  |  |  |  |  |
| No | 67,526 (22.8%) | 33,820 (22.8%) | 3,866 (31.0%) | 1,948 (31.4%) | 71,392 (23.2%) | 35,768 (23.2%) |
| Yes | 227,512 (77.0%) | 113,979 (77.0%) | 8,601 (69.0%) | 4,261 (68.6%) | 236,113 (76.7%) | 118,240 (76.6%) |
| Missing | 510 (0.2%) | 257 (0.2%) | 0 (0%) | 0 (0%) | 510 (0.2%) | 257 (0.2%) |
| **Headache** |  |  |  |  |  |  |
| No | 69,009 (23.3%) | 34,571 (23.4%) | 4,828 (38.7%) | 2,331 (37.5%) | 73,837 (24.0%) | 36,902 (23.9%) |
| Yes | 226,029 (76.5%) | 113,228 (76.5%) | 7,639 (61.3%) | 3,878 (62.5%) | 233,668 (75.9%) | 117,106 (75.9%) |
| Missing | 510 (0.2%) | 257 (0.2%) | 0 (0%) | 0 (0%) | 510 (0.2%) | 257 (0.2%) |
| **Vomiting** |  |  |  |  |  |  |
| No | 232,946 (78.8%) | 116,833 (78.9%) | 10,793 (86.6%) | 5,370 (86.5%) | 243,739 (79.1%) | 122,203 (79.2%) |
| Yes | 62,092 (21.0%) | 30,966 (20.9%) | 1,674 (13.4%) | 839 (13.5%) | 63,766 (20.7%) | 31,805 (20.6%) |
| Missing | 510 (0.2%) | 257 (0.2%) | 0 (0%) | 0 (0%) | 510 (0.2%) | 257 (0.2%) |
| **Back pain** |  |  |  |  |  |  |
| No | 233,717 (79.1%) | 117,061 (79.1%) | 8,804 (70.6%) | 4,425 (71.3%) | 242,521 (78.7%) | 121,486 (78.8%) |
| Yes | 61,321 (20.7%) | 30,738 (20.8%) | 3,663 (29.4%) | 1,784 (28.7%) | 64,984 (21.1%) | 32,522 (21.1%) |
| Missing | 510 (0.2%) | 257 (0.2%) | 0 (0%) | 0 (0%) | 510 (0.2%) | 257 (0.2%) |
| **Arthritis** |  |  |  |  |  |  |
| No | 276,582 (93.6%) | 138,663 (93.7%) | 9,950 (79.8%) | 4,989 (80.4%) | 286,532 (93.0%) | 143,652 (93.1%) |
| Yes | 18,456 (6.2%) | 9,136 (6.2%) | 2,517 (20.2%) | 1,220 (19.6%) | 20,973 (6.8%) | 10,356 (6.7%) |
| Missing | 510 (0.2%) | 257 (0.2%) | 0 (0%) | 0 (0%) | 510 (0.2%) | 257 (0.2%) |
| **Petechiae** |  |  |  |  |  |  |
| No | 281,870 (95.4%) | 141,411 (95.5%) | 11,401 (91.4%) | 5,668 (91.3%) | 293,271 (95.2%) | 147,079 (95.3%) |
| Yes | 13,168 (4.5%) | 6,388 (4.3%) | 1,066 (8.6%) | 541 (8.7%) | 14,234 (4.6%) | 6,929 (4.5%) |
| Missing | 510 (0.2%) | 257 (0.2%) | 0 (0%) | 0 (0%) | 510 (0.2%) | 257 (0.2%) |
| **Tourniquet test** |  |  |  |  |  |  |
| No | 291,309 (98.6%) | 146,069 (98.7%) | 12,274 (98.5%) | 6,102 (98.3%) | 303,583 (98.6%) | 152,171 (98.6%) |
| Yes | 3,729 (1.3%) | 1,730 (1.2%) | 193 (1.5%) | 107 (1.7%) | 3,922 (1.3%) | 1,837 (1.2%) |
| Missing | 510 (0.2%) | 257 (0.2%) | 0 (0%) | 0 (0%) | 510 (0.2%) | 257 (0.2%) |
| **Myalgia** |  |  |  |  |  |  |
| No | 80,788 (27.3%) | 40,567 (27.4%) | 3,999 (32.1%) | 2,012 (32.4%) | 84,787 (27.5%) | 42,579 (27.6%) |
| Yes | 214,250 (72.5%) | 107,232 (72.4%) | 8,468 (67.9%) | 4,197 (67.6%) | 222,718 (72.3%) | 111,429 (72.2%) |
| Missing | 510 (0.2%) | 257 (0.2%) | 0 (0%) | 0 (0%) | 510 (0.2%) | 257 (0.2%) |
| **Rash** |  |  |  |  |  |  |
| No | 282,918 (95.7%) | 141,674 (95.7%) | 11,158 (89.5%) | 5,570 (89.7%) | 294,076 (95.5%) | 147,244 (95.4%) |
| Yes | 12,120 (4.1%) | 6,125 (4.1%) | 1,309 (10.5%) | 639 (10.3%) | 13,429 (4.4%) | 6,764 (4.4%) |
| Missing | 510 (0.2%) | 257 (0.2%) | 0 (0%) | 0 (0%) | 510 (0.2%) | 257 (0.2%) |
| **Nausea** |  |  |  |  |  |  |
| No | 191,592 (64.8%) | 96,115 (64.9%) | 8,706 (69.8%) | 4,313 (69.5%) | 200,298 (65.0%) | 100,428 (65.1%) |
| Yes | 103,446 (35.0%) | 51,684 (34.9%) | 3,761 (30.2%) | 1,896 (30.5%) | 107,207 (34.8%) | 53,580 (34.7%) |
| Missing | 510 (0.2%) | 257 (0.2%) | 0 (0%) | 0 (0%) | 510 (0.2%) | 257 (0.2%) |
| **Conjunctivitis** |  |  |  |  |  |  |
| No | 292,216 (98.9%) | 146,426 (98.9%) | 12,182 (97.7%) | 6,078 (97.9%) | 304,398 (98.8%) | 152,504 (98.9%) |
| Yes | 2,822 (1.0%) | 1,373 (0.9%) | 285 (2.3%) | 131 (2.1%) | 3,107 (1.0%) | 1,504 (1.0%) |
| Missing | 510 (0.2%) | 257 (0.2%) | 0 (0%) | 0 (0%) | 510 (0.2%) | 257 (0.2%) |
| **Intense arthralgia** |  |  |  |  |  |  |
| No | 239,410 (81.0%) | 119,935 (81.0%) | 5,512 (44.2%) | 2,799 (45.1%) | 244,922 (79.5%) | 122,734 (79.6%) |
| Yes | 55,628 (18.8%) | 27,864 (18.8%) | 6,955 (55.8%) | 3,410 (54.9%) | 62,583 (20.3%) | 31,274 (20.3%) |
| Missing | 510 (0.2%) | 257 (0.2%) | 0 (0%) | 0 (0%) | 510 (0.2%) | 257 (0.2%) |
| **Leukopenia** |  |  |  |  |  |  |
| No | 289,668 (98.0%) | 145,098 (98.0%) | 12,252 (98.3%) | 6,096 (98.2%) | 301,920 (98.0%) | 151,194 (98.0%) |
| Yes | 5,370 (1.8%) | 2,701 (1.8%) | 215 (1.7%) | 113 (1.8%) | 5,585 (1.8%) | 2,814 (1.8%) |
| Missing | 510 (0.2%) | 257 (0.2%) | 0 (0%) | 0 (0%) | 510 (0.2%) | 257 (0.2%) |
| **Retro-orbital pain** |  |  |  |  |  |  |
| No | 205,369 (69.5%) | 102,698 (69.4%) | 8,866 (71.1%) | 4,462 (71.9%) | 214,235 (69.6%) | 107,160 (69.5%) |
| Yes | 89,669 (30.3%) | 45,101 (30.5%) | 3,601 (28.9%) | 1,747 (28.1%) | 93,270 (30.3%) | 46,848 (30.4%) |
| Missing | 510 (0.2%) | 257 (0.2%) | 0 (0%) | 0 (0%) | 510 (0.2%) | 257 (0.2%) |
| **Diabetes** |  |  |  |  |  |  |
| No | 286,766 (97.0%) | 143,719 (97.1%) | 11,567 (92.8%) | 5,741 (92.5%) | 298,333 (96.9%) | 149,460 (96.9%) |
| Yes | 7,929 (2.7%) | 3,933 (2.7%) | 840 (6.7%) | 443 (7.1%) | 8,769 (2.8%) | 4,376 (2.8%) |
| Ignored | 343 (0.1%) | 147 (0.1%) | 60 (0.5%) | 25 (0.4%) | 403 (0.1%) | 172 (0.1%) |
| Missing | 510 (0.2%) | 257 (0.2%) | 0 (0%) | 0 (0%) | 510 (0.2%) | 257 (0.2%) |
| **Hepatopathies** |  |  |  |  |  |  |
| No | 294,377 (99.6%) | 147,485 (99.6%) | 12,378 (99.3%) | 6,161 (99.2%) | 306,755 (99.6%) | 153,646 (99.6%) |
| Yes | 325 (0.1%) | 165 (0.1%) | 25 (0.2%) | 20 (0.3%) | 350 (0.1%) | 185 (0.1%) |
| Ignored | 336 (0.1%) | 149 (0.1%) | 64 (0.5%) | 28 (0.5%) | 400 (0.1%) | 177 (0.1%) |
| Missing | 510 (0.2%) | 257 (0.2%) | 0 (0%) | 0 (0%) | 510 (0.2%) | 257 (0.2%) |
| **Arterial hypertension** |  |  |  |  |  |  |
| No | 273,769 (92.6%) | 137,326 (92.8%) | 10,456 (83.9%) | 5,222 (84.1%) | 284,225 (92.3%) | 142,548 (92.4%) |
| Yes | 20,930 (7.1%) | 10,325 (7.0%) | 1,952 (15.7%) | 963 (15.5%) | 22,882 (7.4%) | 11,288 (7.3%) |
| Ignored | 339 (0.1%) | 148 (0.1%) | 59 (0.5%) | 24 (0.4%) | 398 (0.1%) | 172 (0.1%) |
| Missing | 510 (0.2%) | 257 (0.2%) | 0 (0%) | 0 (0%) | 510 (0.2%) | 257 (0.2%) |
| **Autoimmune diseases** |  |  |  |  |  |  |
| No | 293,835 (99.4%) | 147,228 (99.4%) | 12,303 (98.7%) | 6,126 (98.7%) | 306,138 (99.4%) | 153,354 (99.4%) |
| Yes | 862 (0.3%) | 421 (0.3%) | 98 (0.8%) | 52 (0.8%) | 960 (0.3%) | 473 (0.3%) |
| Ignored | 341 (0.1%) | 150 (0.1%) | 66 (0.5%) | 31 (0.5%) | 407 (0.1%) | 181 (0.1%) |
| Missing | 510 (0.2%) | 257 (0.2%) | 0 (0%) | 0 (0%) | 510 (0.2%) | 257 (0.2%) |
| **Hematological diseases** |  |  |  |  |  |  |
| No | 294,451 (99.6%) | 147,504 (99.6%) | 12,376 (99.3%) | 6,172 (99.4%) | 306,827 (99.6%) | 153,676 (99.6%) |
| Yes | 259 (0.1%) | 151 (0.1%) | 27 (0.2%) | 9 (0.1%) | 286 (0.1%) | 160 (0.1%) |
| Ignored | 328 (0.1%) | 144 (0.1%) | 64 (0.5%) | 28 (0.5%) | 392 (0.1%) | 172 (0.1%) |
| Missing | 510 (0.2%) | 257 (0.2%) | 0 (0%) | 0 (0%) | 510 (0.2%) | 257 (0.2%) |
| **Chronic kidney disease** |  |  |  |  |  |  |
| No | 294,316 (99.6%) | 147,468 (99.6%) | 12,374 (99.3%) | 6,164 (99.3%) | 306,690 (99.6%) | 153,632 (99.6%) |
| Yes | 396 (0.1%) | 185 (0.1%) | 29 (0.2%) | 17 (0.3%) | 425 (0.1%) | 202 (0.1%) |
| Ignored | 326 (0.1%) | 146 (0.1%) | 64 (0.5%) | 28 (0.5%) | 390 (0.1%) | 174 (0.1%) |
| Missing | 510 (0.2%) | 257 (0.2%) | 0 (0%) | 0 (0%) | 510 (0.2%) | 257 (0.2%) |
| **Peptic acid disease** |  |  |  |  |  |  |
| No | 294,571 (99.7%) | 147,560 (99.7%) | 12,385 (99.3%) | 6,169 (99.4%) | 306,956 (99.7%) | 153,729 (99.7%) |
| Yes | 142 (0.0%) | 95 (0.1%) | 18 (0.1%) | 12 (0.2%) | 160 (0.1%) | 107 (0.1%) |
| Ignored | 325 (0.1%) | 144 (0.1%) | 64 (0.5%) | 28 (0.5%) | 389 (0.1%) | 172 (0.1%) |
| Missing | 510 (0.2%) | 257 (0.2%) | 0 (0%) | 0 (0%) | 510 (0.2%) | 257 (0.2%) |
| **Case outcome** |  |  |  |  |  |  |
| Recovered | 174,165 (58.9%) | 87,001 (58.8%) | 9,293 (74.5%) | 4,609 (74.2%) | 183,458 (59.6%) | 91,610 (59.4%) |
| Death (arbovirus) | 78 (0.0%) | 37 (0.0%) | 1 (0.0%) | 0 (0%) | 79 (0.0%) | 37 (0.0%) |
| Death (other causes) | 225 (0.1%) | 111 (0.1%) | 8 (0.1%) | 1 (0.0%) | 233 (0.1%) | 112 (0.1%) |
| Death (investigating) | 1 (0.0%) | 3 (0.0%) | 0 (0%) | 0 (0%) | 1 (0.0%) | 3 (0.0%) |
| Ignored | 97,995 (33.2%) | 49,383 (33.4%) | 3,069 (24.6%) | 1,532 (24.7%) | 101,064 (32.8%) | 50,915 (33.0%) |
| Missing | 23,084 (7.8%) | 11,521 (7.8%) | 96 (0.8%) | 67 (1.1%) | 23,180 (7.5%) | 11,588 (7.5%) |
| **Hospitalization** |  |  |  |  |  |  |
| No | 289,110 (97.8%) | 144,869 (97.8%) | 12,144 (97.4%) | 6,064 (97.7%) | 301,254 (97.8%) | 150,933 (97.8%) |
| Yes | 5,859 (2.0%) | 2,902 (2.0%) | 316 (2.5%) | 144 (2.3%) | 6,175 (2.0%) | 3,046 (2.0%) |
| Ignored | 55 (0.0%) | 23 (0.0%) | 5 (0.0%) | 1 (0.0%) | 60 (0.0%) | 24 (0.0%) |
| Missing | 524 (0.2%) | 262 (0.2%) | 2 (0.0%) | 0 (0%) | 526 (0.2%) | 262 (0.2%) |
| **Disease timing at presentation** |  |  |  |  |  |  |
| Acute | 295,356 (99.9%) | 147,968 (99.9%) | 10,622 (85.2%) | 5,265 (84.8%) | 305,978 (99.3%) | 153,233 (99.3%) |
| Chronic | 192 (0.1%) | 88 (0.1%) | 460 (3.7%) | 224 (3.6%) | 652 (0.2%) | 312 (0.2%) |
| Missing | 0 (0%) | 0 (0%) | 1,385 (11.1%) | 720 (11.6%) | 1,385 (0.4%) | 720 (0.5%) |
| IQR = interquartile range; SD = Standard deviation. | | | | | | |

**Table S 6:** Final dataset variables (including epidemiological, clinical at presentations and disease progressions) counts and frequencies by zika diagnostic investigation outcome.

|  | **No** | | **Yes** | | **Overall** | |
| --- | --- | --- | --- | --- | --- | --- |
|  | **Train (N=307531)** | **Test (N=153991)** | **Train (N=484)** | **Test (N=274)** | **Train (N=308015)** | **Test (N=154265)** |
| **Age at notification** |  |  |  |  |  |  |
| Mean (SD) | 35.2 (19.1) | 35.1 (19.0) | 40.7 (17.9) | 39.0 (18.2) | 35.2 (19.1) | 35.1 (19.0) |
| Median [Min, Max] | 33.0 [0, 106] | 33.0 [0, 104] | 41.0 [0, 90.0] | 40.0 [0, 79.0] | 33.0 [0, 106] | 33.0 [0, 104] |
| Missing | 7,276 (2.4%) | 3,701 (2.4%) | 32 (6.6%) | 25 (9.1%) | 7,308 (2.4%) | 3,726 (2.4%) |
| **Age at notification** |  |  |  |  |  |  |
| [0,1] | 3,492 (1.1%) | 1,825 (1.2%) | 1 (0.2%) | 2 (0.7%) | 3,493 (1.1%) | 1,827 (1.2%) |
| (1,12] | 29,862 (9.7%) | 15,049 (9.8%) | 29 (6.0%) | 17 (6.2%) | 29,891 (9.7%) | 15,066 (9.8%) |
| (12,18] | 28,330 (9.2%) | 14,279 (9.3%) | 28 (5.8%) | 22 (8.0%) | 28,358 (9.2%) | 14,301 (9.3%) |
| (18,25] | 44,942 (14.6%) | 22,680 (14.7%) | 41 (8.5%) | 25 (9.1%) | 44,983 (14.6%) | 22,705 (14.7%) |
| (25,35] | 55,362 (18.0%) | 27,843 (18.1%) | 77 (15.9%) | 38 (13.9%) | 55,439 (18.0%) | 27,881 (18.1%) |
| (35,45] | 51,106 (16.6%) | 25,556 (16.6%) | 96 (19.8%) | 53 (19.3%) | 51,202 (16.6%) | 25,609 (16.6%) |
| (45,55] | 37,791 (12.3%) | 18,702 (12.1%) | 73 (15.1%) | 40 (14.6%) | 37,864 (12.3%) | 18,742 (12.1%) |
| (55,65] | 26,997 (8.8%) | 13,358 (8.7%) | 69 (14.3%) | 37 (13.5%) | 27,066 (8.8%) | 13,395 (8.7%) |
| (65,75] | 15,328 (5.0%) | 7,506 (4.9%) | 29 (6.0%) | 11 (4.0%) | 15,357 (5.0%) | 7,517 (4.9%) |
| (75,85] | 5,642 (1.8%) | 2,764 (1.8%) | 8 (1.7%) | 4 (1.5%) | 5,650 (1.8%) | 2,768 (1.8%) |
| (85,95] | 1,343 (0.4%) | 683 (0.4%) | 1 (0.2%) | 0 (0%) | 1,344 (0.4%) | 683 (0.4%) |
| Missing | 7,336 (2.4%) | 3,746 (2.4%) | 32 (6.6%) | 25 (9.1%) | 7,368 (2.4%) | 3,771 (2.4%) |
| **Sex at notification** |  |  |  |  |  |  |
| Female | 162,510 (52.8%) | 81,474 (52.9%) | 262 (54.1%) | 137 (50.0%) | 162,772 (52.8%) | 81,611 (52.9%) |
| Male | 137,733 (44.8%) | 68,811 (44.7%) | 190 (39.3%) | 112 (40.9%) | 137,923 (44.8%) | 68,923 (44.7%) |
| Ignored | 12 (0.0%) | 5 (0.0%) | 0 (0%) | 0 (0%) | 12 (0.0%) | 5 (0.0%) |
| Missing | 7,276 (2.4%) | 3,701 (2.4%) | 32 (6.6%) | 25 (9.1%) | 7,308 (2.4%) | 3,726 (2.4%) |
| **Pregnancy** |  |  |  |  |  |  |
| No | 268,824 (87.4%) | 134,313 (87.2%) | 421 (87.0%) | 221 (80.7%) | 269,245 (87.4%) | 134,534 (87.2%) |
| Yes | 3,338 (1.1%) | 1,744 (1.1%) | 13 (2.7%) | 11 (4.0%) | 3,351 (1.1%) | 1,755 (1.1%) |
| Ignored/Unknown | 28,093 (9.1%) | 14,233 (9.2%) | 18 (3.7%) | 17 (6.2%) | 28,111 (9.1%) | 14,250 (9.2%) |
| Missing | 7,276 (2.4%) | 3,701 (2.4%) | 32 (6.6%) | 25 (9.1%) | 7,308 (2.4%) | 3,726 (2.4%) |
| **Race or skin color** |  |  |  |  |  |  |
| White | 92,435 (30.1%) | 46,300 (30.1%) | 195 (40.3%) | 95 (34.7%) | 92,630 (30.1%) | 46,395 (30.1%) |
| Black | 160,475 (52.2%) | 80,354 (52.2%) | 157 (32.4%) | 93 (33.9%) | 160,632 (52.2%) | 80,447 (52.1%) |
| Yellow | 27,439 (8.9%) | 13,726 (8.9%) | 45 (9.3%) | 31 (11.3%) | 27,484 (8.9%) | 13,757 (8.9%) |
| Indigenous | 281 (0.1%) | 148 (0.1%) | 0 (0%) | 0 (0%) | 281 (0.1%) | 148 (0.1%) |
| Ignored/Unknown | 19,625 (6.4%) | 9,762 (6.3%) | 55 (11.4%) | 30 (10.9%) | 19,680 (6.4%) | 9,792 (6.3%) |
| Missing | 7,276 (2.4%) | 3,701 (2.4%) | 32 (6.6%) | 25 (9.1%) | 7,308 (2.4%) | 3,726 (2.4%) |
| **Indigenous ethnicity** |  |  |  |  |  |  |
| Tupiniquim | 113 (0.0%) | 49 (0.0%) | 0 (0%) | 0 (0%) | 113 (0.0%) | 49 (0.0%) |
| Guarani | 25 (0.0%) | 18 (0.0%) | 0 (0%) | 0 (0%) | 25 (0.0%) | 18 (0.0%) |
| Ignored/Unknown | 143 (0.0%) | 81 (0.1%) | 0 (0%) | 0 (0%) | 143 (0.0%) | 81 (0.1%) |
| Not applicable | 299,974 (97.5%) | 150,142 (97.5%) | 452 (93.4%) | 249 (90.9%) | 300,426 (97.5%) | 150,391 (97.5%) |
| Missing | 7,276 (2.4%) | 3,701 (2.4%) | 32 (6.6%) | 25 (9.1%) | 7,308 (2.4%) | 3,726 (2.4%) |
| **Person with special needs** |  |  |  |  |  |  |
| Yes | 307 (0.1%) | 157 (0.1%) | 0 (0%) | 1 (0.4%) | 307 (0.1%) | 158 (0.1%) |
| No | 299,948 (97.5%) | 150,133 (97.5%) | 452 (93.4%) | 248 (90.5%) | 300,400 (97.5%) | 150,381 (97.5%) |
| Missing | 7,276 (2.4%) | 3,701 (2.4%) | 32 (6.6%) | 25 (9.1%) | 7,308 (2.4%) | 3,726 (2.4%) |
| **Homeless** |  |  |  |  |  |  |
| Yes | 102 (0.0%) | 46 (0.0%) | 0 (0%) | 0 (0%) | 102 (0.0%) | 46 (0.0%) |
| No | 300,153 (97.6%) | 150,244 (97.6%) | 452 (93.4%) | 249 (90.9%) | 300,605 (97.6%) | 150,493 (97.6%) |
| Missing | 7,276 (2.4%) | 3,701 (2.4%) | 32 (6.6%) | 25 (9.1%) | 7,308 (2.4%) | 3,726 (2.4%) |
| **Education** |  |  |  |  |  |  |
| Illiterate | 14,029 (4.6%) | 6,881 (4.5%) | 37 (7.6%) | 15 (5.5%) | 14,066 (4.6%) | 6,896 (4.5%) |
| Elementary School Complete | 31,079 (10.1%) | 15,497 (10.1%) | 62 (12.8%) | 47 (17.2%) | 31,141 (10.1%) | 15,544 (10.1%) |
| Middle School Complete | 28,351 (9.2%) | 14,133 (9.2%) | 53 (11.0%) | 19 (6.9%) | 28,404 (9.2%) | 14,152 (9.2%) |
| High School Complete | 52,615 (17.1%) | 26,370 (17.1%) | 108 (22.3%) | 59 (21.5%) | 52,723 (17.1%) | 26,429 (17.1%) |
| Higher Education Complete | 14,561 (4.7%) | 7,240 (4.7%) | 60 (12.4%) | 29 (10.6%) | 14,621 (4.7%) | 7,269 (4.7%) |
| Ignored/Unknown | 159,620 (51.9%) | 80,169 (52.1%) | 132 (27.3%) | 80 (29.2%) | 159,752 (51.9%) | 80,249 (52.0%) |
| Missing | 7,276 (2.4%) | 3,701 (2.4%) | 32 (6.6%) | 25 (9.1%) | 7,308 (2.4%) | 3,726 (2.4%) |
| **Home address** |  |  |  |  |  |  |
| Urban | 234,449 (76.2%) | 117,129 (76.1%) | 297 (61.4%) | 170 (62.0%) | 234,746 (76.2%) | 117,299 (76.0%) |
| Rural | 45,503 (14.8%) | 22,886 (14.9%) | 100 (20.7%) | 59 (21.5%) | 45,603 (14.8%) | 22,945 (14.9%) |
| Peri-urban | 933 (0.3%) | 490 (0.3%) | 4 (0.8%) | 2 (0.7%) | 937 (0.3%) | 492 (0.3%) |
| Unknown | 19,367 (6.3%) | 9,782 (6.4%) | 51 (10.5%) | 18 (6.6%) | 19,418 (6.3%) | 9,800 (6.4%) |
| Missing | 7,279 (2.4%) | 3,704 (2.4%) | 32 (6.6%) | 25 (9.1%) | 7,311 (2.4%) | 3,729 (2.4%) |
| **Autochthonous case** |  |  |  |  |  |  |
| No | 509 (0.2%) | 217 (0.1%) | 5 (1.0%) | 1 (0.4%) | 514 (0.2%) | 218 (0.1%) |
| Yes | 305,978 (99.5%) | 153,248 (99.5%) | 449 (92.8%) | 254 (92.7%) | 306,427 (99.5%) | 153,502 (99.5%) |
| Unknown | 333 (0.1%) | 183 (0.1%) | 3 (0.6%) | 1 (0.4%) | 336 (0.1%) | 184 (0.1%) |
| Missing | 711 (0.2%) | 343 (0.2%) | 27 (5.6%) | 18 (6.6%) | 738 (0.2%) | 361 (0.2%) |
| **Fever** |  |  |  |  |  |  |
| No | 71,310 (23.2%) | 35,714 (23.2%) | 82 (16.9%) | 54 (19.7%) | 71,392 (23.2%) | 35,768 (23.2%) |
| Yes | 235,738 (76.7%) | 118,038 (76.7%) | 375 (77.5%) | 202 (73.7%) | 236,113 (76.7%) | 118,240 (76.6%) |
| Missing | 483 (0.2%) | 239 (0.2%) | 27 (5.6%) | 18 (6.6%) | 510 (0.2%) | 257 (0.2%) |
| **Headache** |  |  |  |  |  |  |
| No | 73,741 (24.0%) | 36,859 (23.9%) | 96 (19.8%) | 43 (15.7%) | 73,837 (24.0%) | 36,902 (23.9%) |
| Yes | 233,307 (75.9%) | 116,893 (75.9%) | 361 (74.6%) | 213 (77.7%) | 233,668 (75.9%) | 117,106 (75.9%) |
| Missing | 483 (0.2%) | 239 (0.2%) | 27 (5.6%) | 18 (6.6%) | 510 (0.2%) | 257 (0.2%) |
| **Vomiting** |  |  |  |  |  |  |
| No | 243,404 (79.1%) | 122,014 (79.2%) | 335 (69.2%) | 189 (69.0%) | 243,739 (79.1%) | 122,203 (79.2%) |
| Yes | 63,644 (20.7%) | 31,738 (20.6%) | 122 (25.2%) | 67 (24.5%) | 63,766 (20.7%) | 31,805 (20.6%) |
| Missing | 483 (0.2%) | 239 (0.2%) | 27 (5.6%) | 18 (6.6%) | 510 (0.2%) | 257 (0.2%) |
| **Back pain** |  |  |  |  |  |  |
| No | 242,216 (78.8%) | 121,309 (78.8%) | 305 (63.0%) | 177 (64.6%) | 242,521 (78.7%) | 121,486 (78.8%) |
| Yes | 64,832 (21.1%) | 32,443 (21.1%) | 152 (31.4%) | 79 (28.8%) | 64,984 (21.1%) | 32,522 (21.1%) |
| Missing | 483 (0.2%) | 239 (0.2%) | 27 (5.6%) | 18 (6.6%) | 510 (0.2%) | 257 (0.2%) |
| **Arthritis** |  |  |  |  |  |  |
| No | 286,131 (93.0%) | 143,432 (93.1%) | 401 (82.9%) | 220 (80.3%) | 286,532 (93.0%) | 143,652 (93.1%) |
| Yes | 20,917 (6.8%) | 10,320 (6.7%) | 56 (11.6%) | 36 (13.1%) | 20,973 (6.8%) | 10,356 (6.7%) |
| Missing | 483 (0.2%) | 239 (0.2%) | 27 (5.6%) | 18 (6.6%) | 510 (0.2%) | 257 (0.2%) |
| **Petechiae** |  |  |  |  |  |  |
| No | 292,890 (95.2%) | 146,856 (95.4%) | 381 (78.7%) | 223 (81.4%) | 293,271 (95.2%) | 147,079 (95.3%) |
| Yes | 14,158 (4.6%) | 6,896 (4.5%) | 76 (15.7%) | 33 (12.0%) | 14,234 (4.6%) | 6,929 (4.5%) |
| Missing | 483 (0.2%) | 239 (0.2%) | 27 (5.6%) | 18 (6.6%) | 510 (0.2%) | 257 (0.2%) |
| **Tourniquet test** |  |  |  |  |  |  |
| No | 303,135 (98.6%) | 151,923 (98.7%) | 448 (92.6%) | 248 (90.5%) | 303,583 (98.6%) | 152,171 (98.6%) |
| Yes | 3,913 (1.3%) | 1,829 (1.2%) | 9 (1.9%) | 8 (2.9%) | 3,922 (1.3%) | 1,837 (1.2%) |
| Missing | 483 (0.2%) | 239 (0.2%) | 27 (5.6%) | 18 (6.6%) | 510 (0.2%) | 257 (0.2%) |
| **Myalgia** |  |  |  |  |  |  |
| No | 84,698 (27.5%) | 42,531 (27.6%) | 89 (18.4%) | 48 (17.5%) | 84,787 (27.5%) | 42,579 (27.6%) |
| Yes | 222,350 (72.3%) | 111,221 (72.2%) | 368 (76.0%) | 208 (75.9%) | 222,718 (72.3%) | 111,429 (72.2%) |
| Missing | 483 (0.2%) | 239 (0.2%) | 27 (5.6%) | 18 (6.6%) | 510 (0.2%) | 257 (0.2%) |
| **Rash** |  |  |  |  |  |  |
| No | 293,676 (95.5%) | 147,019 (95.5%) | 400 (82.6%) | 225 (82.1%) | 294,076 (95.5%) | 147,244 (95.4%) |
| Yes | 13,372 (4.3%) | 6,733 (4.4%) | 57 (11.8%) | 31 (11.3%) | 13,429 (4.4%) | 6,764 (4.4%) |
| Missing | 483 (0.2%) | 239 (0.2%) | 27 (5.6%) | 18 (6.6%) | 510 (0.2%) | 257 (0.2%) |
| **Nausea** |  |  |  |  |  |  |
| No | 200,063 (65.1%) | 100,279 (65.1%) | 235 (48.6%) | 149 (54.4%) | 200,298 (65.0%) | 100,428 (65.1%) |
| Yes | 106,985 (34.8%) | 53,473 (34.7%) | 222 (45.9%) | 107 (39.1%) | 107,207 (34.8%) | 53,580 (34.7%) |
| Missing | 483 (0.2%) | 239 (0.2%) | 27 (5.6%) | 18 (6.6%) | 510 (0.2%) | 257 (0.2%) |
| **Conjunctivitis** |  |  |  |  |  |  |
| No | 303,947 (98.8%) | 152,254 (98.9%) | 451 (93.2%) | 250 (91.2%) | 304,398 (98.8%) | 152,504 (98.9%) |
| Yes | 3,101 (1.0%) | 1,498 (1.0%) | 6 (1.2%) | 6 (2.2%) | 3,107 (1.0%) | 1,504 (1.0%) |
| Missing | 483 (0.2%) | 239 (0.2%) | 27 (5.6%) | 18 (6.6%) | 510 (0.2%) | 257 (0.2%) |
| **Intense arthralgia** |  |  |  |  |  |  |
| No | 244,586 (79.5%) | 122,543 (79.6%) | 336 (69.4%) | 191 (69.7%) | 244,922 (79.5%) | 122,734 (79.6%) |
| Yes | 62,462 (20.3%) | 31,209 (20.3%) | 121 (25.0%) | 65 (23.7%) | 62,583 (20.3%) | 31,274 (20.3%) |
| Missing | 483 (0.2%) | 239 (0.2%) | 27 (5.6%) | 18 (6.6%) | 510 (0.2%) | 257 (0.2%) |
| **Leukopenia** |  |  |  |  |  |  |
| No | 301,480 (98.0%) | 150,948 (98.0%) | 440 (90.9%) | 246 (89.8%) | 301,920 (98.0%) | 151,194 (98.0%) |
| Yes | 5,568 (1.8%) | 2,804 (1.8%) | 17 (3.5%) | 10 (3.6%) | 5,585 (1.8%) | 2,814 (1.8%) |
| Missing | 483 (0.2%) | 239 (0.2%) | 27 (5.6%) | 18 (6.6%) | 510 (0.2%) | 257 (0.2%) |
| **Retro-orbital pain** |  |  |  |  |  |  |
| No | 213,974 (69.6%) | 107,005 (69.5%) | 261 (53.9%) | 155 (56.6%) | 214,235 (69.6%) | 107,160 (69.5%) |
| Yes | 93,074 (30.3%) | 46,747 (30.4%) | 196 (40.5%) | 101 (36.9%) | 93,270 (30.3%) | 46,848 (30.4%) |
| Missing | 483 (0.2%) | 239 (0.2%) | 27 (5.6%) | 18 (6.6%) | 510 (0.2%) | 257 (0.2%) |
| **Diabetes** |  |  |  |  |  |  |
| No | 297,893 (96.9%) | 149,212 (96.9%) | 440 (90.9%) | 248 (90.5%) | 298,333 (96.9%) | 149,460 (96.9%) |
| Yes | 8,752 (2.8%) | 4,368 (2.8%) | 17 (3.5%) | 8 (2.9%) | 8,769 (2.8%) | 4,376 (2.8%) |
| Ignored | 403 (0.1%) | 172 (0.1%) | 0 (0%) | 0 (0%) | 403 (0.1%) | 172 (0.1%) |
| Missing | 483 (0.2%) | 239 (0.2%) | 27 (5.6%) | 18 (6.6%) | 510 (0.2%) | 257 (0.2%) |
| **Hepatopathies** |  |  |  |  |  |  |
| No | 306,299 (99.6%) | 153,390 (99.6%) | 456 (94.2%) | 256 (93.4%) | 306,755 (99.6%) | 153,646 (99.6%) |
| Yes | 349 (0.1%) | 185 (0.1%) | 1 (0.2%) | 0 (0%) | 350 (0.1%) | 185 (0.1%) |
| Ignored | 400 (0.1%) | 177 (0.1%) | 0 (0%) | 0 (0%) | 400 (0.1%) | 177 (0.1%) |
| Missing | 483 (0.2%) | 239 (0.2%) | 27 (5.6%) | 18 (6.6%) | 510 (0.2%) | 257 (0.2%) |
| **Arterial hypertension** |  |  |  |  |  |  |
| No | 283,826 (92.3%) | 142,325 (92.4%) | 399 (82.4%) | 223 (81.4%) | 284,225 (92.3%) | 142,548 (92.4%) |
| Yes | 22,824 (7.4%) | 11,255 (7.3%) | 58 (12.0%) | 33 (12.0%) | 22,882 (7.4%) | 11,288 (7.3%) |
| Ignored | 398 (0.1%) | 172 (0.1%) | 0 (0%) | 0 (0%) | 398 (0.1%) | 172 (0.1%) |
| Missing | 483 (0.2%) | 239 (0.2%) | 27 (5.6%) | 18 (6.6%) | 510 (0.2%) | 257 (0.2%) |
| **Autoimmune diseases** |  |  |  |  |  |  |
| No | 305,684 (99.4%) | 153,098 (99.4%) | 454 (93.8%) | 256 (93.4%) | 306,138 (99.4%) | 153,354 (99.4%) |
| Yes | 957 (0.3%) | 473 (0.3%) | 3 (0.6%) | 0 (0%) | 960 (0.3%) | 473 (0.3%) |
| Ignored | 407 (0.1%) | 181 (0.1%) | 0 (0%) | 0 (0%) | 407 (0.1%) | 181 (0.1%) |
| Missing | 483 (0.2%) | 239 (0.2%) | 27 (5.6%) | 18 (6.6%) | 510 (0.2%) | 257 (0.2%) |
| **Hematological diseases** |  |  |  |  |  |  |
| No | 306,370 (99.6%) | 153,420 (99.6%) | 457 (94.4%) | 256 (93.4%) | 306,827 (99.6%) | 153,676 (99.6%) |
| Yes | 286 (0.1%) | 160 (0.1%) | 0 (0%) | 0 (0%) | 286 (0.1%) | 160 (0.1%) |
| Ignored | 392 (0.1%) | 172 (0.1%) | 0 (0%) | 0 (0%) | 392 (0.1%) | 172 (0.1%) |
| Missing | 483 (0.2%) | 239 (0.2%) | 27 (5.6%) | 18 (6.6%) | 510 (0.2%) | 257 (0.2%) |
| **Chronic kidney disease** |  |  |  |  |  |  |
| No | 306,233 (99.6%) | 153,377 (99.6%) | 457 (94.4%) | 255 (93.1%) | 306,690 (99.6%) | 153,632 (99.6%) |
| Yes | 425 (0.1%) | 201 (0.1%) | 0 (0%) | 1 (0.4%) | 425 (0.1%) | 202 (0.1%) |
| Ignored | 390 (0.1%) | 174 (0.1%) | 0 (0%) | 0 (0%) | 390 (0.1%) | 174 (0.1%) |
| Missing | 483 (0.2%) | 239 (0.2%) | 27 (5.6%) | 18 (6.6%) | 510 (0.2%) | 257 (0.2%) |
| **Peptic acid disease** |  |  |  |  |  |  |
| No | 306,499 (99.7%) | 153,473 (99.7%) | 457 (94.4%) | 256 (93.4%) | 306,956 (99.7%) | 153,729 (99.7%) |
| Yes | 160 (0.1%) | 107 (0.1%) | 0 (0%) | 0 (0%) | 160 (0.1%) | 107 (0.1%) |
| Ignored | 389 (0.1%) | 172 (0.1%) | 0 (0%) | 0 (0%) | 389 (0.1%) | 172 (0.1%) |
| Missing | 483 (0.2%) | 239 (0.2%) | 27 (5.6%) | 18 (6.6%) | 510 (0.2%) | 257 (0.2%) |
| **Case outcome** |  |  |  |  |  |  |
| Recovered | 183,084 (59.5%) | 91,402 (59.4%) | 374 (77.3%) | 208 (75.9%) | 183,458 (59.6%) | 91,610 (59.4%) |
| Death (arbovirus) | 79 (0.0%) | 37 (0.0%) | 0 (0%) | 0 (0%) | 79 (0.0%) | 37 (0.0%) |
| Death (other causes) | 233 (0.1%) | 112 (0.1%) | 0 (0%) | 0 (0%) | 233 (0.1%) | 112 (0.1%) |
| Death (investigating) | 1 (0.0%) | 3 (0.0%) | 0 (0%) | 0 (0%) | 1 (0.0%) | 3 (0.0%) |
| Ignored | 100,993 (32.8%) | 50,877 (33.0%) | 71 (14.7%) | 38 (13.9%) | 101,064 (32.8%) | 50,915 (33.0%) |
| Missing | 23,141 (7.5%) | 11,560 (7.5%) | 39 (8.1%) | 28 (10.2%) | 23,180 (7.5%) | 11,588 (7.5%) |
| **Hospitalization** |  |  |  |  |  |  |
| No | 300,814 (97.8%) | 150,688 (97.9%) | 440 (90.9%) | 245 (89.4%) | 301,254 (97.8%) | 150,933 (97.8%) |
| Yes | 6,159 (2.0%) | 3,035 (2.0%) | 16 (3.3%) | 11 (4.0%) | 6,175 (2.0%) | 3,046 (2.0%) |
| Ignored | 59 (0.0%) | 24 (0.0%) | 1 (0.2%) | 0 (0%) | 60 (0.0%) | 24 (0.0%) |
| Missing | 499 (0.2%) | 244 (0.2%) | 27 (5.6%) | 18 (6.6%) | 526 (0.2%) | 262 (0.2%) |
| IQR = interquartile range; SD = Standard deviation in each dataset split. | | | | | | |

**Table S 7** Final dataset variables (including epidemiological, clinical at presentations and disease progressions) counts and frequencies by oropouche diagnostic investigation outcome in each dataset split.

|  | **Train** | | **Test** | | **Overall** | |
| --- | --- | --- | --- | --- | --- | --- |
|  | **No (N=299887)** | **Yes (N=8128)** | **No (N=150219)** | **Yes (N=4046)** | **No (N=450106)** | **Yes (N=12174)** |
| **Age at notification** |  |  |  |  |  |  |
| Mean (SD) | 35.1 (19.0) | 41.9 (18.1) | 34.9 (19.0) | 42.1 (18.5) | 35.0 (19.0) | 42.0 (18.2) |
| Median [Min, Max] | 33.0 [0, 106] | 41.0 [0, 96.0] | 33.0 [0, 104] | 42.0 [0, 99.0] | 33.0 [0, 106] | 41.0 [0, 99.0] |
| Missing | 7,031 (2.3%) | 277 (3.4%) | 3,591 (2.4%) | 135 (3.3%) | 10,622 (2.4%) | 412 (3.4%) |
| **Age at notification** |  |  |  |  |  |  |
| [0,1] | 3,487 (1.2%) | 6 (0.1%) | 1,823 (1.2%) | 4 (0.1%) | 5,310 (1.2%) | 10 (0.1%) |
| (1,12] | 29,597 (9.9%) | 294 (3.6%) | 14,912 (9.9%) | 154 (3.8%) | 44,509 (9.9%) | 448 (3.7%) |
| (12,18] | 27,841 (9.3%) | 517 (6.4%) | 14,048 (9.4%) | 253 (6.3%) | 41,889 (9.3%) | 770 (6.3%) |
| (18,25] | 44,148 (14.7%) | 835 (10.3%) | 22,270 (14.8%) | 435 (10.8%) | 66,418 (14.8%) | 1,270 (10.4%) |
| (25,35] | 54,049 (18.0%) | 1,390 (17.1%) | 27,233 (18.1%) | 648 (16.0%) | 81,282 (18.1%) | 2,038 (16.7%) |
| (35,45] | 49,646 (16.6%) | 1,556 (19.1%) | 24,852 (16.5%) | 757 (18.7%) | 74,498 (16.6%) | 2,313 (19.0%) |
| (45,55] | 36,556 (12.2%) | 1,308 (16.1%) | 18,090 (12.0%) | 652 (16.1%) | 54,646 (12.1%) | 1,960 (16.1%) |
| (55,65] | 26,003 (8.7%) | 1,063 (13.1%) | 12,839 (8.5%) | 556 (13.7%) | 38,842 (8.6%) | 1,619 (13.3%) |
| (65,75] | 14,737 (4.9%) | 620 (7.6%) | 7,207 (4.8%) | 310 (7.7%) | 21,944 (4.9%) | 930 (7.6%) |
| (75,85] | 5,428 (1.8%) | 222 (2.7%) | 2,659 (1.8%) | 109 (2.7%) | 8,087 (1.8%) | 331 (2.7%) |
| (85,95] | 1,305 (0.4%) | 39 (0.5%) | 652 (0.4%) | 31 (0.8%) | 1,957 (0.4%) | 70 (0.6%) |
| Missing | 7,090 (2.4%) | 278 (3.4%) | 3,634 (2.4%) | 137 (3.4%) | 10,724 (2.4%) | 415 (3.4%) |
| **Sex at notification** |  |  |  |  |  |  |
| Female | 159,169 (53.1%) | 3,603 (44.3%) | 79,766 (53.1%) | 1,845 (45.6%) | 238,935 (53.1%) | 5,448 (44.8%) |
| Male | 133,675 (44.6%) | 4,248 (52.3%) | 66,858 (44.5%) | 2,065 (51.0%) | 200,533 (44.6%) | 6,313 (51.9%) |
| Ignored | 12 (0.0%) | 0 (0%) | 4 (0.0%) | 1 (0.0%) | 16 (0.0%) | 1 (0.0%) |
| Missing | 7,031 (2.3%) | 277 (3.4%) | 3,591 (2.4%) | 135 (3.3%) | 10,622 (2.4%) | 412 (3.4%) |
| **Pregnancy** |  |  |  |  |  |  |
| No | 261,909 (87.3%) | 7,336 (90.3%) | 130,866 (87.1%) | 3,668 (90.7%) | 392,775 (87.3%) | 11,004 (90.4%) |
| Yes | 3,172 (1.1%) | 179 (2.2%) | 1,688 (1.1%) | 67 (1.7%) | 4,860 (1.1%) | 246 (2.0%) |
| Ignored/Unknown | 27,775 (9.3%) | 336 (4.1%) | 14,074 (9.4%) | 176 (4.4%) | 41,849 (9.3%) | 512 (4.2%) |
| Missing | 7,031 (2.3%) | 277 (3.4%) | 3,591 (2.4%) | 135 (3.3%) | 10,622 (2.4%) | 412 (3.4%) |
| **Race or skin color** |  |  |  |  |  |  |
| White | 88,236 (29.4%) | 4,394 (54.1%) | 44,194 (29.4%) | 2,201 (54.4%) | 132,430 (29.4%) | 6,595 (54.2%) |
| Black | 158,051 (52.7%) | 2,581 (31.8%) | 79,148 (52.7%) | 1,299 (32.1%) | 237,199 (52.7%) | 3,880 (31.9%) |
| Yellow | 26,615 (8.9%) | 869 (10.7%) | 13,347 (8.9%) | 410 (10.1%) | 39,962 (8.9%) | 1,279 (10.5%) |
| Indigenous | 274 (0.1%) | 7 (0.1%) | 147 (0.1%) | 1 (0.0%) | 421 (0.1%) | 8 (0.1%) |
| Ignored/Unknown | 19,680 (6.6%) | 0 (0%) | 9,792 (6.5%) | 0 (0%) | 29,472 (6.5%) | 0 (0%) |
| Missing | 7,031 (2.3%) | 277 (3.4%) | 3,591 (2.4%) | 135 (3.3%) | 10,622 (2.4%) | 412 (3.4%) |
| **Indigenous ethnicity** |  |  |  |  |  |  |
| Tupiniquim | 112 (0.0%) | 1 (0.0%) | 49 (0.0%) | 0 (0%) | 161 (0.0%) | 1 (0.0%) |
| Guarani | 25 (0.0%) | 0 (0%) | 18 (0.0%) | 0 (0%) | 43 (0.0%) | 0 (0%) |
| Ignored/Unknown | 137 (0.0%) | 6 (0.1%) | 80 (0.1%) | 1 (0.0%) | 217 (0.0%) | 7 (0.1%) |
| Not applicable | 292,582 (97.6%) | 7,844 (96.5%) | 146,481 (97.5%) | 3,910 (96.6%) | 439,063 (97.5%) | 11,754 (96.6%) |
| Missing | 7,031 (2.3%) | 277 (3.4%) | 3,591 (2.4%) | 135 (3.3%) | 10,622 (2.4%) | 412 (3.4%) |
| **Person with special needs** |  |  |  |  |  |  |
| Yes | 307 (0.1%) | 0 (0%) | 157 (0.1%) | 1 (0.0%) | 464 (0.1%) | 1 (0.0%) |
| No | 292,549 (97.6%) | 7,851 (96.6%) | 146,471 (97.5%) | 3,910 (96.6%) | 439,020 (97.5%) | 11,761 (96.6%) |
| Missing | 7,031 (2.3%) | 277 (3.4%) | 3,591 (2.4%) | 135 (3.3%) | 10,622 (2.4%) | 412 (3.4%) |
| **Homeless** |  |  |  |  |  |  |
| Yes | 102 (0.0%) | 0 (0%) | 46 (0.0%) | 0 (0%) | 148 (0.0%) | 0 (0%) |
| No | 292,754 (97.6%) | 7,851 (96.6%) | 146,582 (97.6%) | 3,911 (96.7%) | 439,336 (97.6%) | 11,762 (96.6%) |
| Missing | 7,031 (2.3%) | 277 (3.4%) | 3,591 (2.4%) | 135 (3.3%) | 10,622 (2.4%) | 412 (3.4%) |
| **Education** |  |  |  |  |  |  |
| Illiterate | 13,443 (4.5%) | 623 (7.7%) | 6,563 (4.4%) | 333 (8.2%) | 20,006 (4.4%) | 956 (7.9%) |
| Elementary School Complete | 29,774 (9.9%) | 1,367 (16.8%) | 14,834 (9.9%) | 710 (17.5%) | 44,608 (9.9%) | 2,077 (17.1%) |
| Middle School Complete | 27,381 (9.1%) | 1,023 (12.6%) | 13,665 (9.1%) | 487 (12.0%) | 41,046 (9.1%) | 1,510 (12.4%) |
| High School Complete | 50,932 (17.0%) | 1,791 (22.0%) | 25,536 (17.0%) | 893 (22.1%) | 76,468 (17.0%) | 2,684 (22.0%) |
| Higher Education Complete | 14,119 (4.7%) | 502 (6.2%) | 7,042 (4.7%) | 227 (5.6%) | 21,161 (4.7%) | 729 (6.0%) |
| Ignored/Unknown | 157,207 (52.4%) | 2,545 (31.3%) | 78,988 (52.6%) | 1,261 (31.2%) | 236,195 (52.5%) | 3,806 (31.3%) |
| Missing | 7,031 (2.3%) | 277 (3.4%) | 3,591 (2.4%) | 135 (3.3%) | 10,622 (2.4%) | 412 (3.4%) |
| **Home address** |  |  |  |  |  |  |
| Urban | 231,036 (77.0%) | 3,710 (45.6%) | 115,430 (76.8%) | 1,869 (46.2%) | 346,466 (77.0%) | 5,579 (45.8%) |
| Rural | 42,106 (14.0%) | 3,497 (43.0%) | 21,249 (14.1%) | 1,696 (41.9%) | 63,355 (14.1%) | 5,193 (42.7%) |
| Peri-urban | 892 (0.3%) | 45 (0.6%) | 469 (0.3%) | 23 (0.6%) | 1,361 (0.3%) | 68 (0.6%) |
| Unknown | 18,819 (6.3%) | 599 (7.4%) | 9,477 (6.3%) | 323 (8.0%) | 28,296 (6.3%) | 922 (7.6%) |
| Missing | 7,034 (2.3%) | 277 (3.4%) | 3,594 (2.4%) | 135 (3.3%) | 10,628 (2.4%) | 412 (3.4%) |
| **Autochthonous case** |  |  |  |  |  |  |
| No | 455 (0.2%) | 59 (0.7%) | 199 (0.1%) | 19 (0.5%) | 654 (0.1%) | 78 (0.6%) |
| Yes | 298,628 (99.6%) | 7,799 (96.0%) | 149,607 (99.6%) | 3,895 (96.3%) | 448,235 (99.6%) | 11,694 (96.1%) |
| Unknown | 327 (0.1%) | 9 (0.1%) | 180 (0.1%) | 4 (0.1%) | 507 (0.1%) | 13 (0.1%) |
| Missing | 477 (0.2%) | 261 (3.2%) | 233 (0.2%) | 128 (3.2%) | 710 (0.2%) | 389 (3.2%) |
| **Fever** |  |  |  |  |  |  |
| No | 70,217 (23.4%) | 1,175 (14.5%) | 35,180 (23.4%) | 588 (14.5%) | 105,397 (23.4%) | 1,763 (14.5%) |
| Yes | 229,418 (76.5%) | 6,695 (82.4%) | 114,909 (76.5%) | 3,331 (82.3%) | 344,327 (76.5%) | 10,026 (82.4%) |
| Missing | 252 (0.1%) | 258 (3.2%) | 130 (0.1%) | 127 (3.1%) | 382 (0.1%) | 385 (3.2%) |
| **Headache** |  |  |  |  |  |  |
| No | 72,531 (24.2%) | 1,306 (16.1%) | 36,221 (24.1%) | 681 (16.8%) | 108,752 (24.2%) | 1,987 (16.3%) |
| Yes | 227,104 (75.7%) | 6,564 (80.8%) | 113,868 (75.8%) | 3,238 (80.0%) | 340,972 (75.8%) | 9,802 (80.5%) |
| Missing | 252 (0.1%) | 258 (3.2%) | 130 (0.1%) | 127 (3.1%) | 382 (0.1%) | 385 (3.2%) |
| **Vomiting** |  |  |  |  |  |  |
| No | 237,024 (79.0%) | 6,715 (82.6%) | 118,868 (79.1%) | 3,335 (82.4%) | 355,892 (79.1%) | 10,050 (82.6%) |
| Yes | 62,611 (20.9%) | 1,155 (14.2%) | 31,221 (20.8%) | 584 (14.4%) | 93,832 (20.8%) | 1,739 (14.3%) |
| Missing | 252 (0.1%) | 258 (3.2%) | 130 (0.1%) | 127 (3.1%) | 382 (0.1%) | 385 (3.2%) |
| **Back pain** |  |  |  |  |  |  |
| No | 237,093 (79.1%) | 5,428 (66.8%) | 118,722 (79.0%) | 2,764 (68.3%) | 355,815 (79.1%) | 8,192 (67.3%) |
| Yes | 62,542 (20.9%) | 2,442 (30.0%) | 31,367 (20.9%) | 1,155 (28.5%) | 93,909 (20.9%) | 3,597 (29.5%) |
| Missing | 252 (0.1%) | 258 (3.2%) | 130 (0.1%) | 127 (3.1%) | 382 (0.1%) | 385 (3.2%) |
| **Arthritis** |  |  |  |  |  |  |
| No | 279,328 (93.1%) | 7,204 (88.6%) | 140,079 (93.2%) | 3,573 (88.3%) | 419,407 (93.2%) | 10,777 (88.5%) |
| Yes | 20,307 (6.8%) | 666 (8.2%) | 10,010 (6.7%) | 346 (8.6%) | 30,317 (6.7%) | 1,012 (8.3%) |
| Missing | 252 (0.1%) | 258 (3.2%) | 130 (0.1%) | 127 (3.1%) | 382 (0.1%) | 385 (3.2%) |
| **Petechiae** |  |  |  |  |  |  |
| No | 285,573 (95.2%) | 7,698 (94.7%) | 143,248 (95.4%) | 3,831 (94.7%) | 428,821 (95.3%) | 11,529 (94.7%) |
| Yes | 14,062 (4.7%) | 172 (2.1%) | 6,841 (4.6%) | 88 (2.2%) | 20,903 (4.6%) | 260 (2.1%) |
| Missing | 252 (0.1%) | 258 (3.2%) | 130 (0.1%) | 127 (3.1%) | 382 (0.1%) | 385 (3.2%) |
| **Tourniquet test** |  |  |  |  |  |  |
| No | 295,751 (98.6%) | 7,832 (96.4%) | 148,269 (98.7%) | 3,902 (96.4%) | 444,020 (98.6%) | 11,734 (96.4%) |
| Yes | 3,884 (1.3%) | 38 (0.5%) | 1,820 (1.2%) | 17 (0.4%) | 5,704 (1.3%) | 55 (0.5%) |
| Missing | 252 (0.1%) | 258 (3.2%) | 130 (0.1%) | 127 (3.1%) | 382 (0.1%) | 385 (3.2%) |
| **Myalgia** |  |  |  |  |  |  |
| No | 82,920 (27.7%) | 1,867 (23.0%) | 41,559 (27.7%) | 1,020 (25.2%) | 124,479 (27.7%) | 2,887 (23.7%) |
| Yes | 216,715 (72.3%) | 6,003 (73.9%) | 108,530 (72.2%) | 2,899 (71.7%) | 325,245 (72.3%) | 8,902 (73.1%) |
| Missing | 252 (0.1%) | 258 (3.2%) | 130 (0.1%) | 127 (3.1%) | 382 (0.1%) | 385 (3.2%) |
| **Rash** |  |  |  |  |  |  |
| No | 286,367 (95.5%) | 7,709 (94.8%) | 143,420 (95.5%) | 3,824 (94.5%) | 429,787 (95.5%) | 11,533 (94.7%) |
| Yes | 13,268 (4.4%) | 161 (2.0%) | 6,669 (4.4%) | 95 (2.3%) | 19,937 (4.4%) | 256 (2.1%) |
| Missing | 252 (0.1%) | 258 (3.2%) | 130 (0.1%) | 127 (3.1%) | 382 (0.1%) | 385 (3.2%) |
| **Nausea** |  |  |  |  |  |  |
| No | 195,407 (65.2%) | 4,891 (60.2%) | 97,986 (65.2%) | 2,442 (60.4%) | 293,393 (65.2%) | 7,333 (60.2%) |
| Yes | 104,228 (34.8%) | 2,979 (36.7%) | 52,103 (34.7%) | 1,477 (36.5%) | 156,331 (34.7%) | 4,456 (36.6%) |
| Missing | 252 (0.1%) | 258 (3.2%) | 130 (0.1%) | 127 (3.1%) | 382 (0.1%) | 385 (3.2%) |
| **Conjunctivitis** |  |  |  |  |  |  |
| No | 296,580 (98.9%) | 7,818 (96.2%) | 148,613 (98.9%) | 3,891 (96.2%) | 445,193 (98.9%) | 11,709 (96.2%) |
| Yes | 3,055 (1.0%) | 52 (0.6%) | 1,476 (1.0%) | 28 (0.7%) | 4,531 (1.0%) | 80 (0.7%) |
| Missing | 252 (0.1%) | 258 (3.2%) | 130 (0.1%) | 127 (3.1%) | 382 (0.1%) | 385 (3.2%) |
| **Intense arthralgia** |  |  |  |  |  |  |
| No | 238,414 (79.5%) | 6,508 (80.1%) | 119,409 (79.5%) | 3,325 (82.2%) | 357,823 (79.5%) | 9,833 (80.8%) |
| Yes | 61,221 (20.4%) | 1,362 (16.8%) | 30,680 (20.4%) | 594 (14.7%) | 91,901 (20.4%) | 1,956 (16.1%) |
| Missing | 252 (0.1%) | 258 (3.2%) | 130 (0.1%) | 127 (3.1%) | 382 (0.1%) | 385 (3.2%) |
| **Leukopenia** |  |  |  |  |  |  |
| No | 294,180 (98.1%) | 7,740 (95.2%) | 147,331 (98.1%) | 3,863 (95.5%) | 441,511 (98.1%) | 11,603 (95.3%) |
| Yes | 5,455 (1.8%) | 130 (1.6%) | 2,758 (1.8%) | 56 (1.4%) | 8,213 (1.8%) | 186 (1.5%) |
| Missing | 252 (0.1%) | 258 (3.2%) | 130 (0.1%) | 127 (3.1%) | 382 (0.1%) | 385 (3.2%) |
| **Retro-orbital pain** |  |  |  |  |  |  |
| No | 209,096 (69.7%) | 5,139 (63.2%) | 104,552 (69.6%) | 2,608 (64.5%) | 313,648 (69.7%) | 7,747 (63.6%) |
| Yes | 90,539 (30.2%) | 2,731 (33.6%) | 45,537 (30.3%) | 1,311 (32.4%) | 136,076 (30.2%) | 4,042 (33.2%) |
| Missing | 252 (0.1%) | 258 (3.2%) | 130 (0.1%) | 127 (3.1%) | 382 (0.1%) | 385 (3.2%) |
| **Diabetes** |  |  |  |  |  |  |
| No | 290,769 (97.0%) | 7,564 (93.1%) | 145,687 (97.0%) | 3,773 (93.3%) | 436,456 (97.0%) | 11,337 (93.1%) |
| Yes | 8,464 (2.8%) | 305 (3.8%) | 4,231 (2.8%) | 145 (3.6%) | 12,695 (2.8%) | 450 (3.7%) |
| Ignored | 402 (0.1%) | 1 (0.0%) | 171 (0.1%) | 1 (0.0%) | 573 (0.1%) | 2 (0.0%) |
| Missing | 252 (0.1%) | 258 (3.2%) | 130 (0.1%) | 127 (3.1%) | 382 (0.1%) | 385 (3.2%) |
| **Hepatopathies** |  |  |  |  |  |  |
| No | 298,896 (99.7%) | 7,859 (96.7%) | 149,734 (99.7%) | 3,912 (96.7%) | 448,630 (99.7%) | 11,771 (96.7%) |
| Yes | 340 (0.1%) | 10 (0.1%) | 179 (0.1%) | 6 (0.1%) | 519 (0.1%) | 16 (0.1%) |
| Ignored | 399 (0.1%) | 1 (0.0%) | 176 (0.1%) | 1 (0.0%) | 575 (0.1%) | 2 (0.0%) |
| Missing | 252 (0.1%) | 258 (3.2%) | 130 (0.1%) | 127 (3.1%) | 382 (0.1%) | 385 (3.2%) |
| **Arterial hypertension** |  |  |  |  |  |  |
| No | 277,182 (92.4%) | 7,043 (86.7%) | 139,045 (92.6%) | 3,503 (86.6%) | 416,227 (92.5%) | 10,546 (86.6%) |
| Yes | 22,056 (7.4%) | 826 (10.2%) | 10,873 (7.2%) | 415 (10.3%) | 32,929 (7.3%) | 1,241 (10.2%) |
| Ignored | 397 (0.1%) | 1 (0.0%) | 171 (0.1%) | 1 (0.0%) | 568 (0.1%) | 2 (0.0%) |
| Missing | 252 (0.1%) | 258 (3.2%) | 130 (0.1%) | 127 (3.1%) | 382 (0.1%) | 385 (3.2%) |
| **Autoimmune diseases** |  |  |  |  |  |  |
| No | 298,287 (99.5%) | 7,851 (96.6%) | 149,445 (99.5%) | 3,909 (96.6%) | 447,732 (99.5%) | 11,760 (96.6%) |
| Yes | 943 (0.3%) | 17 (0.2%) | 464 (0.3%) | 9 (0.2%) | 1,407 (0.3%) | 26 (0.2%) |
| Ignored | 405 (0.1%) | 2 (0.0%) | 180 (0.1%) | 1 (0.0%) | 585 (0.1%) | 3 (0.0%) |
| Missing | 252 (0.1%) | 258 (3.2%) | 130 (0.1%) | 127 (3.1%) | 382 (0.1%) | 385 (3.2%) |
| **Hematological diseases** |  |  |  |  |  |  |
| No | 298,964 (99.7%) | 7,863 (96.7%) | 149,761 (99.7%) | 3,915 (96.8%) | 448,725 (99.7%) | 11,778 (96.7%) |
| Yes | 280 (0.1%) | 6 (0.1%) | 157 (0.1%) | 3 (0.1%) | 437 (0.1%) | 9 (0.1%) |
| Ignored | 391 (0.1%) | 1 (0.0%) | 171 (0.1%) | 1 (0.0%) | 562 (0.1%) | 2 (0.0%) |
| Missing | 252 (0.1%) | 258 (3.2%) | 130 (0.1%) | 127 (3.1%) | 382 (0.1%) | 385 (3.2%) |
| **Chronic kidney disease** |  |  |  |  |  |  |
| No | 298,826 (99.6%) | 7,864 (96.8%) | 149,718 (99.7%) | 3,914 (96.7%) | 448,544 (99.7%) | 11,778 (96.7%) |
| Yes | 420 (0.1%) | 5 (0.1%) | 198 (0.1%) | 4 (0.1%) | 618 (0.1%) | 9 (0.1%) |
| Ignored | 389 (0.1%) | 1 (0.0%) | 173 (0.1%) | 1 (0.0%) | 562 (0.1%) | 2 (0.0%) |
| Missing | 252 (0.1%) | 258 (3.2%) | 130 (0.1%) | 127 (3.1%) | 382 (0.1%) | 385 (3.2%) |
| **Peptic acid disease** |  |  |  |  |  |  |
| No | 299,087 (99.7%) | 7,869 (96.8%) | 149,814 (99.7%) | 3,915 (96.8%) | 448,901 (99.7%) | 11,784 (96.8%) |
| Yes | 160 (0.1%) | 0 (0%) | 104 (0.1%) | 3 (0.1%) | 264 (0.1%) | 3 (0.0%) |
| Ignored | 388 (0.1%) | 1 (0.0%) | 171 (0.1%) | 1 (0.0%) | 559 (0.1%) | 2 (0.0%) |
| Missing | 252 (0.1%) | 258 (3.2%) | 130 (0.1%) | 127 (3.1%) | 382 (0.1%) | 385 (3.2%) |
| **Case outcome** |  |  |  |  |  |  |
| Recovered | 182,025 (60.7%) | 1,433 (17.6%) | 90,840 (60.5%) | 770 (19.0%) | 272,865 (60.6%) | 2,203 (18.1%) |
| Death (arbovirus) | 79 (0.0%) | 0 (0%) | 37 (0.0%) | 0 (0%) | 116 (0.0%) | 0 (0%) |
| Death (other causes) | 227 (0.1%) | 6 (0.1%) | 108 (0.1%) | 4 (0.1%) | 335 (0.1%) | 10 (0.1%) |
| Death (investigating) | 0 (0%) | 1 (0.0%) | 3 (0.0%) | 0 (0%) | 3 (0.0%) | 1 (0.0%) |
| Ignored | 97,376 (32.5%) | 3,688 (45.4%) | 49,039 (32.6%) | 1,876 (46.4%) | 146,415 (32.5%) | 5,564 (45.7%) |
| Missing | 20,180 (6.7%) | 3,000 (36.9%) | 10,192 (6.8%) | 1,396 (34.5%) | 30,372 (6.7%) | 4,396 (36.1%) |
| **Hospitalization** |  |  |  |  |  |  |
| No | 293,423 (97.8%) | 7,831 (96.3%) | 147,047 (97.9%) | 3,886 (96.0%) | 440,470 (97.9%) | 11,717 (96.2%) |
| Yes | 6,136 (2.0%) | 39 (0.5%) | 3,013 (2.0%) | 33 (0.8%) | 9,149 (2.0%) | 72 (0.6%) |
| Ignored | 60 (0.0%) | 0 (0%) | 24 (0.0%) | 0 (0%) | 84 (0.0%) | 0 (0%) |
| Missing | 268 (0.1%) | 258 (3.2%) | 135 (0.1%) | 127 (3.1%) | 403 (0.1%) | 385 (3.2%) |
| IQR = interquartile range; SD = Standard deviation. | | | | | | |
